# Lassa fever outbreaks, mathematical models, and disease parameters: a systematic review and meta-analysis

**DOI:** 10.1101/2024.03.23.24304596

**Authors:** Patrick Doohan, David Jorgensen, Tristan M. Naidoo, Kelly McCain, Joseph T. Hicks, Ruth McCabe, Sangeeta Bhatia, Kelly Charniga, Gina Cuomo-Dannenburg, Arran Hamlet, Rebecca K. Nash, Dariya Nikitin, Thomas Rawson, Richard J. Sheppard, H. Juliette T. Unwin, Sabine van Elsland, Anne Cori, Christian Morgenstern, Natsuko Imai-Eaton, the Pathogen Epidemiology Review Group

## Abstract

**Background:** Lassa fever, caused by Lassa virus (LASV), poses a significant public health threat in West Africa. Understanding the epidemiological parameters and transmission dynamics of LASV is crucial for informing evidence-based interventions and outbreak response strategies.

**Methods:** We conducted a systematic review (PROSPERO CRD42023393345) to compile and analyse key epidemiological parameters, mathematical models, and past outbreaks of LASV. Data were double extracted from published literature, focusing on past outbreaks, seroprevalence, transmissibility, epidemiological delays, and disease severity.

**Findings:** We found 157 publications meeting our inclusion criteria and extracted 374 relevant parameter estimates. Although LASV is endemic in West Africa, spatiotemporal coverage of recent seroprevalence estimates, ranging from 0.06% to 35%, was poor. Highlighting the uncertainty in LASV risk spatially. Similarly, only two basic reproduction number estimates at 1.13 and 1.19 were available. We estimated a pooled total random effect case fatality ratio of 33.1% (95% CI: 25.7 – 41.5, I^2^ = 94%) and found potential variation in severity by geographic regions typically associated with specific LASV lineages. We estimated a pooled total random effect mean symptom-onset-to-hospital-admission delay of 8.3 days (95% CI: 7.4 – 9.3, I2 = 92%), but other epidemiological delays were poorly characterised.

**Interpretation:** Our findings highlight the relative lack of empirical LASV parameter estimates despite its high severity. Improved surveillance to capture mild cases and approaches that integrate rodent populations are needed to better understand LASV transmission dynamics. Addressing these gaps is essential for developing accurate mathematical models and informing evidence-based interventions to mitigate the impact of Lassa fever on public health in endemic regions.

**Funding:** UK Medical Research Council, National Institute for Health and Care Research, Academy of Medical Sciences, Wellcome, UK Department for Business, Energy, and Industrial Strategy, British Heart Foundation, Diabetes UK, Schmidt Foundation, Community Jameel, Royal Society, and Imperial College London.

**Research in Context:** *Evidence before this study:* We searched PubMed up to August 2, 2023 for ((lassa fever) or (lassa virus)) and (epidemiology or outbreak or (models not image) or transmissibility or severity or delays or (risk factors) or (mutation rate) or seroprevalence). We found ten systematic reviews. Three on ribavirin as a Lassa fever treatment, two on Lassa virus (LASV) vaccine candidates, and one each on historical importations of Lassa fever cases from West Africa to non-endemic countries, clinical characteristics for protocol development, and Lassa fever in pregnancy. Two systematically reviewed epidemiological parameters. One on basic reproduction number estimates which ranged from 1.1 to 1.8 for human-to-human and 1.5 to 1.7 for rodent-to-rodent transmission. However, no meta-analyses were conducted. The other focused on LASV infection case fatality ratios (CFRs): 29.7% (22.3–37.5) in humans and prevalence: 8.7% (95% confidence interval: 6.8– 10.8) in humans, 3.2% (1.9–4.6) in rodents, and 0.7% (0.0–2.3) in other mammals. There were no systematic reviews on LASV transmission models.

*Added value of this study:* We provide a comprehensive overview of published outbreaks, transmission models and epidemiological parameters for LASV. We highlight the sparsity of key epidemiological parameter estimates such as the serial interval or generation time. The discrepancy between the high overall severity and the high seroprevalence in the general population suggests a high proportion of infections are asymptomatic or only result in mild disease. Therefore, current surveillance systems may need refining to better characterise LASV transmission dynamics.

*Implications of all the available evidence:* Epidemiological models are useful tools for real-time analysis of outbreaks, assessing epidemic trajectories and the impact of interventions. Our study is a useful basis to inform future LASV models, but highlights uncertainties and knowledge gaps that need to be filled in LASV transmission and natural history. Future LASV studies will benefit from integrating human and rodent reservoir surveillance.

## Introduction

Exemplified by the COVID-19 pandemic, the emergence and spread of high-consequence infectious disease pathogens continue to pose a global public health threat. Since 2018, the World Health Organization (WHO) R&D blueprint has defined a priority list of diseases likely to cause future epidemics, but for which medical countermeasures are limited, or not available. Lassa virus (LASV), a rodent-borne arenavirus, is the causative agent of the acute viral haemorrhagic illness Lassa fever (LF). LASV has the second highest spillover risk amongst viral pathogens and is on both the WHO and the Coalition for Epidemic Preparedness Innovations (CEPI) priority lists (1). Currently two of the four LASV vaccine candidates under development are in phase 1 trials, and whilst the antiviral ribavirin is commonly administered in Lassa endemic regions, its role in effective treatment of LASV is not clear (2–4).

LF was first described in humans in the 1950s, with the virus later identified in 1969 (5). It is endemic in Benin, Ghana, Guinea, Liberia, Mali, Sierra Leone, and Nigeria, and transmission is likely in other West African countries where *Mastomys* rodent reservoir hosts are present (6). Transmission to humans occurs through contact with infected animals or direct contact, inhalation, or ingestion of their excreta. Outbreaks due to human-to-human transmission can occur through direct contact with bodily fluids, contaminated medical equipment, or rarely via sexual contact. In endemic regions, larger outbreaks of LASV can occur seasonally as climate affects food availability and therefore rodent population sizes (7).

Although 80% of infections are asymptomatic or mild, with symptoms including fever, headache, nausea, and vomiting, a small proportion of patients develop severe disease which can be fatal or lead to long term sequelae such as deafness (8). Pregnant women in their 3^rd^ trimester are particularly at risk, with maternal death and/or adverse birth outcomes occurring in >80% of cases (8). Despite the reportedly low infection fatality ratio, LASV remains one of the highest-burden priority pathogens amongst the WHO R&D priority pathogens, with an estimated 300,000 to 500,000 cases and 5000 deaths annually in West Africa (9). Seven distinct lineages with substantial genetic divergence are typically present in specific geographic regions across West Africa, separated by environmental barriers such as rivers which limit the movement of the rodent reservoir (10–12).

Epidemiological models of disease transmission during outbreaks are used to understand the epidemic trajectory, severity, and the potential effect of interventions. These outputs support policymakers to respond to outbreaks in real-time by informing the allocation of resources and public policies. Despite robust epidemiological parameters estimates being critical for making reliable projections, at the beginning of an outbreak, parameters are often inferred from similar pathogens or drawn from the literature until enough data are collected to make estimates about the current outbreak. To address this knowledge gap, we conducted a systematic review of published studies to provide a comprehensive resource that synthesizes key epidemiological information for LASV.

## Methods

We followed the Preferred Reporting Items for Systematic Reviews and Meta-Analyses (PRISMA) guidelines, and we registered our study protocol with PROSPERO (International Prospective Register of Systematic Reviews) under the identifier #CRD42023393345.

### Search strategy and study selection

We searched PubMed and Web of Science for studies published from database inception up to March 8, 2019, the search was then repeated to include publications up to August 2, 2023. Results were imported into Covidence (2023) and de-duplicated (13). Titles and abstracts, and then full texts were independently screened by two reviewers (PD, DJ, TN, KM, RM, KC, GC-D, AH, HJTU, CM, NI) and conflicts resolved by consensus. Studies reporting on LASV past outbreak size, transmission models, viral evolution, transmission, natural history, severity, or seroprevalence in human populations were included. Non-peer reviewed literature and non-English language studies were excluded. See appendix A.1 for further details on study selection, table B.1 for full inclusion and exclusion criteria and section E for the PRISMA checklists.

### Data extraction

13% of the full texts that met the inclusion criteria were randomly selected and double extracted to validate the extraction process. A consensus on discordant results was established, after which 10 reviewers (PD, TN, KM, JTH, RM, GC-D, DN, TR, RS, CM) conducted single extraction on the remaining full texts. Data were extracted on publication details, risk of bias (see Data analysis), outbreak locations, dates, case numbers and deaths (as reported by articles – see appendix A.2), transmission model details, basic and effective reproduction numbers, epidemiological delays (such as the incubation period, or symptom-onset-to-hospital-admission or -outcome delays), case fatality ratios (CFRs), seroprevalence, and risk factors, using a Microsoft Access database (version 2305). As Lassa outbreaks are difficult to demarcate due to frequent rodent-human transmission in endemic areas, we only extract “outbreaks” or “epidemics” as described by study authors. For risk factors, only statistical significance and/or adjustment were extracted, as differences in reference groups and stratification made it challenging to compare other measures (e.g. odds ratios) across studies. Full details of the data extraction process (section A.2), database structure (tables B.2-B.5), and extracted data (tables B.6-B.12) are provided in the appendix.

### Data analysis

Meta-analyses of published estimates were conducted using the *meta* R package (14) for the CFR and the mean symptom-onset-to-hospital-admission delay. Fixed and random effects models were used to generate pooled CFR and delay estimates with 95% confidence interval (CI) and *I*^*2*^ estimates of heterogeneity (appendix A.3). For the CFR meta-analysis, we considered lineage regions (attributed based on the geographic area reported in the article, as phylogenetic analyses were not available in all cases), country, and sub-population affected (such as children or pregnant women) as potential drivers of differences in CFR. We did not perform meta-analyses for other epidemiological delays, such as the incubation period, due to insufficient numbers of estimates of central tendency paired with sample uncertainty. Risk of bias was assessed using a custom-designed quality assessment questionnaire and quality assessment scores were calculated as the mean number of ‘Yes’ responses to the seven questions (excluding NA responses). A local polynomial regression fit was used to analyse trends in quality assessment scores by publication year (appendix figure C.1). We excluded systematic reviews from our study but used them to cross-check that all eligible studies were included (appendix F). All analyses were conducted in *R* (version 4.2.2) and the code to reproduce our results is available at https://github.com/mrc-ide/priority-pathogens. Curated data on outbreaks, models, and epidemiological parameters were added to the *epireview* R package (15) (appendix D).

## Results

The search returned 5,414 potentially relevant articles. After de-duplication 2,685 articles were screened with 2,215 excluded at the title/abstract stage. 470 studies were retained for full text screening with 157 studies meeting the criteria for final inclusion. Three studies with no full text were excluded (Figure 1). 24 studies reported 18 distinct LASV outbreaks, with the highest number of outbreaks (n) reported in Nigeria (n = 9), Sierra Leone (n = 4), Liberia (n = 3), and Benin (n = 2) between 1969 and 2019 (Appendix table B.6). Confirmed case numbers reported in studies ranged from 2 cases in Benin (16) to 1463 cases in Nigeria during a multi-year outbreak in 2018-19 (17).

**Figure 1:**
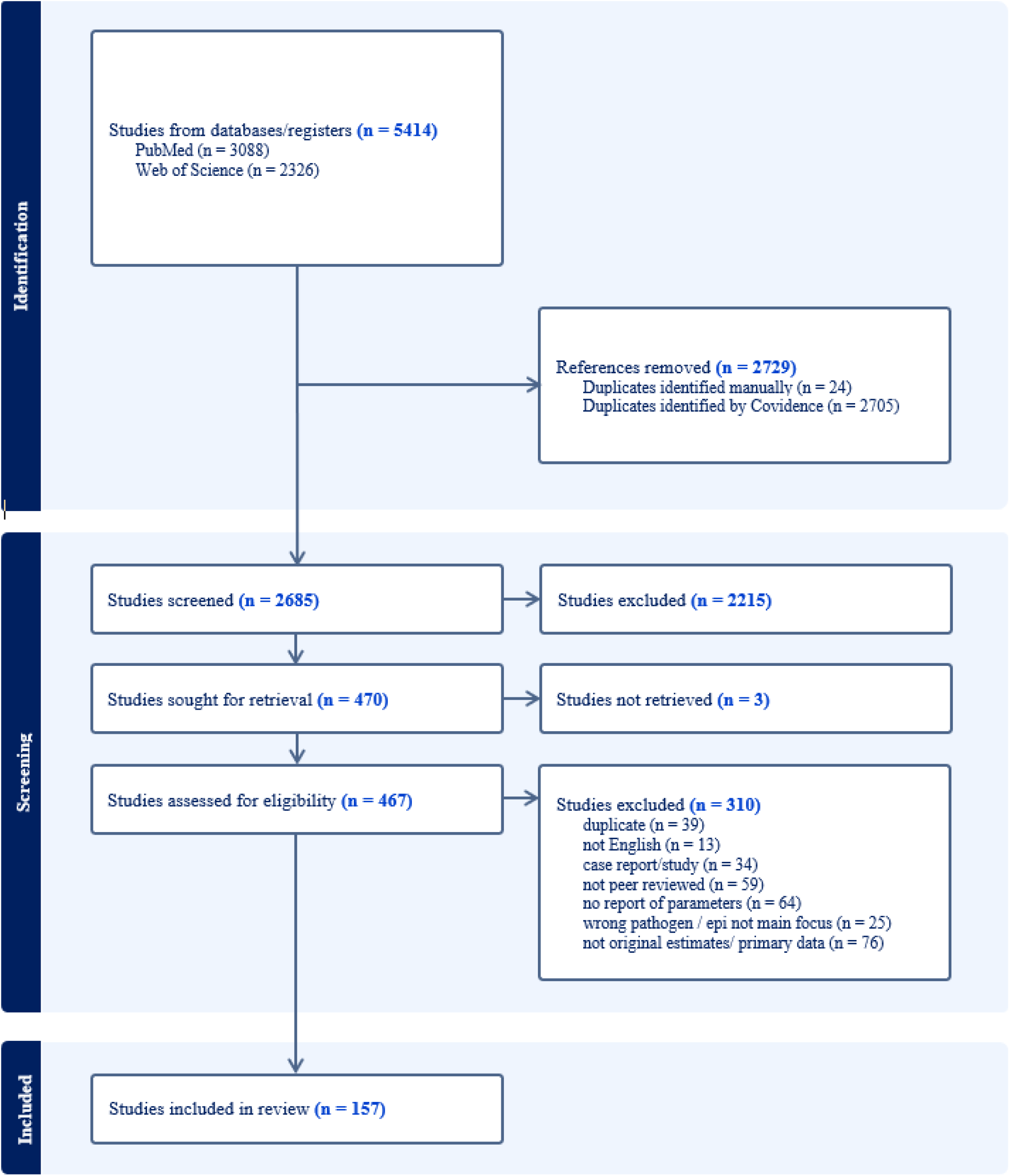
Study selection according to PRISMA guidelines and criteria as described in Table B.1. (Reasons for abstract exclusion not provided by Covidence).

34 studies employed LASV transmission models, the majority of which were compartmental models and accounted for both rodent-human and human-human transmission pathways (Appendix figure C.2).

We extracted 371 epidemiological parameter estimates in total including 123 seroprevalence estimates from 29 countries, risk factors (n=105) and severity or CFR estimates (n=72) (Appendix figure C.3).

Figure 2A shows that for population- and community-based studies between 1965 – 1992, aggregated seroprevalence estimates ranged from 0.03% (Haut-Ogooue in Gabon) to 30.5% (Eastern Province in Sierra Leone). The underlying data ranges from 0% (Kenya (not plotted) (18) and Gabon (19)) to 52% (Sierra Leone (20)). Seroprevalence estimates using IgG assays from more recent studies (1993-2018) were only available from 7 countries where reported seroprevalence was generally higher (Figure 2B). No seroprevalence estimates were available from Chad, Cameroon, Central African Republic, Republic of Congo, Gabon, or Equatorial Guinea, where serosurveys were conducted pre-1993 (Figure 2A). Due to differences in assays and exact geographic locations of the study, temporal trends in population seroprevalence cannot be inferred. Figure 2B shows evidence of past infection in areas of known outbreaks, but also highlights high seroprevalence in Bougouni District in Mali, and Abidjan and Northeastern Region in Cote d’Ivoire, despite a lack of reported cases or outbreaks, indicating underreporting of LASV cases or transmission.

**Figure 2:**
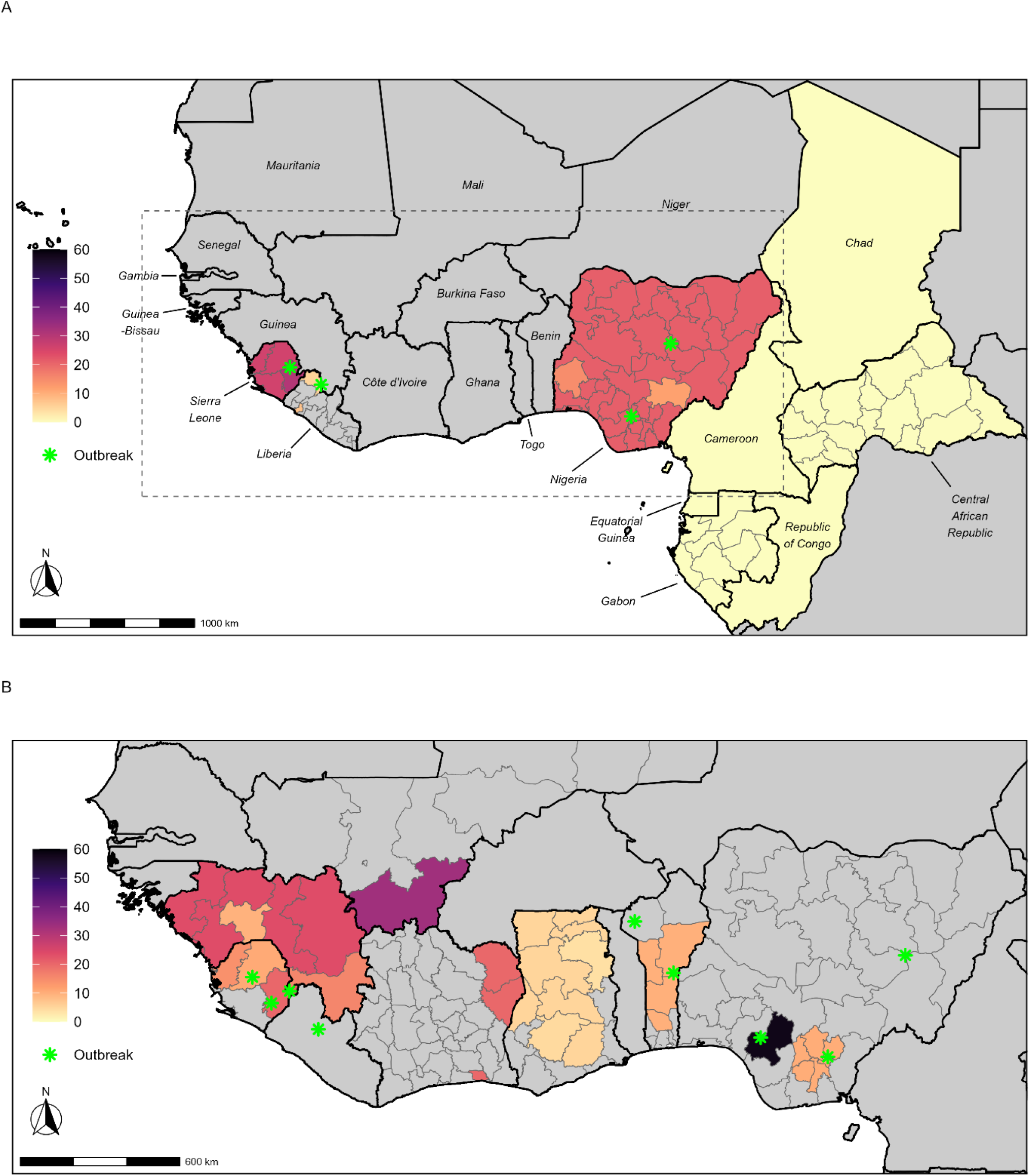
LASV seroprevalence (%) in population and community settings (general population) of West Africa. (A) Mean LASV seroprevalence (%) from studies conducted between 1965 and 1992 (target antibodies not specified in the papers). (B) Mean LASV seroprevalence (%) using IgG assays from studies conducted between 1993 and 2018. Subnational estimates are shown at administrative level 1. Green asterisks show reported LASV outbreak locations and grey areas show absence of estimates. The dotted grey box in panel (A) shows the corresponding area depicted in panel (B).

72 CFR estimates (n) were reported across 58 studies between 1970 and 2021 in Nigeria (n=34), Sierra Leone (n=22), Liberia (n=12), Guinea (n=2), and Benin (n=1), with one joint estimate across Guinea, Liberia, and Sierra Leone, in the LASV lineage IV region (Fig. 3B). We estimated a pooled CFR of 23.1% (95% CI: 22.1 – 24.1, *I*^*2*^ = 94%) for total common effect and 33.1% (95% CI: 25.7 – 41.5, *I*^*2*^ = 94%) for total random effect models. CFR estimates differed most notably by lineage regions or geographic areas associated with the transmission of different genetic lineages of LASV (Figure 3A, B), with the highest CFR in the lineage III region at 54.5% (95% CI: 46.3-62.5%). Lineage regions II and IV reported broadly consistent CFRs. There was a single study reporting a CFR of 44% within lineage region VII (16). CFR estimates did not differ significantly when comparing random effects across countries, but CFR in Sierra Leone was significantly higher than in Nigeria and Liberia using the common effects model (Figure 3C). CFR estimates for studies reporting more than 1000 cases were based on large retrospective studies conducted by Nigeria Centre for Disease Control and Prevention which reported cases and deaths over multiple years. Due to the endemic nature of LASV in Nigeria, these CFR estimates likely represent different circumstances to more acute and smaller outbreaks typical of other settings with a higher CFR. This is reflected in Figure 3D which shows a decreasing trend in CFR as reported case numbers increase. Common effect model CFR for <30 reported cases is significantly higher than estimates from studies with larger case numbers. Sensitivity analyses performed without deduplicating the CFR estimates produced similar results (Appendix Figures C.10-C.11).

**Figure 3:**
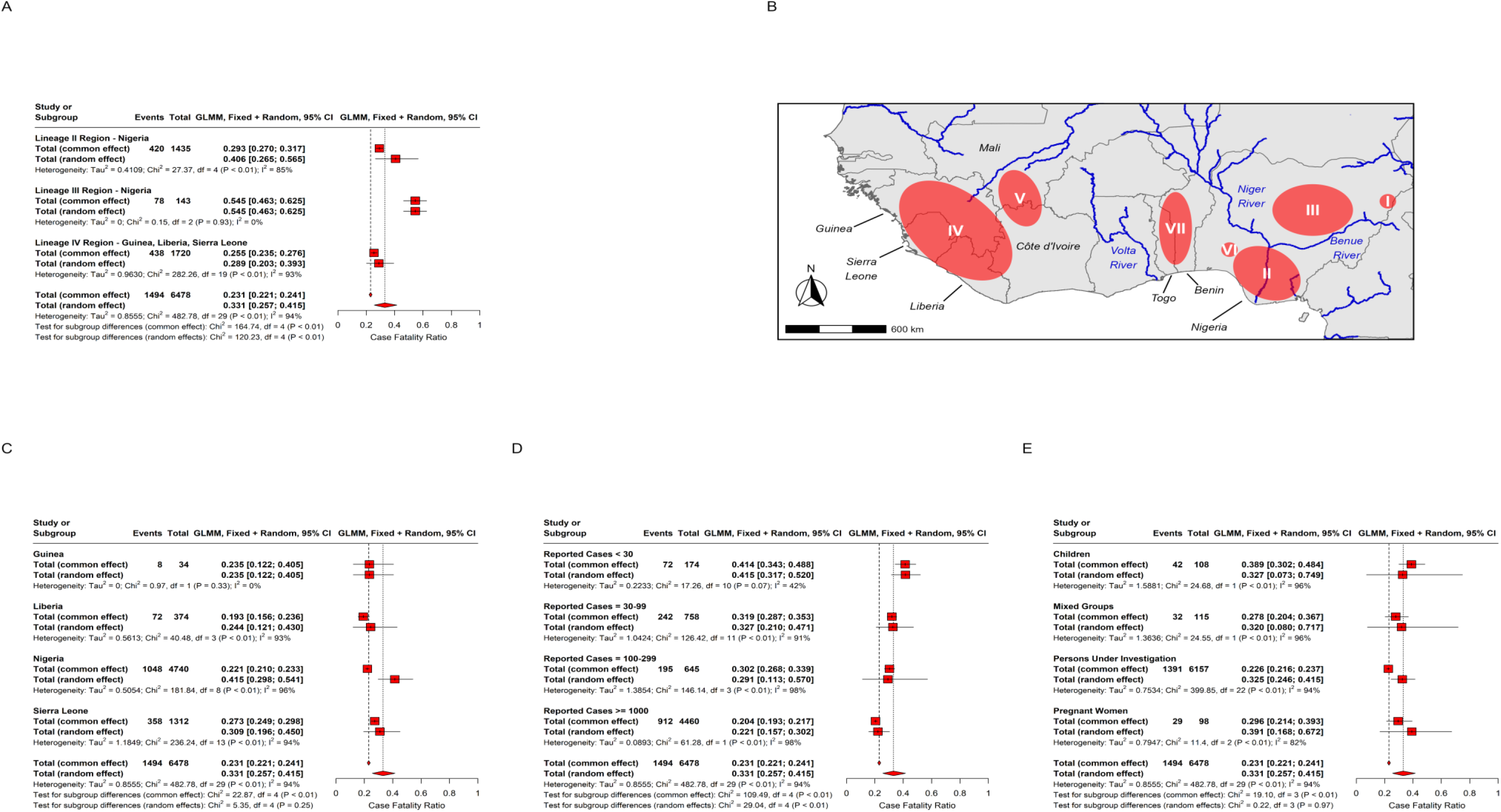
Case fatality ratio (CFR) meta-analyses using logit-transformed proportions and a generalised linear mixed-effects model. (A) CFR estimates by LASV lineage region, inferred from location as reported in each study. (B) Reported geographic distribution of LASV lineages. (C) CFR estimates by country. (D) CFR estimates by total number of reported cases. (E) CFR estimates by population group under study. Red squares show total common and random effects by subgroup and diamonds show an overall common effect estimate, where data are pooled and assumed to come from a single data-generating process with one common CFR, and an overall random effect estimate, which allows the CFR to vary by study and assigns different weights when calculating an overall estimate (appendix section A.3). The number of events corresponds to the number of reported deaths. Individual study estimates and further subgroup decompositions are shown in appendix figures C.5-C.9.

We extracted 50 estimates of epidemiological delays from 24 studies (Figure 4), including five incubation period estimates, 18 symptom-onset-to-hospital-admission delays, nine admission-to-clinical-outcomes delays, and nine symptom-onset-to-clinical-outcome delays. These estimates were a mixture of means, medians, and ranges with one unspecified incubation period estimate. The five incubation periods were broadly consistent across the studies with central estimates ranging between 7 and 12.8 days. Hospital-admission-to-death delays were consistent across the four estimates and were shorter than admission-to-recovery (three estimates) or admission-to-other-unspecified-outcomes (two estimates) (Figure 4C). Mean symptom-onset-to-admission delays included in the meta-analysis ranged from 6 to 11 days (Figure 4E). Pooled mean symptom onset to admission estimates were 8.6 days (95% CI: 8.3 – 8.8, *I*^*2*^ = 92%) for total common effect and 8.3 days (95% CI: 7.4 – 9.3, *I*^*2*^ = 92%) for total random effect models (21).

**Figure 4:**
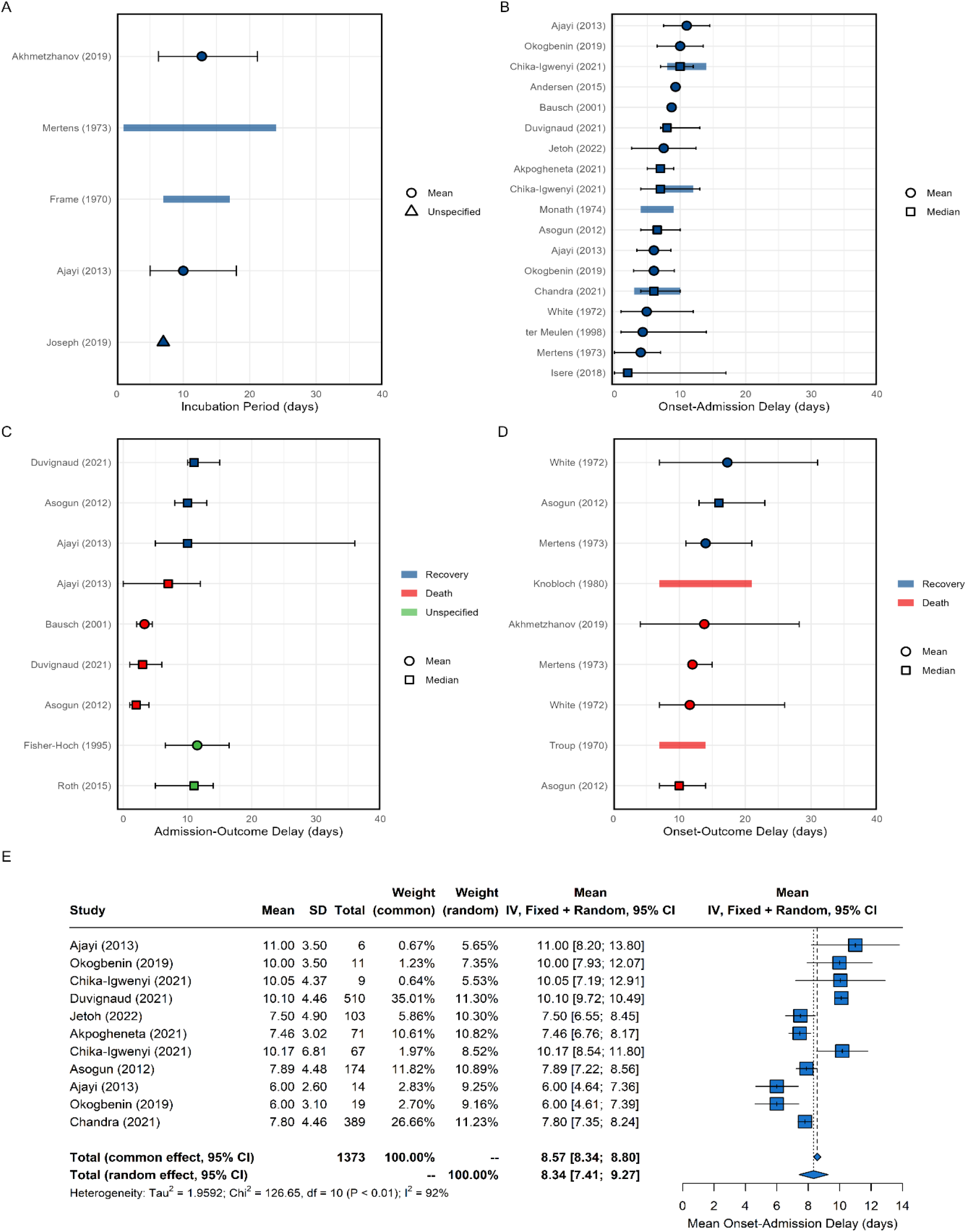
Overview of LASV epidemiological delays and mean symptom onset to hospital admission delay meta-analysis. (A) Estimates of the incubation period. (B) Estimates of the symptom onset to hospital admission delay. (C) Estimates of hospital admission to recovery, death, or unspecified outcome delay. (D) Estimates of symptom onset to recovery or death delay. Circles, squares and triangles represent mean, median and unspecified estimates respectively. Thin solid lines are uncertainty intervals and thick shaded lines the range across central estimates of disaggregated parameters (e.g., regions, time, age, sex). (E) Meta-analysis of mean symptom onset to hospital admission delay. Blue squares are individual study estimates and diamonds show common and random effect estimates. Hypothetical symptom onset to hospital admission delay probability distributions are shown in Appendix Figure C.4.

We extracted information on a range of LASV transmission parameters (Figure 5), including four studies reporting LASV reproduction numbers, one study reporting on growth rates, three studies on attack rates, and two estimating the percentage contribution of human-human (as opposed to the rodent-human) transmission. Three studies (1 on genes and 2 on segments) reporting LASV evolutionary and substitution rates were largely consistent (Fig. 5E, F). Two basic reproduction number estimates were available from two studies, with similar results at 1.13 and 1.19 (22,23). Two other studies reported effective reproduction numbers, 0.73 for nosocomial transmission (24) and between 1.0 to 1.6 (25). However, due to the consistent spillover from the rodent population, it is difficult to interpret the reproduction number estimates as the actual risk of sustained human transmission, as highlighted by the three mathematical modelling studies estimating that human-human transmission contributes to less than a third of transmission (24). Furthermore, the few available attack rate estimates are also low at between 2.2 – 3.1 per 1000 for primary (26,27) and 0.56% for secondary attack rates (28).

**Figure 5:**
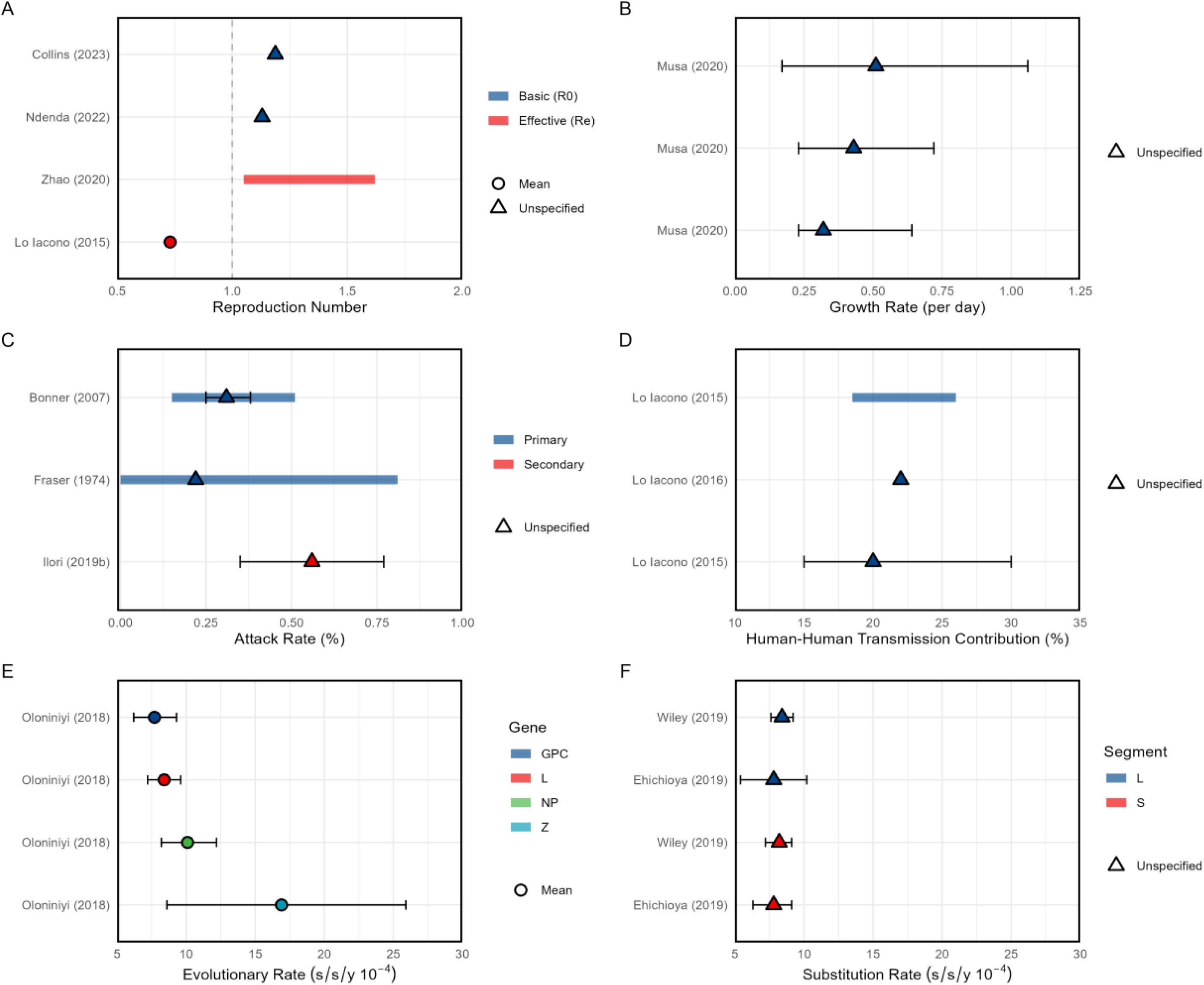
Overview of LASV (A) reproduction numbers, (B) growth rates, (C) attack rates, (D) percentage human to human transmission contribution, LASV genome (E) evolutionary rates for each protein-encoding gene and, (F) substitution rates for both genome segments. Both E and F are measured in nucleotide substitutions per site per year (s/s/y). Circles and triangles represent mean (panels A and E) and unspecified (panels A-F) estimates respectively. Thin solid lines are uncertainty intervals and thick shaded lines the range across central estimates of disaggregated parameters (e.g., regions, time, age, sex).

We also extracted reported risk factors for seropositivity, infection, severe disease or hospitalization, deaths and other outcomes (Appendix Table B.12).

In assessing risk of bias, descriptive studies (e.g. seroprevalence studies or articles not using a mathematical model) account for the majority of non-applicable responses, as per the design of the quality assessment questionnaire. We find that the LOESS curve fitted to the quality assessment score, excluding mathematical modelling studies, monotonically increased since 1970 (appendix figure C.1). In general, we found that data extractors assigned lower quality score to modelling studies published after 2017, potentially because most of these studies presented transmission models that were not fitted to data (appendix figure C.2).

## Discussion

In this systematic review, we compiled and analysed key epidemiological parameters, mathematical models, and past outbreaks of LASV that can inform future models of LASV transmission. Despite the wide distribution of environmental suitability of LASV transmission across West Africa (29,30), the geographic distribution of empirical LASV outbreak or seroprevalence data are very limited with 29 outbreaks described in 22 papers. Previous studies suggest this is due to lack of effective diagnostics and a bias towards areas with established LF treatment units (30,31). Furthermore, the spatial distribution of LASV remains poorly characterised with a paucity of modern serosurveys. The recent identification of new reservoirs beyond *M. natalensis* including domestic animals, farm animals, reptiles, and other rodent species adds to this uncertainty in the true distribution (32). Spatiotemporal modelling studies have combined predictions of rodent reservoir range with human LASV seroprevalence data to estimate geographic patterns of spillover risk (33). While some statistical models suggest that LASV risk extends into Central African countries, conflicting evidence exists, and our results show that recent human seroprevalence data from this region are lacking (29,33–35). The highly spatially heterogeneous distribution of LASV may also be due to differences in potential human-rodent interactions, which are difficult to capture. This poses challenges for surveillance and highlights the need for improved approaches, such as integrating human and peri-domestic rodent surveillance, especially given the increasing numbers of reported LASV cases and deaths in Nigeria (36,37). Furthermore, climate and land-use change and their impact on the rodent reservoir are anticipated to influence the future spatial spread and extent of LASV transmission (7). Spatial and environmental modelling suggests that the LASV endemic area will spread, increasing the population at risk of infection (35). However, change in LASV spillover risk will not be spatially uniform (38). Therefore, a greater understanding of how individual reservoir hosts respond to environmental stressors is needed to accurately predict any future changes in transmission risk.

Our overall pooled CFR estimates were consistent with previous LASV reviews and with CFR estimates for other rodent-borne arenaviruses causing haemorrhagic fevers, such as Machupo virus (39) and Guanarito virus (40), with slightly lower CFRs observed in Junín virus (41). However, there remains an apparent contradiction between high LASV CFR estimates and the high seroprevalence in the general population which suggests a high proportion of infections are asymptomatic or only result in mild disease (20). As many of the CFR estimates were based on patients seeking healthcare and thus by definition having more severe disease, it is likely that these high severity estimates are biased. Simons (2022) has also shown that LF burden and cases are systemically underreported in countries such as Ghana, Guinea, and Togo where LF surveillance is not routine but also in countries like Nigeria (42). Therefore, characterising these asymptomatic or mild cases will be critical for refining CFR estimates and for developing more robust models of LASV transmission which will help inform the design of LASV vaccine trials (43). The evolutionary dynamics of LASV, including the emergence of distinct LASV lineages are important considerations for understanding variations in disease severity. Our results suggest that CFRs may differ by lineage with the highest CFR estimates derived from outbreaks in the lineage III region in Nigeria. However, we attributed these lineage regions based on the geographic area reported in the paper and not on specific phylogenetic analyses of the outbreaks. Past papers have shown that these lineage regions are distinct due to different physical environmental barriers such as the Niger and Benue rivers restricting the movement of the reservoir populations (11,12). Gomerep et al. have also suggested that differences in apparent severity by lineage region may also be attributable to variable health-seeking behaviours by area (44). Published estimates of LASV evolutionary rates are low and Klitting et al. have also shown slow rates of LASV circulation with a weighted lineage dispersal velocity between 0.8 and 1.0 km/year (35)

We only found 5 estimates of the incubation period which reflects the challenges in attributing likely exposure times due to the complex transmission dynamics and the multiple potential routes of infection. A much greater number of delay distributions were available for those related to hospital admissions and treatment. Whilst the symptom onset or hospital admission to outcome (recovery or death) delays were broadly consistent across studies, these observed delays are conditional on cases having severe disease and these delay distributions will likely differ for mild cases. There were also a limited number of transmission parameter estimates available in the published literature with only two estimates of the basic reproduction number (*R*_*0*_ =1.13 and 1.19) and two studies reporting estimates of the effective reproduction number (*R*_*e*_) with Zhao et al. estimating a range of Re between 1.06 and 1.62 (25).

There were very few mathematical models fitted to empirical data, with the majority of identified papers describing theoretical models of transmission. This reflects the challenges in parameterising complex reservoir-human meta-population models and makes it difficult to robustly project the potential impact of different interventions such as the use of ribavirin or targeted vaccination campaigns (45). Modelling frameworks developed to explore potential vaccine demand for CEPI priority pathogens have highlighted surveillance and data gaps in the true geographic distribution of LASV, the key routes of rodent-human spillover, and the role of human-human transmission outside nosocomial transmission (46). With *R*_*0*_ estimates slightly >1 LASV has the potential to spillover and sustain transmission in human populations, but that the human-human transmission risk is highly variable. The three available estimates of the relative contribution of human-human transmission suggests that it contributes less than a third of the overall transmission events and that spillover from the rodent population is the largest cause of human infections (24). The lower levels of human-to-human transmission and the challenges associated with quantifying this may also explain the absence of generation time or serial interval estimates. *M. natalensis* is the primary reservoir host for LASV and play a crucial role in the virus’s transmission dynamics. Integrating rodent seroprevalence studies with human seroprevalence data will provide insights into the spatial and temporal patterns of LASV circulation and will be important in developing mathematical models of LASV transmission. However, in this review we focused on the human hosts and hence excluded seroprevalence studies in rodents.

Our findings are also limited by our exclusion of pre-prints, grey literature, and non-English language publications. This language limitation is particularly important in the context of the LASV endemic region, which includes many non-English speaking countries. Where studies reported a parameter of interest disaggregated by factors such as region, sex, or time we chose to extract the overall results for pragmatic reasons. Information on which studies report such data is marked in our database in the *epireview* R package. We also limited our data extraction to results in tables or the main text and hence did not extract any results or data that were presented only in figures. However, the number of results represented solely in figures likely represents a very small proportion of papers. Finally, although we validated the data extraction process small discrepancies between individual extractors cannot be ruled out.

We have provided a comprehensive overview of key epidemiological parameters, mathematical models, and past outbreaks of LASV, aiming to inform future models of LASV transmission and enhance our understanding of the disease dynamics. Several significant gaps remain, with significant uncertainty in the true spatial distribution and burden of LASV and its reservoirs. This underscores the need for improved surveillance methods and integrated approaches, especially considering the increasing burden reported in endemic regions. Additionally, the anticipated impact of climate and land-use change on LASV transmission highlights the importance of understanding how environmental factors influence disease dynamics. Addressing these limitations and incorporating a broader range of data sources will be essential for advancing our knowledge of LASV transmission dynamics and mitigating the impact of LF on public health. We encourage readers to contribute to the *epireview* package which will ensure that the latest available LASV epidemiological information is publicly and easily accessible.

## Data Availability

All data produced are available online at https://github.com/mrc-ide/priority-pathogens and https://github.com/mrc-ide/epireview

https://github.com/mrc-ide/epireview

## Declarations

### Availability of data and materials

https://github.com/mrc-ide/epireview/tree/main/data

### Code availability

https://github.com/mrc-ide/priority-pathogens, https://github.com/mrc-ide/epireview

### PROSPERO

CRD42023393345

(https://www.crd.york.ac.uk/prospero/display_record.php?RecordID&RecordID=393345)

### Authors’ contributions

SB, SvE, AC, and NI-E conceptualised this systematic review. PD, DJ, KC, AH, CM, and NI-E searched the literature and screened the titles and abstracts. PD, DJ, TN, KM, RM, KC, GC-D, HJTU, CM, and NI-E reviewed all full-text articles. PD, TN, KM, JTH, RM, GC-D, DN, TR, RS, and CM extracted the data. PD, DJ, CM, and NI-E did formal analysis of, visualised, and validated the data. PD, SB, and RKN were responsible for software infrastructure. AC acquired funding. GC-D, SB, SvE, AC, CM, and NI were responsible for project administration. PD, GC-D, RKN, DN, HJTU, and SvE were responsible for training individuals on and accessing Covidence and designing the Access system. CM and NI-E supervised the systematic review. PD, DJ, CM, and NI-E wrote the original draft of the manuscript. All authors were responsible for the methodology and review and editing of the manuscript. All authors debated, discussed, edited, and approved the final version of the manuscript. All authors had final responsibility for the decision to submit the manuscript for publication.

