## Supplementary Information for "Lassa fever outbreaks, mathematical models, and disease parameters: a systematic review and meta-analysis"

March 2024

**B Additional Tables** **4**

**C Additional Figures** **22**

**D epireview** **33**

**E PRISMA 2020 Checklists** **34**

**F Other Systematic Reviews** **37**

**G Excluded Studies** **38**

**H Pathogen Epidemiology Review Group (PERG) Membership** **66**

### A Additional Information on Methods

#### A.1 Study Selection

Original research papers in English were included if reporting on Lassa virus (LASV) transmission, evolution, natural history, severity, seroprevalence, size of previous outbreaks or published mathematical transmission models. Non-peer reviewed literature was excluded. Papers identified in the search were imported into *Covidence*, a software program used to manage systematic reviews. From a team of seven reviewers, two independent reviewers first screened titles and abstracts then full texts to assess eligibility for data extraction. Disagreements in eligibility determination were resolved by consensus between the two independent reviewers.

**Search term:** `lassa AND ((transmission OR epidemiology) OR (model* NOT imag*) OR (burden OR severity OR "case fatality ratio" OR CFR) OR ("serial interval" OR "incubation period" OR "generation time") OR ("heterogeneity" OR "superspread*") OR ("reproduction number" OR "reproductive number" OR "R0") OR ("pre-existing immunity" OR "serological" OR "serology" OR "serosurveys") OR (diagnostic OR diagnosis OR test*) OR ("evolutionary rate" OR "genetic mutation" OR evolution) OR (outbreak OR cluster OR epidemic) OR ("risk factor*") OR ("case definition"))`

#### A.2 Data Extraction

##### Outbreaks

It is difficult to define and demarcate outbreaks of LASV, given its endemicity and the frequency of rodent-human transmission events. Therefore, we only extract "outbreaks" or "epidemics" as described by study authors. Possible methods of case confirmation include rapid diagnostic tests (RDTs), polymerase chain reaction (PCR) tests or serological assays.

##### Models

Details of disease transmission models were extracted, including model type, whether it modelled deterministic or stochastic processes, transmission pathways included, and in the case of compartmental models, human health states (e.g., SIR, SEIR). Furthermore, we extracted information on underlying model assumptions, whether the model is theoretical or fitted to data, interventions considered and the availability of model code.

##### Parameters

For each parameter, we extracted all available information, including types of values (e.g., mean, median, standard deviation), uncertainty intervals (capturing the precision of estimates), and ranges (if, e.g., multiple estimates were obtained from different populations or using different methods). Study context for parameter values included survey location and dates, sample size, basic demographic information, and timing of the survey in relation to reported outbreaks.

For genomic data, we noted the gene studied if specified and if newly sequenced data were available. For reproduction numbers, we recorded the methods used for estimation (e.g. renewal equations, compartmental models, empirical methods). For case fatality ratios, we extracted whether the estimation approach accounted for cases with unknown final status or not [1]. For both case fatality ratios and seroprevalence, we additionally recorded numerators and denominators where available.

For risk factors, we extracted the outcome (e.g. infection or death), the risk factor for that outcome (e.g. age, sex or occupation), the type of occupation if specified, and whether the risk factor(s) estimates were statistically significant and/or adjusted. We chose not to extract odds ratio estimates because studies may have used different stratifications or reference groups, making it challenging to compare values across studies. The information we extract offers an overview of risk factors explored across studies that may affect the risk of infection and death, which may be useful to consider when designing LASV transmission models.

#### A.3 Meta-Analysis

The meta-analysis of CFRs and the mean onset-admission delay (in Figures 3 and 4 in the main text, respectively) followed a standard approach and was conducted using the `meta` R package [2].

A mixed-effects model is a linear model $y_i = \beta_0 + \sum_{j=1} \beta_j x_{ij} + u_i + \epsilon_i$, where $y_i$ are the observed data, $x_{ij}$ any explanatory variable, $\beta_j$ are the fixed effects, $u_i$ are the random effects (centered around zero and independent across $i$), and $\epsilon_i$ are error terms. Meta-analysis is a special case of the above mixed-effects model with only an intercept term $\beta_0$ (a fixed-effects/common-effects model) or only an intercept term $\beta_0$ and a random-effects term $u_i$ associated with that intercept (a random-effects model).

For the CFR meta-analysis, a generalised logistic mixed-effects model was used, in which the individual CFR estimates are transformed using the logit-transformation $y_i = \log\left(\frac{CFR_i}{1-CFR_i}\right)$ to ensure that the distribution was approximately normal, and then a generalised linear mixed-effects model is applied to the transformed CFRs to estimate the pooled effect. A comprehensive overview of the methodology is provided by [3].

In Figure 3 of the main text, we perform a sub-group analysis of CFRs, in which estimates are grouped by lineage region, country, number of reported cases, and population group. The meta-analysis in panels A, C, D, and E reports the common and random effects for each sub-group to investigate any patterns between groups. We note that

- The overall common and random effects are the same across all different sub-group analyses, as the underlying estimates/studies are the same.
- We provide the same meta-analysis results plotting all underlying CFR estimates in figures C.5 to C.9.
- In Figure 3, we have removed both *known* and *assumed* duplicates in the CFR data. CFRs are considered known duplicates if they refer to the same underlying data (e.g. naive and adjusted CFRs, a subset of reported cases). CFRs are considered assumed duplicates if the report CFRs based on overlapping locations or time periods, for example. To assess the sensitivity of the results, we provide a CFR meta-analysis including known duplicates in figure C.10 and for all CFR data in figure C.11. We note that there is no substantial difference in the total common and random effects.

The mean onset-admission delay meta-analysis in Figure 4E requires paired mean and standard deviation estimates of the onset to admission delay. However, the underlying studies report several different parameter value types (e.g., means, medians, standard deviations, interquartile range, minimum/maximum), so we use the functionality of *metamean*, which is implemented in the `meta` package, to infer the means and standard deviations for each study. In particular, we use the method provided by Cai et al (2021) [4], which allows for unknown non-normal distributions.

In Figure C.4, we use the overall common and random effects, and the calculated standard error thereof, to display the probability density function for common probability distributions for delays (Gamma, Lognormal and Weibull).

The code to reproduce our results is available at https://github.com/mrc-ide/priority-pathogens.

### B Additional Tables

| **Inclusion Criteria** | **Exclusion Criteria** |
|---|---|
| Language: English | Language: not English |
| Form/Study Type: peer-reviewed, original research | Form/Study Type: not peer-reviewed (posters, conference proceedings, correspondence), not original research (literature reviews, meta-analyses) |
| Participants/Population: human (genomic studies with animal samples are included provided that the majority of samples are human), sample size $\geq 2$, case series[1] | Participants/Population: in-vitro studies, solely animal studies, sample size $< 2$, i.e. case reports |
| Contents: any one of the following;<br>(A) mention of any human outbreak – size, year, location, duration, spatial scale (local, regional, national, international)<br>(B) mathematical or statistical model of transmission<br>(C) measures/estimates of human: R, R0, Rt, r, Re, overdispersion, growth rate, attack rate, relative ratio of human-human vs. animal transmission, evolutionary rate, mutation rate, substitution rate, generation time, serial interval, incubation/latent period, other human delays, CFR, seroprevalence, risk factors (risk and the measure) | Contents: pathogen not primary focus of study, no inclusion criteria satisfied, qualitative studies (e.g., KAP studies) |

Table B.1: Inclusion and exclusion criteria.

| **Theme** | **Question** |
|---|---|
| Is the methodological/statistical approach suitable? (how the data are used) | 1. Clear and reproducible<br>2. Robust and appropriate for the aim [subjective criteria] |
| Are the assumptions appropriate? (input parameters/assumptions - what goes into the methodology) | 3. Clear and reproducible<br>4. Justified (published study or analysis of data)[objective criteria] |
| Are the data appropriate for the selected methodological approach? | 5. Clearly described and reproducible<br>6. Are issues in the data clearly discussed and acknowledged?<br>7. Are issues in the data accounted for in the chosen methodological approach? |

Table B.2: Quality assessment questionnaire: possible responses for each question listed were Yes, No, and NA (non-applicable).

| Data field | Variable name | Expected data type | Notes |
|---|---|---|---|
| Article ID | article_id | integer | ID to connect to article form |
| Outbreak ID | outbreak_id | integer | ID assigned by database |
| Outbreak start day | outbreak_start_day | integer | Day of outbreak start if reported |
| Outbreak start month | outbreak_start_month | character | Month of outbreak start if reported |
| Outbreak start year | outbreak_start_year | integer | Year of outbreak start if reported |
| Outbreak end day | outbreak_end_day | integer | Day of outbreak end if reported |
| Outbreak end month | outbreak_end_month | character | Month of outbreak end if reported |
| Outbreak end year | outbreak_date_year | integer | Year of outbreak end if reported |
| Outbreak ongoing | ongoing | logical | Tick box to indicate if the outbreak is reported to be ongoing at the time of publication |
| Duration (months) | outbreak_duration_months | integer | Duration of outbreak in months if reported. No calculation of duration is done |

[1] As stated in the PROSPERO registration for the overall priority pathogen project, the inclusion of case reports and case series are determined on a case-by-case basis for each pathogen.

| | | | |
|---|---|---|---|
| Outbreak country | outbreak_country | character | Country or countries where the outbreak took place - from dropdown list |
| Outbreak location | outbreak_location | character | Region/district/province/city where the outbreak took place |
| Pre-outbreak endemicity | pre_outbreak | character | Endemicity of the pathogen in the location pre-outbreak if reported |
| Cases suspected | cases_suspected | integer | Number of suspected cases as reported |
| Cases confirmed | cases_confirmed | integer | Number of confirmed cases as reported |
| Mode of detection of cases | cases_mode_detection | character | Method for case detection - from dropdown list |
| Asymptomatic transmission described | asymptomatic_transmission | logical | Tick box whether asymptomatic transmission is described in paper or not. |
| Asymptomatic cases | cases_asymptomatic | integer | Number of asymptomatic cases as reported |
| Severe cases | cases_severe | integer | Number of severe or hospitalized cases as reported |
| Deaths | deaths | integer | Number of deaths as reported |

Table B.3: Outbreak data fields: refer to epireview in Supplement D for dropdown options.

| Data field | Variable name | Expected data type | Notes |
|---|---|---|---|
| Article ID | article_id | integer | ID to connect to article form |
| Model data ID | model_data_id | integer | ID assigned by database |
| Model type | model_type | character | General type of model - from dropdown list |
| Stochastic or deterministic | stoch_deter | character | Stochastic or deterministic model as reported |
| Transmission route | transmission_route | character | Transmission route(s) modelled - from dropdown list |
| Assumptions | assumptions | character | General assumptions for the model - from dropdown list |
| Compartmental type | compartmental_type | character | Specific type of compartmental model - from dropdown list |
| Theoretical model | theoretical_model | logical | Tick box whether the model was fitted to data (NA) or just theoretical (TRUE) |
| Intervention type | interventions_type | character | Type of intervention(s) modelled - from dropdown list |
| Code available | code_available | logical | Tick box whether code for the model was publicly available and reported in paper |

Table B.4: Model data fields: refer to epireview in Supplement D for dropdown options.

| Data field | Variable name | Expected data type | Notes |
|---|---|---|---|
| Article ID | article_id | integer | ID to connect to article form |
| Parameter data ID | parameter_data_id | integer | ID assigned by database |
| Parameter type | parameter_type | character | Category of parameter - see dropdown list |
| Parameter value | parameter_value | numeric | Central parameter value |
| Parameter exponent | exponent | integer | Parameter value exponent (base 10) |
| Inverse parameter | inverse_param | logical | Tick box to indicate that only inverse of parameter is reported (e.g., recovery rate from fitted model instead of infectious period) |
| Parameter unit | parameter_unit | character | Units for parameter value, applies to central estimate and ranges/uncertainty intervals - see dropdown list |
| Parameter value type | parameter_value_type | character | Type of central parameter value - see dropdown list |
| Parameter lower bound | parameter_lower_bound | numeric | Lower bound of the parameter range if a range was reported or if data are disaggregated |
| Parameter upper bound | parameter_upper_bound | numeric | Upper bound of the parameter range if a range was reported or if data are disaggregated |

| | | | |
|---|---|---|---|
| Parameter uncertainty - single type | parameter_uncertainty_single_type | character | Type of uncertainty for the central parameter value if a single value was reported - see dropdown list |
| Parameter uncertainty - single value | parameter_uncertainty_single_value | numeric | Value for uncertainty for the central parameter value if a single value was reported (e.g., the value of the std. dev.) |
| Parameter uncertainty paired type | parameter_uncertainty_type | character | Type of uncertainty for the central parameter value if paired values were reported - see dropdown list |
| Parameter uncertainty - lower value | parameter_uncertainty_lower_value | numeric | Lower bound for uncertainty for the central parameter value if paired values were reported |
| Parameter uncertainty - upper value | parameter_uncertainty_upper_value | numeric | Upper bound for uncertainty for the central parameter value if paired values were reported |
| Distribution type | distribution_type | character | Type of distribution for the estimated parameter - see dropdown list |
| First distribution parameter type | distribution_par1_type | character | Type of value for the first distribution parameter - see dropdown list |
| First distribution parameter value | distribution_par1_value | numeric | Value for the first distribution parameter (e.g., shape or scale parameter for a gamma distribution) |
| First distribution parameter uncertainty | distribution_par1_uncertainty | logical | Tick box for whether uncertainty is estimated for the first distribution parameter (TRUE) or not (FALSE) |
| Second distribution parameter type | distribution_par2_type | character | Type of value for the second distribution parameter - see dropdown list |
| Second distribution parameter value | distribution_par2_value | numeric | Value for the second distribution parameter (e.g., shape or scale parameter for a gamma distribution) |
| Second distribution parameter uncertainty | distribution_par2_uncertainty | logical | Tick box for whether uncertainty is estimated for the second distribution parameter (TRUE) or not (FALSE) |
| Disaggregated data available | method_disaggregated | logical | Tick box if disaggregated estimates are available (TRUE) or not (FALSE) |
| Parameter estimates disaggregated by | method_disaggregated_by | character | Categories for disaggregation of parameter estimates |
| Only disaggregated data available | method_disaggregated_only | logical | Tick box if ONLY disaggregated estimates are available (TRUE) or if a central estimate is also available (FALSE) |
| Is parameter from supplement? | method_from_supplement | logical | Tick box for whether the parameter was extracted from a supplement (TRUE) or not (FALSE) |
| Is parameter in a figure only? | parameter_fromfigure | logical | Tick box to indicate that the parameter is plotted in a figure but not reported numerically in the text/table, and thus could not be extracted |
| Study population country | population_country | character | Country of the survey population - see dropdown list |
| Study population location | population_location | character | Region/district/province/city of the survey population - see dropdown list |
| Start day of study | population_study_start_day | integer | Study start day |
| Start month of study | population_study_start_month | character | Study start month - see dropdown list |
| Start year of study | population_study_start_year | integer | Study start year - see dropdown list |
| End day of study | population_study_end_day | integer | Study end day |
| End month of study | population_study_end_month | character | Study end month - see dropdown list |
| End year of study | population_study_end_year | integer | Study end year - see dropdown list |
| Survey timing related to outbreak | method_moment_value | character | Timing of the survey in relation to the outbreak, if specified in paper - see dropdown list |
| Study population sample size | population_sample_size | integer | Sample size of the population used for parameter estimation |
| Study population minimum age (years) | population_age_min | numeric | Minimum age of the survey population in years |
| Study population maximum age (years) | population_age_max | numeric | Maximum age of the survey population in years |
| Sex of study population | population_sex | character | Sex of survey population - see dropdown list |
| Population sample setting | population_sample_type | character | General setting of the survey - see dropdown list |

| | | | |
|---|---|---|---|
| Population group | population_group | character | Specific group of the survey population - see dropdown list |
| Genome site | genome_site | character | Site of genome or gene studied |
| Genomic sequence available? | genomic_sequence_available | logical | Tick box whether genomic sequence data are available (TRUE) or not (FALSE) |
| Reproduction number pathway | r_pathway | character | Transmission pathway that reproduction number is based on for vector-borne diseases |
| Method to estimate R | method_r | character | Method used for estimation of the reproduction number - see dropdown list |
| Other delay start point | other_delay_start | character | Start point for delays not in the parameter type dropdown list (e.g., delay from ... to ...) |
| Other delay end point | other_delay_end | character | End point for delays not in the parameter type dropdown list (e.g., delay from ... to ...) |
| Numerator | cfr_ifr_numerator | integer | Numerator of either CFR/IFR (deaths) or seroprevalence (number seropositive) estimates |
| Denominator | cfr_ifr_denominator | integer | Denominator of either CFR/IFR (cases) or seroprevalence (number tested) estimates |
| Is the CFR/IFR estimate adjusted? | cfr_ifr_method | character | Is the CFR/IFR estimate adjusted, unadjusted, or unspecified - see dropdown list |
| Outcome for risk factor(s) | riskfactor_outcome | character | Outcome for risk factor(s) - see dropdown list |
| Risk factor name | riskfactor_name | character | Risk factor name - see dropdown list |
| Risk factor occupation | riskfactor_occupation | character | If risk factor is an occupation, then specified occupation as risk factor - see dropdown list |
| Risk factor adjusted | riskfactor_adjusted | character | Adjustment status of risk factor(s)- see dropdown list |
| Risk factor significant | riskfactor_significant | character | Statistical significance of risk factor(s) - see dropdown list |
| Parameter class | parameter_class | character | General parameter class (delays, seroprevalence, reproduction numbers, mutations, severity, risk factors, relative contribution) |

Table B.5: Parameter data fields: refer to epireview in Supplement D for dropdown options.

| Location | Outbreak Dates | Suspected Cases | Confirmed Cases | Confirmation Method | Severe Cases | Deaths | Source |
|---|---|---|---|---|---|---|---|
| **Benin** | | | | | | | |
| Borgou, Donga, Collines, Alibori, Plateau, Ouémé, Atlantique, Litoral | Dec 2015 - Mar 2016 | 3 | 18 | Molecular | | 11 | Yadouleton (2020) |
| Northern Benin | Oct 2014 - Nov 2014 | 11 | 2 | Molecular | | 11 | Yadouleton (2020) |
| **Liberia** | | | | | | | |
| Bong, Nimba, Montserrado | Jan 2016 - Jun 2016 | 14 | 14 | Molecular | | 19 | Hamblion (2018) |
| Curran Lutheran Hospital, Zorzor | Jul 1980 - Apr 1982 | | 14 | Molecular | | | Frame (1984b) |
| Zorzor | Mar 1972 - Apr 1972 | | 11 | Unspecified | | | Lo Iacono (2015) |
| Zorzor | Mar 1972 - Apr 1972 | | 11 | Unspecified | | 4 | Mertens (1973) |
| **Nigeria** | | | | | | | |
| Edo, Ondo, Ebonyi | 1 Jan 2019 - 29 Dec 2019 | 5057 | 833 | Molecular | | 174 | Nwafor (2021) |
| | 2018 - 2019 | | 1463 | Molecular | | 344 | Olayinka (2022) |
| | 1 Jan 2018 - 6 May 2018 | 10 | 423 | Molecular | | 106 | Ilori (2019b) |
| | 1 Jan 2018 - 29 Apr 2018 | | 420 | Unspecified | | 106 | Tuite (2019) |
| | 1 Jan 2018 - 25 Mar 2018 | | 394 | Unspecified | | 104 | Ilori (2019a) |
| Anambra, Bauchi, Benue, Delta, Ebonyi, Edo, Ekiti, FCT, Abuja, Gombe, Imo, Kaduna, Kogi, Lagos, Nasarawa, Ondo, Osun, Plateau, Rivers, Taraba | 1 Jan 2018 - 18 Mar 2018 | 1495 | 376 | Unspecified | | 95 | Loyinmi (2021) |
| | Dec 2017 - May 2018 | | 431 | Molecular | | | Siddle (2018) |
| | 2017 | | 312 | Unspecified | | 78 | Joseph (2019) |
| | 1 Jan 2017 - 25 Mar 2017 | | 107 | Unspecified | | 50 | Ilori (2019a) |
| Ondo State | 1 Dec 2015 - 30 Apr 2016 | | 19 | Molecular | | 12 | Isere (2018) |
| Taraba, Edo, Lagos, Bauchi, Rivers, Gombe, Ekiti | Oct 2015 - Feb 2016 | | 47 | Molecular | | 28 | Buba (2018) |
| Ebonyi State | 1 Jan 2012 - 25 Mar 2012 | 10 | 10 | Molecular | | 6 | Ajayi (2013) |
| Onitsha | Jan 1974 - Feb 1974 | | 3 | Molecular | | 1 | Bowen (1975) |
| Jos Plateau | Jan 1970 - | 26 | 13 | Unspecified | | 10 | Troup (1970) |
| Jos | Jan 1970 - Feb 1970 | 13 | 15 | Molecular | | 13 | Carey (1972) |
| Jos | 25 Dec 1969 - | | 23 | Unspecified | | | Lo Iacono (2015) |
| Lassa, Jos | 12 Jan 1969 - 3 May 1969 | 3 | | Unspecified | | 2 | Frame (1970) |
| **Sierra Leone** | | | | | | | |
| Tonkolili District | 30 Oct 2019 - 16 Nov 2019 | 2 | 3 | Molecular | | 3 | Njuguna (2022) |
| Kenema Government Hospital | 27 Apr 2010 - 31 Jan 2012 | 1002 | | Unspecified | | | Lo Iacono (2016) |
| Kenema Government Hospital | 27 Apr 2010 - 31 Jan 2012 | 1002 | 295 | Molecular | | | Lo Iacono (2015) |
| Kenema, Kailahun | Jan 1997 - Apr 1997 | | | Molecular | 352 | | Allan (1999) |
| Panguma, Tongo | 23 Sep 1972 - 15 Oct 1972 | 20 | 12 | Molecular | | | Fraser (1974) |

Table B.6: Characteristics of extracted ‘outbreaks’ reported by study authors.

| Transmission Route(s) | Human Transmission Heterogeneity | Human Compartments | Calibration | Interventions | Source |
|---|---|---|---|---|---|
| **Branching Process** | | | | | |
| Rodent-Human, Human-Human | | | Fitted | | Lo Iacono (2015) |
| Rodent-Human, Human-Human | Spatial, Time | | Fitted | | Akhmetzhanov (2019) |
| Rodent-Human, Human-Human | Time | | Fitted | | Lo Iacono (2016) |
| **Compartmental - Deterministic** | | | | | |
| Rodent-Human | | Other | Theoretical | Treatment, Rodent Control | Faniran (2022) |
| Human-Human | | Other | Theoretical | Treatment | Atangana (2015) |
| Human-Human | | Other | Theoretical | | Chu (2023) |
| Human-Human | | Other | Theoretical | | Goyal (2019) |
| Human-Human | | Other | Theoretical | | Higazy (2021) |
| Human-Human | | Other | Theoretical | | Gao (2020) |
| Rodent-Human, Human-Human | | SIR | Theoretical | | Alsallami (2022) |
| Rodent-Human, Human-Human | | SIR | Theoretical | Rodent Control | Abdullahi (2021) |
| Rodent-Human, Human-Human | | SEIR | Theoretical | Vaccination | Davies (2019) |
| Rodent-Human, Human-Human | | SEIR | Theoretical | Behaviour Changes, Other, Quarantine, Treatment, Rodent Control | Bakare (2020) |
| Rodent-Human, Human-Human | | SEIR | Theoretical | | Shoaib (2022) |
| Rodent-Human, Human-Human | | SEIR | Theoretical | | Loyinmi (2021) |
| Rodent-Human, Human-Human | | SEIR | Theoretical | | Alkahtani (2020) |
| Rodent-Human, Human-Human | | Other | Fitted | Behaviour Changes, Treatment | Musa (2022) |
| Rodent-Human, Human-Human | | Other | Fitted | Hospitals, Quarantine | Musa (2020) |
| Rodent-Human, Human-Human | | Other | Fitted | Quarantine | Abdulhamid (2022) |
| Rodent-Human, Human-Human | | Other | Fitted | Quarantine, Treatment, Rodent Control | Collins (2023) |
| Rodent-Human, Human-Human | | Other | Fitted | Behaviour Changes, Rodent Control | Ndenda (2022) |
| Rodent-Human, Human-Human | Age | SEIR | Theoretical | | Onifade (2021) |
| Rodent-Human, Human-Human | Community Hygiene | Other | Theoretical | Behaviour Changes, Treatment, Rodent Control | Abidemi (2022) |
| Rodent-Human, Human-Human | Community Hygiene | Other | Theoretical | Behaviour Changes, Other, Treatment, Rodent Control | Abidemi (2023) |
| Rodent-Human, Human-Human | Protective Behaviour | Other, SIR | Theoretical | Other, Quarantine, Treatment, Rodent Control | Onah (2020b) |
| Rodent-Human, Human-Human | Quarantine Status, Time | Other | Fitted | Behaviour Changes, Quarantine, Treatment, Rodent Control | Barua (2021) |
| Rodent-Human, Human-Human | Socio-Economic Status | SIR | Theoretical | | Onah (2020a) |
| Rodent-Human, Human-Human | Socio-Economic Status | Other | Fitted | Contact Tracing, Hospitals, Quarantine | Ogunmiloro (2023) |
| Rodent-Human, Human-Human | Time | Other | Fitted | Treatment | Ibrahim (2021) |
| Rodent-Human, Human-Human, Airborne | | SEIR | Theoretical | | Madueme (2023) |
| **Compartmental - Stochastic** | | | | | |
| Rodent-Human, Human-Human | | SIR | Theoretical | Behaviour Changes | Hamam (2022) |
| **Other - Deterministic** | | | | | |
| Rodent-Human | Spatial | | Fitted | | Basinski (2021) |
| **Other - Stochastic** | | | | | |
| Rodent-Human | Spatial | | Fitted | | Redding (2016) |

Table B.7: Characteristics of extracted transmission models.

| Parameter | Uncertainty | Disaggregated By | Gene | Method | Sample Size | Country | Study Dates | Study Setting | Study Group | Source |
|---|---|---|---|---|---|---|---|---|---|---|
| **Reproduction Number** | | | | | | | | | | |
| 0.73 | Maximum: 11.7 | | | Renewal Equations / Branching Process | | Liberia, Nigeria | 26 Dec 1969 - Apr 1972 | Hospital | Persons Under Investigation | Lo Iacono (2015) |
| 1.1299 | | | | Next Generation Matrix | | Nigeria | 3 Jan 2021 - 19 May 2021 | Population | Persons Under Investigation | Ndenda (2022) |
| 1.1868 | | | | Next Generation Matrix | | Nigeria | 2018 - 2022 | Population | Persons Under Investigation | Collins (2023) |
| 1.05 - 1.62 | | Method, Other, Region, Time | | Growth Rate | | Nigeria | Jan 2016 - Mar 2019 | Population | General Population | Zhao (2020) |
| **Growth Rate** | | | | | | | | | | |
| 0.32 per day | CI95%: 0.23-0.64 | | | | | Nigeria | 29 Oct 2018 - 21 Jan 2019 | Population | Persons Under Investigation | Musa (2020) |
| 0.43 per day | CI95%: 0.23-0.72 | | | | | Nigeria | 30 Oct 2017 - 22 Jan 2018 | Population | Persons Under Investigation | Musa (2020) |
| 0.51 per day | CI95%: 0.17-1.06 | | | | | Nigeria | 31 Oct 2016 - 23 Jan 2017 | Population | Persons Under Investigation | Musa (2020) |
| **Attack Rate** | | | | | | | | | | |
| 2.2 per 1000 | | Age, Sex | | | | Sierra Leone | 1 Oct 1970 - 1 Oct 1972 | Population | General Population | Fraser (1974) |
| 3.1 per 1000 | CI95%: 2.5-3.8 | Age, Sex | | | 28882 | Sierra Leone | Sep 2002 - Jul 2004 | Household | Other | Bonner (2007) |
| 0.56 % | CI95%: 0.35-0.77 | | | | 5001 | Nigeria | 1 Jan 2018 - 6 May 2018 | Contact | Persons Under Investigation | Ilori (2019b) |
| **Human-Human Transmission Contribution** | | | | | | | | | | |
| 20 % | IQR: 15-30 | | | | | Sierra Leone | 27 Apr 2010 - 31 Jan 2012 | Hospital | Persons Under Investigation | Lo Iacono (2015) |
| 22 % | | | | | 1002 | Sierra Leone | 27 Apr 2010 - 31 Jan 2012 | Hospital | Persons Under Investigation | Lo Iacono (2016) |
| 18.5 - 26 % | | Method | | | | Sierra Leone | 27 Apr 2010 - 31 Jan 2012 | Hospital | Persons Under Investigation | Lo Iacono (2015) |
| **Evolutionary Rate** | | | | | | | | | | |
| 7.7 s/s/y $10^{-4}$ | CrI95%: 6.2-9.3 | | GPC | | 11 | Nigeria | 2012 - 2016 | Hospital | Persons Under Investigation | Oloniniyi (2018) |
| 8.4 s/s/y $10^{-4}$ | CrI95%: 7.2-9.6 | | L | | 11 | Nigeria | 2012 - 2016 | Hospital | Persons Under Investigation | Oloniniyi (2018) |
| 10.1 s/s/y $10^{-4}$ | CrI95%: 8.2-12.2 | | NP | | 11 | Nigeria | 2012 - 2016 | Hospital | Persons Under Investigation | Oloniniyi (2018) |
| 16.9 s/s/y $10^{-4}$ | CrI95%: 8.6-25.9 | | Z | | 11 | Nigeria | 2012 - 2016 | Hospital | Persons Under Investigation | Oloniniyi (2018) |
| **Substitution Rate** | | | | | | | | | | |
| 7.8 s/s/y $10^{-4}$ | CrI95%: 5.4-10.2 | | L segment | | 219 | Nigeria | 1969 - 2018 | | | Ehichioya (2019) |
| 8.4 s/s/y $10^{-4}$ | CrI95%: 7.6-9.2 | | L segment | | | | | | | Wiley (2019) |
| 7.8 s/s/y $10^{-4}$ | CrI95%: 6.3-9.1 | | S segment | | 219 | Nigeria | 1969 - 2018 | | | Ehichioya (2019) |
| 8.2 s/s/y $10^{-4}$ | CrI95%: 7.2-9.1 | | S segment | | | | | | | Wiley (2019) |

Table B.8: Transmission parameters grouped by parameter type. Study characteristics are reported for each parameter estimate. Note that s/s/y stands for nucleotide substitutions per site per year.

| Delay | Statistic | Uncertainty | Disaggregated By | Sample Size | Country | Study Dates | Study Setting | Study Group | Source |
|---|---|---|---|---|---|---|---|---|---|
| **Incubation Period** | | | | | | | | | |
| 7 days | Unspecified | | | 62 | Nigeria | 1 Jan 2018 - 31 Mar 2018 | Population | Persons Under Investigation | Joseph (2019) |
| 10 days | Mean | Range: 5-18 | | 20 | Nigeria | 1 Jan 2012 - 25 Mar 2012 | Hospital | Persons Under Investigation | Ajayi (2013) |
| 7 - 17 days | | | | 3 | Nigeria | 12 Jan 1969 - 20 Feb 1969 | Hospital | Healthcare Workers | Frame (1970) |
| 1 - 24 days | | | | 11 | Liberia | Mar 1972 - Apr 1972 | Hospital | Persons Under Investigation | Mertens (1973) |
| 12.8 days | Mean | Gamma SD: 4.6 | | | Nigeria | 2016 - 2018 | Hospital | Persons Under Investigation | Akhmetzhanov (2019) |
| **Onset - Testing** | | | | | | | | | |
| 6.5 days | Median | Range: 0-31 | | 14 | Liberia | Jan 2016 - Jun 2016 | Population | Persons Under Investigation | Hamblion (2018) |
| 7 days | Median | Range: 0-19 | Other | 21 | Benin | Dec 2015 - Mar 2016 | Hospital | Persons Under Investigation | Yadouleton (2020) |
| **Testing - Test Result** | | | | | | | | | |
| 6 days | Median | Range: 2-64 | | 14 | Liberia | Jan 2016 - Jun 2016 | Population | Persons Under Investigation | Hamblion (2018) |
| **Onset - Admission** | | | | | | | | | |
| 2 days | Median | | | 19 | Nigeria | 1 Dec 2015 - 30 Apr 2016 | Hospital | Persons Under Investigation | Isere (2018) |
| 4 days | Mean | Range: 0-7 | | 11 | Liberia | Mar 1972 - Apr 1972 | Hospital | Persons Under Investigation | Mertens (1973) |
| 4.3 days | Mean | Range: 1-14 | | 12 | Guinea | | Hospital | Persons Under Investigation | ter Meulen (1998) |
| 4.9 days | Mean | Range: 1-12 | | 23 | Nigeria | Jan 1970 - Feb 1970 | Hospital | Persons Under Investigation | White (1972) |
| 6 days | Mean | SD: 2.6 | | 14 | Nigeria | 1 Jan 2012 - 25 Mar 2012 | Hospital | Other | Ajayi (2013) |
| 6 days | Mean | SD: 3.1 | | 19 | Nigeria | 2009 - 2018 | Hospital | Other | Okogbenin (2019) |
| 6 days | Median | IQR: 4-10 | Age, Disease Generation, Occupation, Other, Region, Sex | 389 | Nigeria | Dec 2018 - Apr 2019 | Population | Persons Under Investigation | Chandra (2021) |
| 4 - 9 days | | | | 12 | Sierra Leone | 23 Sep 1972 - 15 Oct 1972 | Hospital | Persons Under Investigation | Monath (1974) |
| 6.5 days | Median | IQR: 4-10 | Other | 174 | Nigeria | Jan 2009 - Dec 2010 | Hospital | Persons Under Investigation | Asogun (2012) |
| 7 days | Median | IQR: 5-9 | | 71 | Sierra Leone | 3 Jun 2019 - 28 Sep 2019 | Hospital | Other | Akpogheneta (2021) |
| 7 days | Median | IQR: 4-13 | Method | 67 | Nigeria | Dec 2017 - Apr 2018 | Hospital | Persons Under Investigation | Chika-Igwenyi (2021) |
| 7.5 days | Mean | SD: 4.9 | | 103 | Liberia | Jan 2019 - Dec 2020 | Hospital | Persons Under Investigation | Jetoh (2022) |
| 8 days | Median | IQR: 7-13 | | 510 | Nigeria | 5 Apr 2018 - 15 Mar 2020 | Hospital | Persons Under Investigation | Duvignaud (2021) |
| 8.7 days | Mean | SE: 0.6 | | 311 | Guinea | Mar 1996 - Dec 1999 | Hospital | Persons Under Investigation | Bausch (2001) |
| 9.3 days | Mean | | | 107 | Nigeria, Sierra Leone | 2008 - 2013 | Hospital | Persons Under Investigation | Andersen (2015) |
| 10 days | Mean | SD: 3.5 | | 11 | Nigeria | 2009 - 2018 | Hospital | Other | Okogbenin (2019) |
| 10 days | Median | IQR: 7-12 | Method | 9 | Nigeria | Aug 2018 - Dec 2018 | Hospital | Persons Under Investigation | Chika-Igwenyi (2021) |
| 11 days | Mean | SD: 3.5 | | 6 | Nigeria | 1 Jan 2012 - 25 Mar 2012 | Hospital | Other | Ajayi (2013) |
| **Onset - Start of Treatment** | | | | | | | | | |
| 8 days | Median | IQR: 7-13 | | 510 | Nigeria | 5 Apr 2018 - 15 Mar 2020 | Hospital | Persons Under Investigation | Duvignaud (2021) |
| **Duration of Antiviral Treatment** | | | | | | | | | |
| 10 days | Median | IQR: 9-13 | | 510 | Nigeria | 5 Apr 2018 - 15 Mar 2020 | Hospital | Persons Under Investigation | Duvignaud (2021) |
| **Duration of Antibacterial Therapy** | | | | | | | | | |
| 8 days | Median | IQR: 7-10 | | 459 | Nigeria | 5 Apr 2018 - 15 Mar 2020 | Hospital | Persons Under Investigation | Duvignaud (2021) |
| **Duration of Oxygen Therapy** | | | | | | | | | |
| 3 days | Median | IQR: 2-7 | | 92 | Nigeria | 5 Apr 2018 - 15 Mar 2020 | Hospital | Persons Under Investigation | Duvignaud (2021) |
| **Admission - Symptom Resolution** | | | | | | | | | |
| 8.2 days | Mean | SD: 3.8 | | 14 | Nigeria | 1 Jan 2012 - 25 Mar 2012 | Hospital | Other | Ajayi (2013) |
| **Symptomatic Period** | | | | | | | | | |
| 1 - 4 weeks | | | | | Nigeria | Jan 1970 - Feb 1970 | Hospital | Persons Under Investigation | Troup (1970) |
| **Admission - Recovery** | | | | | | | | | |

continued on next page

continued from previous page

| Delay | Statistic | Uncertainty | Disaggregated By | Sample Size | Country | Study Dates | Study Setting | Study Group | Source |
|---|---|---|---|---|---|---|---|---|---|
| 10 days | Median | IQR: 8-13 | | 101 | Nigeria | Jan 2009 - Dec 2010 | Hospital | Persons Under Investigation | Asogun (2012) |
| 10 days | Median | Range: 5-36 | | 14 | Nigeria | 1 Jan 2012 - 25 Mar 2012 | Hospital | Other | Ajayi (2013) |
| 11 days | Median | IQR: 10-15 | | 449 | Nigeria | 5 Apr 2018 - 15 Mar 2020 | Hospital | Persons Under Investigation | Duvignaud (2021) |
| **Admission - Death** | | | | | | | | | |
| 2 days | Median | IQR: 1-4 | | 57 | Nigeria | Jan 2009 - Dec 2010 | Hospital | Persons Under Investigation | Asogun (2012) |
| 3 days | Median | IQR: 1-6 | | 61 | Nigeria | 5 Apr 2018 - 15 Mar 2020 | Hospital | Persons Under Investigation | Duvignaud (2021) |
| 3.3 days | Mean | SE: 1.2 | | 22 | Guinea | Mar 1996 - Dec 1999 | Hospital | Persons Under Investigation | Bausch (2001) |
| 7 days | Median | Range: 0-12 | | 6 | Nigeria | 1 Jan 2012 - 25 Mar 2012 | Hospital | Other | Ajayi (2013) |
| **Time In Care** | | | | | | | | | |
| 11 days | Median | IQR: 5-14 | | 20 | Sierra Leone | 1 Nov 2011 - 31 Oct 2012 | Hospital | Persons Under Investigation | Roth (2015) |
| 11.5 days | Mean | SD: 4.97 | | | Nigeria | 1 Jan 1989 - 31 Mar 1989 | Hospital | Persons Under Investigation | Fisher-Hoch (1995) |
| **Onset - Recovery** | | | | | | | | | |
| 14 days | Mean | Range: 11-21 | | 11 | Liberia | Mar 1972 - Apr 1972 | Hospital | Persons Under Investigation | Mertens (1973) |
| 16 days | Median | IQR: 13-23 | | 98 | Nigeria | Jan 2009 - Dec 2010 | Hospital | Persons Under Investigation | Asogun (2012) |
| 17.3 days | Mean | Range: 7-31 | | 23 | Nigeria | Jan 1970 - Feb 1970 | Hospital | Persons Under Investigation | White (1972) |
| **Onset - Death** | | | | | | | | | |
| 10 days | Median | IQR: 7-14 | | 52 | Nigeria | Jan 2009 - Dec 2010 | Hospital | Persons Under Investigation | Asogun (2012) |
| 1 - 2 weeks | | | | | Nigeria | Jan 1970 - Feb 1970 | Hospital | Persons Under Investigation | Troup (1970) |
| 11.6 days | Mean | Range: 7-26 | | 23 | Nigeria | Jan 1970 - Feb 1970 | Hospital | Persons Under Investigation | White (1972) |
| 12 days | Mean | Range: 12-15 | | 11 | Liberia | Mar 1972 - Apr 1972 | Hospital | Persons Under Investigation | Mertens (1973) |
| 13.8 days | Mean | Gamma SD: 7.6 | | | Nigeria | 2016 - 2018 | Hospital | Persons Under Investigation | Akhmetzhanov (2019) |
| 7 - 21 days | | | | | Guinea, Liberia, Sierra Leone | Jul 1978 - Apr 1979 | Hospital | Persons Under Investigation | Knobloch (1980) |

Table B.9: Delay parameters grouped by parameter type. Study characteristics are reported for each parameter estimate.

| CFR (%) | Uncertainty | Disaggregated By | Method | Number of Deaths | Sample Size | Study Dates | Study Setting | Study Group | Source |
|---|---|---|---|---|---|---|---|---|---|
| **Benin** | | | | | | | | | |
| 44 | | Other | Adjusted | 8 | 18 | Dec 2015 - Mar 2016 | Hospital | Persons Under Investigation | Yadouleton (2020) |
| **Guinea** | | | | | | | | | |
| 18 | | | Unspecified | 4 | 22 | Mar 1996 - Dec 1999 | Hospital | Persons Under Investigation | Bausch (2001) |
| 33.3 | | | Naive | 4 | 12 | | Hospital | Persons Under Investigation | ter Meulen (1998) |
| **Guinea, Liberia, Sierra Leone** | | | | | | | | | |
| 14 | | | Unspecified | | 42 | Jul 1978 - Apr 1979 | Hospital | Persons Under Investigation | Knobloch (1980) |
| **Liberia** | | | | | | | | | |
| 9.3 | | Age, Other, Sex, Symptoms | Adjusted | 23 | 246 | 1 Jul 1980 - 30 Apr 1986 | Hospital | Persons Under Investigation | Frame (1989) |
| 13.6 | | Age, Other, Sex | Unspecified | 6 | 44 | Jul 1980 - Apr 1982 | Hospital | Persons Under Investigation | Monson (1984) |
| 15.6 | | | Adjusted | 5 | 32 | 1 Jul 1980 - 30 Apr 1986 | Hospital | Pregnant Women | Frame (1989) |
| 17 | | | Unspecified | 3 | 18 | | Hospital | Pregnant Women | Monson (1987) |
| 26.7 | | | Adjusted | 4 | 15 | 1 Jul 1980 - 30 Apr 1986 | Hospital | Children | Frame (1989) |
| 27 | | | Unspecified | 4 | 15 | | Hospital | Children | Monson (1987) |
| 29 | | Region | Unspecified | 4 | 14 | Jan 2016 - Jun 2016 | Population | Persons Under Investigation | Hamblion (2018) |
| 36 | | | Unspecified | 4 | 11 | Mar 1972 - Apr 1972 | Hospital | Persons Under Investigation | Monath (1973) |
| 36 | | | Naive | 4 | 11 | Mar 1972 - Apr 1972 | Hospital | Persons Under Investigation | Mertens (1973) |
| 39.8 | | Time | Unspecified | 41 | 103 | Jan 2019 - Dec 2020 | Hospital | Persons Under Investigation | Jetoh (2022) |
| 66.7 | | | Unspecified | 2 | 3 | Jul 1980 - Apr 1982 | Hospital | Pregnant Women | Monson (1984) |
| 100 | | | Unspecified | 2 | 2 | Jul 1980 - Apr 1982 | Hospital | Children | Monson (1984) |
| **Nigeria** | | | | | | | | | |
| 4.9 | CrI95%: 0-5.4 | | Unspecified | | | 2016 - 2018 | Hospital | Persons Under Investigation | Akhmetzhanov (2019) |
| 12 | | Age, Other, Sex | Adjusted | 62 | 510 | 5 Apr 2018 - 15 Mar 2020 | Hospital | Persons Under Investigation | Duvignaud (2021) |
| 13.8 | | | Adjusted | 100 | 724 | Jan 2018 - Jun 2019 | Population | Persons Under Investigation | Olayinka (2022) |
| 17.4 | | Age, Occupation, Region, Sex, Time | Naive | 550 | 3162 | Jan 2018 - Dec 2021 | Hospital | Persons Under Investigation | Dalhat (2022) |
| 18.5 | | Region, Time | Naive | 516 | 2787 | Dec 2016 - Sep 2020 | Hospital | Persons Under Investigation | Yaro (2021) |
| 19 | | Occupation, Other | Unspecified | 12 | 62 | 1 Jan 2018 - 31 Mar 2018 | Population | Persons Under Investigation | Joseph (2019) |
| 9.3 - 29.2 | | Time | Unspecified | | | Jan 2015 - 26 Sep 2021 | Population | Persons Under Investigation | Grace (2021) |
| 21 | | | Unspecified | 174 | 833 | 1 Jan 2019 - 29 Dec 2019 | Population | Mixed Groups | Nwafor (2021) |
| | | | Naive | 3 | 13 | 1 Dec 2009 - 30 Nov 2010 | Hospital | Children | Akhuemokhan (2017) |
| 24 | | Age, Region | Adjusted | 68 | 284 | Jan 2011 - Nov 2015 | Hospital | Persons Under Investigation | Okokhere (2018) |
| 25.1 | CI95%: 20.9-29.2 | Region | Unspecified | 106 | 423 | 1 Jan 2018 - 6 May 2018 | Hospital | Persons Under Investigation | Ilori (2019b) |
| 25.8 | | | Unspecified | | | 1 Jan 2018 - 25 Mar 2018 | Population | Persons Under Investigation | Ilori (2019a) |
| 27.9 | | Time | Naive | 362 | 1298 | 2008 - 2018 | Hospital | Persons Under Investigation | Akpede (2019) |
| 28 | | Sex | Unspecified | 18 | 64 | Jan 2007 - Dec 2007 | Hospital | Persons Under Investigation | Inegbenebor (2010) |
| 28 | | Age, Disease Generation, Symptoms | Unspecified | 16 | 57 | Jan 2009 - Aug 2017 | Hospital | Children | Adetunji (2021) |
| 29 | | | Unspecified | | | Nov 2005 - Feb 2008 | Hospital | Persons Under Investigation | Ehichioya (2012) |
| 29.9 | | | Adjusted | 20 | 67 | Dec 2017 - Apr 2018 | Hospital | Persons Under Investigation | Chika-Igwenyi (2021) |
| 30 | | Other | Naive | 6 | 20 | 1 Jan 2012 - 25 Mar 2012 | Hospital | Persons Under Investigation | Ajayi (2013) |
| 31 | | | Naive | 61 | 198 | Jan 2009 - Dec 2010 | Hospital | Persons Under Investigation | Asogun (2012) |
| 31.6 | | Occupation, Sex | Adjusted | 5 | 16 | 28 Dec 2017 - 2018 | Hospital | Healthcare Workers | Dan-Nwafor (2019) |
| 36 | | | Unspecified | | 11 | Jan 2016 - Aug 2016 | Hospital | Persons Under Investigation | Shehu (2018) |
| 36 | | | Adjusted | 61 | 170 | Jan 2009 - Dec 2010 | Hospital | Persons Under Investigation | Asogun (2012) |
| 36.7 | | | Unspecified | 11 | 30 | 2009 - 2018 | Hospital | Pregnant Women | Okogbenin (2019) |
| 37.2 | | Age, Sex | Unspecified | 42 | 113 | 1 Jan 2018 - 31 Mar 2019 | Hospital | Persons Under Investigation | Nwafor (2020) |
| 37.6 | | Other | Unspecified | 115 | 306 | Jan 2014 - Apr 2017 | Hospital | Persons Under Investigation | Strampe (2021) |
| 43.9 | | | Unspecified | | | 1 Jan 2017 - 25 Mar 2017 | Population | Persons Under Investigation | Ilori (2019a) |

continued on next page

continued from previous page

| CFR (%) | Uncertainty | Disaggregated By | Method | Number of Deaths | Sample Size | Study Dates | Study Setting | Study Group | Source |
|---|---|---|---|---|---|---|---|---|---|
| 52 | | | Naive | 12 | 23 | Jan 1970 - Feb 1970 | Hospital | Persons Under Investigation | White (1972) |
| 54 | | Age, Time | Unspecified | 41 | 76 | Jan 2015 - Dec 2018 | Hospital | Persons Under Investigation | Abdulkarim (2020) |
| 57 | | | Adjusted | 25 | 44 | 2016 - 2018 | Hospital | Persons Under Investigation | Gomerep (2022) |
| 59.6 | | Age, Other, Region, Sex, Time | Adjusted | 28 | 47 | Oct 2015 - Feb 2016 | Hospital | Persons Under Investigation | Buba (2018) |
| 60 | | | Unspecified | 24 | 40 | 2008 - 2013 | Hospital | Persons Under Investigation | Andersen (2015) |
| 63.2 | | | Unspecified | 12 | 19 | 1 Dec 2015 - 30 Apr 2016 | Hospital | Persons Under Investigation | Isere (2018) |
| 65 | | | Unspecified | 22 | 34 | 1 Jan 1989 - 31 Mar 1989 | Hospital | Persons Under Investigation | Fisher-Hoch (1995) |
| 85.7 | | | Adjusted | 12 | 14 | Aug 2018 - Dec 2018 | Hospital | Persons Under Investigation | Chika-Igwenyi (2021) |
| **Sierra Leone** | | | | | | | | | |
| 8 | | | Unspecified | 1 | 12 | 23 Sep 1972 - 15 Oct 1972 | Hospital | Persons Under Investigation | Monath (1974) |
| 8.8 | | | Naive | 8 | 90 | Sep 2002 - Jul 2004 | Hospital | Persons Under Investigation | Bonner (2007) |
| | | Age, Sex | Naive | 6 | 51 | Aug 1977 - Aug 1981 | Hospital | Children | Webb (1986) |
| 13 | | | Adjusted | 10 | 79 | 1981 - 1985 | Hospital | Other | Price (1988) |
| 15.6 | | Other | Naive | 53 | 333 | 1977 - | Hospital | Persons Under Investigation | McCormick (1986) |
| 15.9 | | | Naive | 22 | 138 | Jan 1973 - Mar 1976 | Hospital | Persons Under Investigation | Keane (1977) |
| 16.5 | | Age, Sex | Unspecified | 73 | 441 | Feb 1977 - Jan 1979 | Hospital | Persons Under Investigation | McCormick (1987a) |
| 16.6 | | | Naive | 16 | 96 | 1975 | Hospital | Persons Under Investigation | Keane (1977) |
| 21 | | Other | Adjusted | 14 | 68 | 1981 - 1985 | Hospital | Pregnant Women | Price (1988) |
| 33 | | | Naive | 6 | 18 | Jan 1973 - Mar 1976 | Hospital | Pregnant Women | Keane (1977) |
| 14.8 - 56 | | Time | Unspecified | | | Jan 1996 - Dec 1997 | Hospital | Persons Under Investigation | Allan (1999) |
| 38 | | Age, Sex | Unspecified | 24 | 63 | 1 Oct 1970 - 1 Oct 1972 | Hospital | Persons Under Investigation | Fraser (1974) |
| 38.1 | | | Adjusted | 8 | 21 | 1 Nov 2011 - 31 Oct 2012 | Hospital | Persons Under Investigation | Roth (2015) |
| 60 | | | Unspecified | 3 | 5 | 30 Oct 2019 - 16 Nov 2019 | Hospital | Persons Under Investigation | Njuguna (2022) |
| 61 | | Age, Other, Sex, Symptoms | Unspecified | 22 | 36 | Apr 2011 - Feb 2012 | Hospital | Mixed Groups | Dahmane (2014) |
| 63 | | | Unspecified | 36 | 57 | 1 Jan 2012 - 31 Dec 2018 | Hospital | Children | Samuels (2021) |
| | | | Unspecified | 6 | 9 | 1 Oct 1970 - 1 Oct 1972 | Hospital | Pregnant Women | Fraser (1974) |
| 67.3 | | Method, Time | Adjusted | 165 | 245 | 2008 - 2016 | Hospital | Persons Under Investigation | Shaffer (2019) |
| 69 | | Method | Adjusted | 109 | 158 | 2008 - 2012 | Hospital | Persons Under Investigation | Shaffer (2014) |
| 75 | | | Naive | 9 | 12 | 1975 | Hospital | Pregnant Women | Keane (1977) |
| 78.5 | | Other, Time | Adjusted | 106 | 135 | 1 Jan 2012 - 31 Dec 2019 | Hospital | Persons Under Investigation | Shaffer (2021) |
| 81 | | | Unspecified | 54 | 67 | 2008 - 2013 | Hospital | Persons Under Investigation | Andersen (2015) |

Table B.10: Case fatality ratios (CFRs) grouped by country. Study characteristics are reported for each CFR estimate.

| Seroprevalence (%) | Uncertainty | Disaggregated By | Assay | Number Seropositive | Sample Size | Study Dates | Study Setting | Study Group | Source |
|---|---|---|---|---|---|---|---|---|---|
| **Benin** | | | | | | | | | |
| 9.9 | | | IgG | | | | Population | General Population | Emmerich (2008) |
| **Botswana** | | | | | | | | | |
| | | Time, Other | IFA | 0 | | Dec 1984 - Aug 1986 | Community | Mixed Groups | Tessier (1987) |
| **Cameroon, Central African Republic, Chad, Republic of the Congo, Equatorial Guinea, Gabon** | | | | | | | | | |
| 0.06 | | Region | IFA | 3 | 5070 | Jan 1985 - Jun 1987 | Population | General Population | Gonzalez (1988) |
| **Central African Republic** | | | | | | | | | |
| | | | IFA | 8 | 4295 | | Population | General Population | Johnson (1993) |
| | | Region | IFA | 5 | 715 | | Unspecified | Unspecified | Georges (1985) |
| **Côte d'Ivoire** | | | | | | | | | |
| 20 | | | IgG | | | | Population | General Population | Emmerich (2008) |
| **Democratic Republic of the Congo, Côte d'Ivoire, Ethiopia, Gabon, Guinea, Kenya, Liberia, Mali, Niger, Nigeria, Tanzania** | | | | | | | | | |
| | | Region | Unspecified | 5 | 712 | | Other | Other | Henderson (1972) |
| **Djibouti** | | | | | | | | | |
| | | Occupation, Symptoms | IFA | 0 | 160 | 1987 | Mixed | Mixed Groups | Salah (1988) |
| **Ethiopia** | | | | | | | | | |
| | | Region | IFA | 0 | | 1961 - 1983 | Unspecified | Unspecified | Tignor (1993) |
| **Gabon** | | | | | | | | | |
| | | | IFA | 0 | 253 | 13 Feb 1980 - 19 Mar 1980 | Population | General Population | Ivanoff (1982) |
| **Germany** | | | | | | | | | |
| 0 | | | IgG | | 94 | | Hospital | Other | ter Meulen (1998) |
| 0 | | | IgG | | 150 | | Unspecified | Other | ter Meulen (1998) |
| | | | IgG | 0 | 200 | | Unspecified | Other | Emmerich (2006) |
| | | | IgM | 0 | 100 | | Unspecified | Other | Emmerich (2006) |
| 0.7 | | | Unspecified | 1 | 150 | | Unspecified | Other | ter Meulen (1998) |
| **Ghana** | | | | | | | | | |
| 3 | | | IgG | 13 | 430 | | Unspecified | Unspecified | Emmerich (2006) |
| 3.8 | | | IgG | | | | Population | General Population | Emmerich (2008) |
| 4 | | | IgG | | 430 | | Unspecified | Unspecified | Emmerich (2006) |
| 5.2 | CI95%: 3.6-7.2 | Age, Other, Sex | IgG | 34 | 657 | Aug 2010 - Aug 2011 | Population | General Population | Nimo-Paintsil (2019) |
| | | | IgM | 0 | 200 | | Unspecified | Unspecified | Emmerich (2006) |
| **Guinea** | | | | | | | | | |
| 2.6 | | | IgG | 6 | 232 | Feb 1993 - Apr 1993 | Population | General Population | ter Meulen (1996) |

continued on next page

continued from previous page

| Seroprevalence (%) | Uncertainty | Disaggregated By | Assay | Number Seropositive | Sample Size | Study Dates | Study Setting | Study Group | Source |
|---|---|---|---|---|---|---|---|---|---|
| 2.6 | | | IgG | | | 1993 | Population | General Population | ter Meulen (1998) |
| 11.3 | CI95%: 9.3-13.3 | Method, Region | IgG | | | 10 Apr 2000 - 22 Apr 2000 | Population | General Population | Kerneis (2009) |
| 11.9 | CI95%: 9.7-14.5 | Region | IgG | 84 | 702 | 2016 | Community | Other | Longet (2023) |
| 12 | | | IgG | | | | Population | General Population | Emmerich (2008) |
| 14 | | Age, Sex | IgG | 105 | 751 | Feb 1993 - Apr 1993 | Population | General Population | ter Meulen (1996) |
| 14 | | | IgG | | | 1993 | Population | General Population | ter Meulen (1998) |
| 15 | | Age, Region | IgG | 48 | 311 | Mar 1996 - Dec 1999 | Hospital | Persons Under Investigation | Bausch (2001) |
| 18.8 | | | IgG | 40 | 213 | | Unspecified | Unspecified | Emmerich (2006) |
| 20 | | | IgG | | | | Population | General Population | Emmerich (2008) |
| 2 - 40 | | Region | IgG | | | Jun 1990 - Jan 1992 | Hospital | Healthcare Workers | Lukashevich (1993) |
| 23.6 | | Age, Region | IgG | 738 | 3126 | Jun 1990 - Jan 1992 | Community | General Population | Lukashevich (1993) |
| 40.3 | | Region, Sex, Time | IgG | 102 | 253 | May 2004 - Oct 2004 | Community | Other | Klempa (2013) |
| | | | IgG | 6 | 12 | | Hospital | Persons Under Investigation | ter Meulen (1998) |
| 59.6 | CI95%: 55.5-63.5 | Region | IgG | 348 | 584 | 2017 - 2018 | Community | Mixed Groups | Longet (2023) |
| | | | IgM | 0 | 100 | | Unspecified | Unspecified | Emmerich (2006) |
| 2.8 | | | IgM | 7 | 253 | May 2004 - Oct 2004 | Community | Other | Klempa (2013) |
| 7 | | Age, Region | IgM | 22 | 311 | Mar 1996 - Dec 1999 | Hospital | Persons Under Investigation | Bausch (2001) |
| 14.9 | | | IgM | 7 | 47 | | Hospital | Other | ter Meulen (1998) |
| 75 | | | IgM | 9 | 12 | | Hospital | Persons Under Investigation | ter Meulen (1998) |
| | | | IgM | 6 | 7 | | Hospital | Other | ter Meulen (1998) |
| 91.7 | | | IgM | 11 | 12 | | Hospital | Persons Under Investigation | ter Meulen (1998) |
| **Guinea, Nigeria** | | | | | | | | | |
| 24 | | | IgG | | 75 | | Unspecified | Unspecified | Gunther (2001) |
| **India** | | | | | | | | | |
| 0 | | Age, Region, Sex | Unspecified | 0 | 1088 | | Unspecified | Unspecified | Rodrigues (1978) |
| **Kenya** | | | | | | | | | |
| | | | IFA | 0 | 1899 | | Population | General Population | Johnson (1983a) |
| | | | IFA | 0 | 741 | Mar 1980 - Dec 1981 | Community | Mixed Groups | Johnson (1983b) |
| **Liberia** | | | | | | | | | |
| 1.5 | | | IFA | 2 | 135 | Feb 1982 - Feb 1982 | Hospital | Children | Monson (1987) |
| 0.9 - 14.1 | | Age, Other, Region, Sex, Time | IFA | | | Sep 1980 - Jan 1982 | Community | General Population | Yalley-Ogunro (1984) |
| 8.4 | | Occupation, Other, Region | IFA | 71 | 844 | Oct 1976 - Oct 1977 | Hospital | Healthcare Workers | Frame (1979) |
| 14.9 | | Other, Region | IFA | 95 | 637 | 1979 - 1982 | Hospital | Healthcare Workers | Frame (1984a) |
| 6 - 28 | | Other, Time | IFA | | | Jul 1980 - Apr 1982 | Hospital | Other | Frame (1984b) |
| 1.3 | | | IgG | 3 | 225 | Nov 1981 - Jan 1982 | Community | Mixed Groups | Van Der Waals (1986) |
| 12.1 | | | IgG | | 99 | 1985 | Unspecified | Unspecified | ter Meulen (1998) |
| | | | Unspecified | 0 | 40 | Mar 1972 - Apr 1972 | Hospital | Healthcare Workers | Monath (1973) |
| | | | Unspecified | 0 | 22 | Mar 1972 - Apr 1972 | Contact | Persons Under Investigation | Monath (1973) |
| 3 | | | Unspecified | 4 | 133 | Mar 1972 - Apr 1972 | Community | General Population | Monath (1973) |
| 3.7 | | | Unspecified | 6 | 165 | Oct 1974 - Dec 1974 | Hospital | Mixed Groups | Bloch (1978) |
| | | | Unspecified | 2 | 17 | Mar 1972 - Apr 1972 | Hospital | Healthcare Workers | Monath (1973) |
| **Madagascar** | | | | | | | | | |
| 0 | | | IFA | 0 | 381 | | Unspecified | Unspecified | Mathiot (1989) |
| **Mali** | | | | | | | | | |
| 33.2 | CI95%: 29.4-37.1 | Age, Region, Sex | IgG | 199 | 600 | Feb 2015 - Feb 2015 | Community | General Population | Sogoba (2016) |
| 0.67 | | Region | IgM | 4 | 600 | Feb 2015 - Feb 2015 | Community | General Population | Sogoba (2016) |
| **Nigeria** | | | | | | | | | |
| 0 | | | IFA | 0 | 47 | 1 Jan 1989 - 31 Mar 1989 | Hospital | Healthcare Workers | Fisher-Hoch (1995) |

continued on next page

continued from previous page

| Seroprevalence (%) | Uncertainty | Disaggregated By | Assay | Number Seropositive | Sample Size | Study Dates | Study Setting | Study Group | Source |
|---|---|---|---|---|---|---|---|---|---|
| 1 | | | IFA | 6 | 415 | 1 Jan 1989 - 31 Mar 1989 | Community | Unspecified | Fisher-Hoch (1995) |
| 1 | | | IFA | 2 | 175 | 1 Jan 1989 - 31 Mar 1989 | Hospital | Healthcare Workers | Fisher-Hoch (1995) |
| 2 | | | IFA | 5 | 240 | 1 Jan 1989 - 31 Mar 1989 | Hospital | Healthcare Workers | Fisher-Hoch (1995) |
| 21.3 | | Region | IFA | 357 | 1677 | | Population | General Population | Tomori (1988) |
| 0 | | | IgG | 0 | 12 | | Hospital | Other | Shaibu (2021) |
| 1.8 | | | IgG | 8 | 451 | | Hospital | Persons Under Investigation | Ehichioya (2012) |
| 7.4 | | | IgG | 22 | 297 | Sep 2011 - Feb 2012 | Hospital | Other | Bukbuk (2014) |
| 10.3 | | | IgG | | | | Population | General Population | Emmerich (2008) |
| 12.3 | | Occupation, Region | IgG | 63 | 552 | Dec 1992 - Mar 1993 | Hospital | Healthcare Workers | Bajani (1997) |
| 45.7 | CI95%: 38.4-53.1 | | IgG | 77 | 170 | 12 Feb 2019 - 20 Dec 2019 | Hospital | Pregnant Women | Kayem (2023b) |
| 49.6 | CI95%: 43.3-55.9 | Other, Time | IgG | 119 | 240 | Feb 2019 - Dec 2019 | Hospital | Pregnant Women | Kayem (2023a) |
| 50 - 64 | | Method | IgG | | | Oct 2018 - Oct 2020 | Contact | Persons Under Investigation | Ugwu (2022) |
| 58.2 | | | IgG | 103 | 177 | | Community | General Population | Tobin (2015) |
| 48 - 77 | | Method | IgG | | | Oct 2018 - Oct 2020 | Hospital | Other | Ugwu (2022) |
| 0 | | | IgM | 0 | 451 | | Hospital | Persons Under Investigation | Ehichioya (2012) |
| 0 | | | IgM | 0 | 12 | | Hospital | Other | Shaibu (2021) |
| | | | IgM | 6 | 552 | Dec 1992 - Mar 1993 | Hospital | Healthcare Workers | Bajani (1997) |
| 1.1 | | | IgM | 2 | 177 | | Community | General Population | Tobin (2015) |
| 8.6 | | Level of Exposure, Occupation, Other, Region | IgM | 20 | 233 | Mar 2018 - Apr 2019 | Mixed | Mixed Groups | Shaibu (2021) |
| 23 - 32 | | Method | IgM | | | Oct 2018 - Oct 2020 | Hospital | Other | Ugwu (2022) |
| 33 - 48 | | Method | IgM | | | Oct 2018 - Oct 2020 | Contact | Persons Under Investigation | Ugwu (2022) |
| 5.4 | | Age, Occupation, Region, Sex | PRNT | 18 | 336 | 3 Mar 1971 - Mar 1971 | Community | Mixed Groups | Arnold (1977) |
| 5.8 | | Age, Occupation, Region, Sex | PRNT | 25 | 434 | 24 Aug 1970 - 24 Aug 1970 | Community | Mixed Groups | Arnold (1977) |
| | | | Unspecified | 0 | 22 | Jan 1970 - Feb 1970 | Hospital | Healthcare Workers | Troup (1970) |
| | | | Unspecified | 0 | 258 | | Mixed | Mixed Groups | Bowen (1975) |
| | | | Unspecified | 10 | 458 | 1965 - 1966 | Community | General Population | Henderson (1972) |
| | | | Unspecified | 4 | 53 | Jan 1970 - Feb 1970 | Contact | Persons Under Investigation | Troup (1970) |
| | | Age, Region | Unspecified | 23 | 281 | 1965 - 1970 | Community | General Population | Henderson (1972) |
| 15.5 | | Age, Occupation, Sex | Unspecified | 15 | 97 | Aug 1976 - Aug 1976 | Trade | Mixed Groups | Smith (1979) |
| 16.1 | | | Unspecified | 10 | 62 | | Community | Unspecified | Bajani (1997) |
| 36.1 | | Level of Exposure, Occupation, Other, Region | Unspecified | 84 | 233 | Mar 2018 - Apr 2019 | Mixed | Mixed Groups | Shaibu (2021) |
| **Sierra Leone** | | | | | | | | | |
| 21 | | | IFA | 51 | 245 | Aug 1977 - Aug 1981 | Hospital | Children | Webb (1986) |
| 22 | | Age, Level of Exposure, Occupation, Sex | IFA | 108 | 496 | 1972 - 1983 | Hospital | Healthcare Workers | Helmick (1986) |
| 26 | | Level of Exposure | IFA | 248 | 953 | | Household | Mixed Groups | Keenlyside (1983) |
| 0 - 52 | | Age, Region, Sex | IFA | | | | Community | General Population | McCormick (1987b) |
| 35 | | Age, Sex | IFA | 360 | 1024 | 1978 - 1981 | Community | General Population | Helmick (1986) |
| 10.61 | | Region | IgG | 339 | 3196 | Jul 2015 - Jun 2018 | Community | General Population | Grant (2023) |
| 14.14 | | Region | IgG | 322 | 2278 | Jul 2015 - Jun 2018 | Community | General Population | Grant (2023) |
| 18 | | | IgG | 125 | 688 | 2011 - 2012 | Hospital | Persons Under Investigation | Shaffer (2014) |
| 20.12 | | Region | IgG | 1040 | 5168 | Jul 2015 - Jun 2018 | Population | General Population | Grant (2023) |
| 33.2 | | Other, Time | IgG | 682 | 2051 | 1 Jan 2012 - 31 Dec 2019 | Hospital | Persons Under Investigation | Shaffer (2021) |

continued on next page

continued from previous page

| Seroprevalence (%) | Uncertainty | Disaggregated By | Assay | Number Seropositive | Sample Size | Study Dates | Study Setting | Study Group | Source |
|---|---|---|---|---|---|---|---|---|---|
| 3 - 23 | | Method, Other, Time | IgM | | | 2008 - 2012 | Hospital | Persons Under Investigation | Shaffer (2014) |
| 18 - 22 | | Disease Generation, Time | IgM | | | 2008 - 2016 | Hospital | Persons Under Investigation | Shaffer (2019) |
| 25.6 | | Other, Time | IgM | 838 | 3277 | 1 Jan 2012 - 31 Dec 2019 | Hospital | Persons Under Investigation | Shaffer (2021) |
| 6 | | Age, Other, Region, Sex | Unspecified | 16 | 255 | 23 Sep 1972 - 15 Oct 1972 | Household | General Population | Fraser (1974) |
| 0 - 13 | | Region | Unspecified | | | 23 Sep 1972 - 15 Oct 1972 | Hospital | Healthcare Workers | Fraser (1974) |
| 7 | | Region | Unspecified | 12 | 170 | Nov 1974 - Nov 1974 | Mixed | Mixed Groups | Fabiyi (1975) |
| | | | Unspecified | 5 | 50 | | Hospital | Children | Sharp (1982) |
| 13 | | Age, Sex | Unspecified | 27 | 206 | 23 Sep 1972 - 15 Oct 1972 | Contact | Persons Under Investigation | Fraser (1974) |
| 0 - 29.4 | | Age | Unspecified | | | | Household | Children | Sharp (1982) |
| | | | Unspecified | 12 | 72 | 20 May 2019 - 24 May 2019 | School | General Population | Akpogheneta (2021) |
| 22 | | Occupation | Unspecified | 12 | 54 | 23 Sep 1972 - 15 Oct 1972 | Hospital | Healthcare Workers | Fraser (1974) |
| 81.7 | | | Unspecified | 58 | 71 | 3 Jun 2019 - 28 Sep 2019 | Unspecified | Other | Akpogheneta (2021) |
| | | | Unspecified | 40 | 45 | Jan 1973 - Mar 1976 | Hospital | Persons Under Investigation | Keane (1977) |
| **Sweden** | | | | | | | | | |
| 0 | | | IgG | 0 | 53 | Mar 2016 | Contact | Healthcare Workers | Grahn (2018) |
| **Uganda** | | | | | | | | | |
| 6 | | Region | IFA | 8 | 132 | May 1984 - May 1984 | Hospital | Other | Rodhain (1989) |
| 0.1 | | | IgG | 2 | 1744 | 2006 - 2007 | Population | Other | Clements (2019) |
| **United Kingdom** | | | | | | | | | |
| 0 | | | Unspecified | 0 | 159 | 1982 | Contact | Mixed Groups | Cooper (1982) |
| **United States** | | | | | | | | | |
| 0 | | | IFA | 0 | 29 | | Contact | Persons Under Investigation | Zweighaft (1977) |
| **Unspecified** | | | | | | | | | |
| 0 | | | Unspecified | 0 | 86 | Nov 1975 - Jun 1977 | Travel | Persons Under Investigation | Woodruff (1978) |
| **Zimbabwe** | | | | | | | | | |
| 0 | | Region | IFA | 0 | 486 | 1980 | School | Children | Blackburn (1982) |

Table B.11: Seroprevalence grouped by country. Study characteristics are reported for each seroprevalence estimate.

| Risk Factor | Significance | Method | Sample Size | Country | Study Dates | Study Setting | Study Group | Source |
|---|---|---|---|---|---|---|---|---|
| **Occurrence** | | | | | | | | |
| Contact with Animal, Other | Significant | Adjusted | 90 | Sierra Leone | Sep 2002 - Jul 2004 | Household | Other | Bonner (2007) |
| Contact with Animal, Other | Significant | Not Adjusted | 90 | Sierra Leone | Sep 2002 - Jul 2004 | Household | Other | Bonner (2007) |
| Other | Significant | Adjusted | 73 | Liberia | 2008 - 2012 | Community | Outdoor Workers | Olugasa (2014) |
| Other | Significant | Adjusted | 3096 | Nigeria | 2016 - 2019 | Population | General Population | Redding (2021) |
| Other | Not Significant | Adjusted | 73 | Liberia | 2008 - 2012 | Community | Outdoor Workers | Olugasa (2014) |
| Other | Not Significant | Adjusted | 90 | Sierra Leone | Sep 2002 - Jul 2004 | Household | Other | Bonner (2007) |
| Other | Not Significant | Not Adjusted | 90 | Sierra Leone | Sep 2002 - Jul 2004 | Household | Other | Bonner (2007) |
| **Infection** | | | | | | | | |
| Age | Significant | Not Adjusted | | Nigeria | Jan 2009 - Dec 2010 | Hospital | Persons Under Investigation | Asogun (2012) |
| Age, Hospitalisation, Other | Significant | Not Adjusted | | Sierra Leone | 1 Jan 2012 - 31 Dec 2019 | Hospital | Persons Under Investigation | Shaffer (2021) |
| Age, Occupation | Significant | Adjusted | 423 | Nigeria | 1 Jan 2018 - 6 May 2018 | Hospital | Persons Under Investigation | Ilori (2019b) |
| Age, Occupation, Other, Sex | Significant | Adjusted | 20027 | Nigeria | 2018 - 2021 | Hospital | Persons Under Investigation | Ochu (2023) |
| Age, Occupation, Other, Sex | Significant | Not Adjusted | 20027 | Nigeria | 2018 - 2021 | Hospital | Persons Under Investigation | Ochu (2023) |
| Age, Occupation, Other, Sex | Significant | Not Adjusted | 1991 | Nigeria | Jan 2018 - Jun 2019 | Population | Persons Under Investigation | Olayinka (2022) |
| Household Contact | Significant | Unspecified | | Nigeria | 3 Mar 1971 - Mar 1971 | Community | Mixed Groups | Arnold (1977) |
| Occupation, Other, Sex | Significant | Adjusted | 1991 | Nigeria | Jan 2018 - Jun 2019 | Population | Persons Under Investigation | Olayinka (2022) |
| Other | Significant | Not Adjusted | | Guinea | Mar 1996 - Dec 1999 | Hospital | Persons Under Investigation | Bausch (2001) |
| Other | Significant | Unspecified | 373 | Nigeria | 1 Dec 2009 - 30 Nov 2010 | Hospital | Children | Akhuemokhan (2017) |
| Other | Significant | Unspecified | 2787 | Nigeria | Dec 2016 - Sep 2020 | Hospital | Persons Under Investigation | Yaro (2021) |
| Sex | Significant | Not Adjusted | 245 | Sierra Leone | Aug 1977 - Aug 1981 | Hospital | Children | Webb (1986) |
| Age, Occupation, Other | Not Significant | Adjusted | 20027 | Nigeria | 2018 - 2021 | Hospital | Persons Under Investigation | Ochu (2023) |
| Age, Occupation, Other | Not Significant | Adjusted | 1991 | Nigeria | Jan 2018 - Jun 2019 | Population | Persons Under Investigation | Olayinka (2022) |
| Age, Occupation, Other | Not Significant | Not Adjusted | 20027 | Nigeria | 2018 - 2021 | Hospital | Persons Under Investigation | Ochu (2023) |
| Age, Occupation, Other, Sex | Not Significant | Not Adjusted | 34 | Nigeria | Jan 2016 - Aug 2016 | Hospital | Persons Under Investigation | Shehu (2018) |
| Age, Other | Not Significant | Not Adjusted | 1991 | Nigeria | Jan 2018 - Jun 2019 | Population | Persons Under Investigation | Olayinka (2022) |
| Age, Other, Sex | Not Significant | Unspecified | 373 | Nigeria | 1 Dec 2009 - 30 Nov 2010 | Hospital | Children | Akhuemokhan (2017) |
| Contact with Animal, Funeral, Sex | Not Significant | Adjusted | 423 | Nigeria | 1 Jan 2018 - 6 May 2018 | Hospital | Persons Under Investigation | Ilori (2019b) |
| Occupation, Other, Sex | Not Significant | Not Adjusted | | Nigeria | Jan 2009 - Dec 2010 | Hospital | Persons Under Investigation | Asogun (2012) |
| Other, Sex | Not Significant | Not Adjusted | | Sierra Leone | 1 Jan 2012 - 31 Dec 2019 | Hospital | Persons Under Investigation | Shaffer (2021) |
| Occupation | Unspecified | Unspecified | 3162 | Nigeria | Jan 2018 - Dec 2021 | Hospital | Persons Under Investigation | Dalhat (2022) |
| **Reproduction Number** | | | | | | | | |
| Other | Significant | Not Adjusted | | Nigeria | | Population | General Population | Zhao (2020) |
| **Attack Rate** | | | | | | | | |
| Age | Significant | Not Adjusted | | Sierra Leone | 1 Oct 1970 - 1 Oct 1972 | Population | General Population | Fraser (1974) |
| Sex | Not Significant | Not Adjusted | | Sierra Leone | 1 Oct 1970 - 1 Oct 1972 | Population | General Population | Fraser (1974) |
| **Incidence** | | | | | | | | |
| Other | Significant | Adjusted | 3096 | Nigeria | 2016 - 2019 | Population | General Population | Redding (2021) |
| **Onset-Admission Delay** | | | | | | | | |
| Age, Close Contact, Funeral, Household Contact, Non-household Contact, Occupation, Other, Sex | Not Significant | Not Adjusted | 389 | Nigeria | Dec 2018 - Apr 2019 | Population | Persons Under Investigation | Chandra (2021) |
| **Viremia** | | | | | | | | |
| Other | Not Significant | Unspecified | | Sierra Leone | 1977 - | Community | Persons Under Investigation | McCormick (1986) |
| **Death** | | | | | | | | |
| Age | Significant | Not Adjusted | | Nigeria | Jan 2009 - Dec 2010 | Hospital | Persons Under Investigation | Asogun (2012) |
| Age, Occupation, Other | Significant | Not Adjusted | 724 | Nigeria | Jan 2018 - Jun 2019 | Population | Persons Under Investigation | Olayinka (2022) |
| Age, Other | Significant | Adjusted | 284 | Nigeria | Jan 2011 - Nov 2015 | Hospital | Persons Under Investigation | Okokhere (2018) |
| Age, Other | Significant | Adjusted | 510 | Nigeria | 5 Apr 2018 - 15 Mar 2020 | Hospital | Persons Under Investigation | Duvignaud (2021) |
| Age, Other | Significant | Adjusted | 103 | Liberia | Jan 2019 - Dec 2020 | Hospital | Persons Under Investigation | Jetoh (2022) |
| Age, Other | Significant | Adjusted | 724 | Nigeria | Jan 2018 - Jun 2019 | Population | Persons Under Investigation | Olayinka (2022) |
| Age, Other | Significant | Not Adjusted | 47 | Nigeria | Oct 2015 - Feb 2016 | Hospital | Persons Under Investigation | Buba (2018) |

continued on next page

continued from previous page

| Risk Factor | Significance | Method | Sample Size | Country | Study Dates | Study Setting | Study Group | Source |
|---|---|---|---|---|---|---|---|---|
| Age, Other | Significant | Not Adjusted | 510 | Nigeria | 5 Apr 2018 - 15 Mar 2020 | Hospital | Persons Under Investigation | Duvignaud (2021) |
| Other | Significant | Adjusted | | Sierra Leone | 1977 - | Hospital | Persons Under Investigation | McCormick (1986) |
| Other | Significant | Adjusted | 76 | Nigeria | Jan 2015 - Dec 2018 | Hospital | Persons Under Investigation | Abdulkarim (2020) |
| Other | Significant | Adjusted | 423 | Nigeria | 1 Jan 2018 - 6 May 2018 | Hospital | Persons Under Investigation | Ilori (2019b) |
| Other | Significant | Adjusted | 83 | Nigeria | Dec 2017 - Dec 2018 | Hospital | Persons Under Investigation | Chika-Igwenyi (2021) |
| Other | Significant | Adjusted | 201 | Nigeria | Jan 2014 - Apr 2017 | Hospital | Persons Under Investigation | Strampe (2021) |
| Other | Significant | Not Adjusted | 68 | Sierra Leone | 1981 - 1985 | Hospital | Pregnant Women | Price (1988) |
| Other | Significant | Not Adjusted | | Sierra Leone | 1 Jan 2012 - 31 Dec 2019 | Hospital | Persons Under Investigation | Shaffer (2021) |
| Other | Significant | Not Adjusted | 1298 | Nigeria | 2008 - 2018 | Hospital | Persons Under Investigation | Akpede (2019) |
| Other | Significant | Not Adjusted | 57 | Sierra Leone | 1 Jan 2012 - 31 Dec 2018 | Hospital | Children | Samuels (2021) |
| Other | Significant | Unspecified | 1740 | Sierra Leone | 2008 - 2012 | Hospital | Persons Under Investigation | Shaffer (2014) |
| Other | Significant | Unspecified | 20 | Nigeria | 1 Jan 2012 - 25 Mar 2012 | Hospital | Persons Under Investigation | Ajayi (2013) |
| Other | Significant | Unspecified | 57 | Nigeria | Jan 2009 - Aug 2017 | Hospital | Children | Adetunji (2021) |
| Age, Hospitalisation, Other, Sex | Not Significant | Not Adjusted | | Sierra Leone | 1 Jan 2012 - 31 Dec 2019 | Hospital | Persons Under Investigation | Shaffer (2021) |
| Age, Other, Sex | Not Significant | Adjusted | 76 | Nigeria | Jan 2015 - Dec 2018 | Hospital | Persons Under Investigation | Abdulkarim (2020) |
| Age, Other, Sex | Not Significant | Not Adjusted | 57 | Sierra Leone | 1 Jan 2012 - 31 Dec 2018 | Hospital | Children | Samuels (2021) |
| Age, Sex | Not Significant | Unspecified | 83 | Nigeria | Dec 2017 - Dec 2018 | Hospital | Persons Under Investigation | Chika-Igwenyi (2021) |
| Cormobidity, Other, Sex | Not Significant | Not Adjusted | 47 | Nigeria | Oct 2015 - Feb 2016 | Hospital | Persons Under Investigation | Buba (2018) |
| Cormobidity, Other, Sex | Not Significant | Not Adjusted | 510 | Nigeria | 5 Apr 2018 - 15 Mar 2020 | Hospital | Persons Under Investigation | Duvignaud (2021) |
| Occupation, Other, Sex | Not Significant | Not Adjusted | 724 | Nigeria | Jan 2018 - Jun 2019 | Population | Persons Under Investigation | Olayinka (2022) |
| Occupation, Sex | Not Significant | Adjusted | 103 | Liberia | Jan 2019 - Dec 2020 | Hospital | Persons Under Investigation | Jetoh (2022) |
| Occupation, Sex | Not Significant | Not Adjusted | | Nigeria | Jan 2009 - Dec 2010 | Hospital | Persons Under Investigation | Asogun (2012) |
| Other | Not Significant | Adjusted | 284 | Nigeria | Jan 2011 - Nov 2015 | Hospital | Persons Under Investigation | Okokhere (2018) |
| Other | Not Significant | Adjusted | 724 | Nigeria | Jan 2018 - Jun 2019 | Population | Persons Under Investigation | Olayinka (2022) |
| Other | Not Significant | Not Adjusted | 147 | Sierra Leone | 1981 - 1985 | Hospital | Other | Price (1988) |
| Other | Not Significant | Not Adjusted | | Sierra Leone | 1977 - | Hospital | Persons Under Investigation | McCormick (1986) |
| Other | Not Significant | Unspecified | 68 | Sierra Leone | 1981 - 1985 | Hospital | Pregnant Women | Price (1988) |
| Sex | Not Significant | Adjusted | 510 | Nigeria | 5 Apr 2018 - 15 Mar 2020 | Hospital | Persons Under Investigation | Duvignaud (2021) |
| Sex | Not Significant | Not Adjusted | 245 | Sierra Leone | Aug 1977 - Aug 1981 | Hospital | Children | Webb (1986) |
| Sex | Not Significant | Not Adjusted | 284 | Nigeria | Jan 2011 - Nov 2015 | Hospital | Persons Under Investigation | Okokhere (2018) |
| Sex | Not Significant | Unspecified | 1740 | Sierra Leone | 2008 - 2012 | Hospital | Persons Under Investigation | Shaffer (2014) |
| Age, Sex | Unspecified | Unspecified | 3162 | Nigeria | Jan 2018 - Dec 2021 | Hospital | Persons Under Investigation | Dalhat (2022) |
| **Serology** | | | | | | | | |
| Age, Hospitalisation, Other | Significant | Not Adjusted | | Sierra Leone | 1 Jan 2012 - 31 Dec 2019 | Hospital | Persons Under Investigation | Shaffer (2021) |
| Age, Household Contact | Significant | Not Adjusted | 953 | Sierra Leone | | Household | Mixed Groups | Keenlyside (1983) |
| Age, Other | Significant | Adjusted | 600 | Mali | Feb 2015 - Feb 2015 | Community | General Population | Sogoba (2016) |
| Age, Other | Significant | Adjusted | 702 | Guinea | 2016 | Community | Other | Longet (2023) |
| Age, Other | Significant | Not Adjusted | 71 | Sierra Leone | 3 Jun 2019 - 28 Sep 2019 | Unspecified | Other | Akpogheneta (2021) |
| Age, Other | Significant | Unspecified | 177 | Nigeria | | Community | General Population | Tobin (2015) |
| Close Contact, Occupation | Significant | Adjusted | | Sierra Leone | 1972 - 1983 | Mixed | Mixed Groups | Helmick (1986) |
| Contact with Animal, Other | Significant | Adjusted | 240 | Nigeria | Feb 2019 - Dec 2019 | Hospital | Pregnant Women | Kayem (2023a) |
| Household Contact, Other | Significant | Adjusted | 72 | Sierra Leone | 20 May 2019 - 24 May 2019 | School | General Population | Akpogheneta (2021) |
| Household Contact, Other | Significant | Not Adjusted | 206 | Sierra Leone | 23 Sep 1972 - 15 Oct 1972 | Household | Mixed Groups | Fraser (1974) |
| Occupation | Significant | Not Adjusted | | Sierra Leone | 1972 - 1983 | Mixed | Mixed Groups | Helmick (1986) |
| Occupation | Significant | Not Adjusted | | Nigeria | Dec 1992 - Mar 1993 | Hospital | Healthcare Workers | Bajani (1997) |
| Other | Significant | Adjusted | 1424 | Guinea | 10 Apr 2000 - 22 Apr 2000 | Population | General Population | Kerneis (2009) |
| Other | Significant | Adjusted | 584 | Guinea | 2017 - 2018 | Community | Mixed Groups | Longet (2023) |
| Other | Significant | Not Adjusted | 1424 | Guinea | 10 Apr 2000 - 22 Apr 2000 | Population | General Population | Kerneis (2009) |
| Other | Significant | Not Adjusted | | Guinea | Mar 1996 - Dec 1999 | Hospital | Persons Under Investigation | Bausch (2001) |
| Age, Contact with Animal, Funeral, Occupation, Other, Sex | Not Significant | Not Adjusted | 1424 | Guinea | 10 Apr 2000 - 22 Apr 2000 | Population | General Population | Kerneis (2009) |

continued on next page

continued from previous page

| Risk Factor | Significance | Method | Sample Size | Country | Study Dates | Study Setting | Study Group | Source |
|---|---|---|---|---|---|---|---|---|
| Age, Contact with Animal, Other, Sex | Not Significant | Adjusted | 991 | Guinea | Feb 1993 - Apr 1993 | Population | General Population | ter Meulen (1996) |
| Age, Contact with Animal, Other, Sex | Not Significant | Not Adjusted | 991 | Guinea | Feb 1993 - Apr 1993 | Population | General Population | ter Meulen (1996) |
| Age, Other, Sex | Not Significant | Adjusted | 253 | Guinea | May 2004 - Oct 2004 | Community | Other | Klempa (2013) |
| Age, Sex | Not Significant | Adjusted | 584 | Guinea | 2017 - 2018 | Community | Mixed Groups | Longet (2023) |
| Close Contact, Occupation | Not Significant | Adjusted | | Sierra Leone | 1972 - 1983 | Mixed | Mixed Groups | Helmick (1986) |
| Contact with Animal | Not Significant | Adjusted | 1424 | Guinea | 10 Apr 2000 - 22 Apr 2000 | Population | General Population | Kerneis (2009) |
| Other | Not Significant | Adjusted | 240 | Nigeria | Feb 2019 - Dec 2019 | Hospital | Pregnant Women | Kayem (2023a) |
| Other | Not Significant | Not Adjusted | 953 | Sierra Leone | | Household | Mixed Groups | Keenlyside (1983) |
| Other, Sex | Not Significant | Not Adjusted | | Sierra Leone | 1 Jan 2012 - 31 Dec 2019 | Hospital | Persons Under Investigation | Shaffer (2021) |
| Other, Sex | Not Significant | Unspecified | 177 | Nigeria | | Community | General Population | Tobin (2015) |
| Sex | Not Significant | Adjusted | 600 | Mali | Feb 2015 - Feb 2015 | Community | General Population | Sogoba (2016) |
| Sex | Not Significant | Adjusted | 702 | Guinea | 2016 | Community | Other | Longet (2023) |
| Sex | Not Significant | Unspecified | 1677 | Nigeria | | Population | General Population | Tomori (1988) |
| Age | Unspecified | Unspecified | 1677 | Nigeria | | Population | General Population | Tomori (1988) |

Table B.12: Risk factors grouped by outcome. Study characteristics are reported for each set of risk factors.

### C Additional Figures

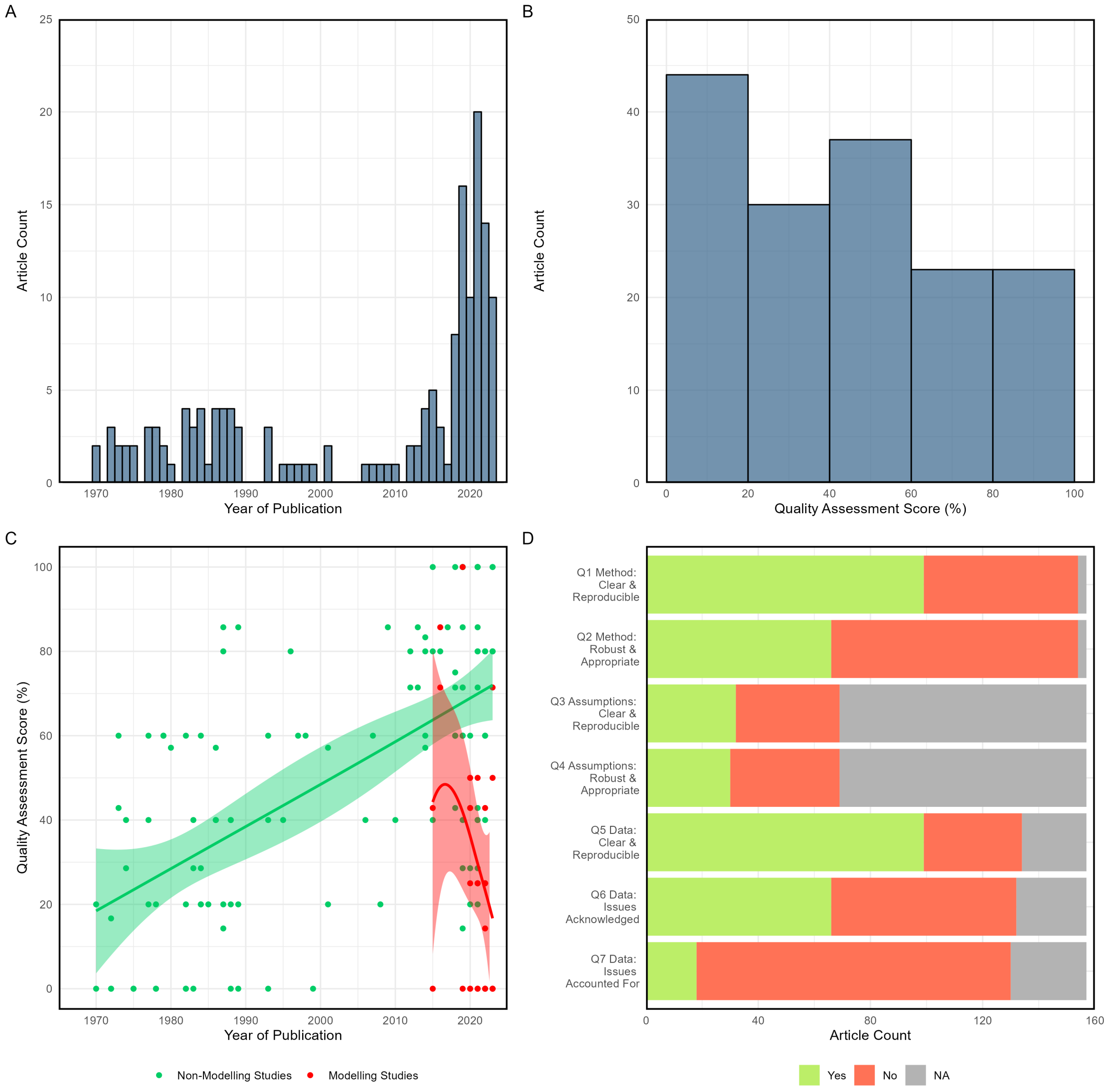

Figure C.1: (A) Article count by year of publication; (B) article count by quality assessment score (defined as percentage of 'Yes' answers relative to sum of 'Yes' and 'No' answers for each paper, removing 'NAs'); (C) quality assessment score by year of publication (time trends for articles with and without transmission models fitted via local polynomial regression); (D) article count for each quality assessment question scoring 'Yes', 'No' or 'Non-Applicable'.

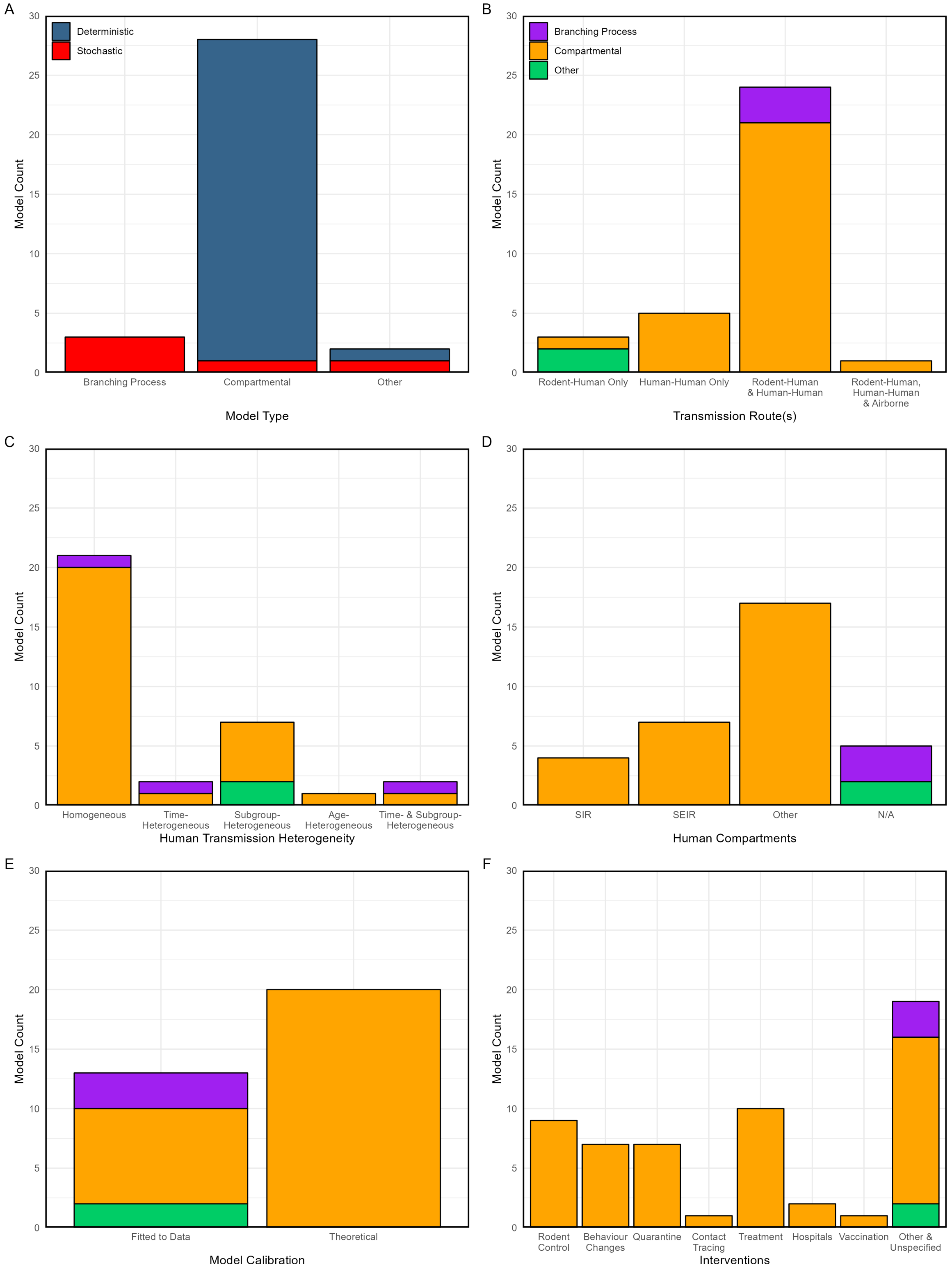

Figure C.2: Transmission model count by (A) model type, (B) transmission routes, (C) human transmission heterogeneity, (D) human compartments (where applicable, otherwise listed as NA for non-compartmental model types), (E) calibration of model outputs, (F) interventions incorporated. Colour corresponds to the deterministic or stochastic nature of the model in (A) and the model type in (B-F).

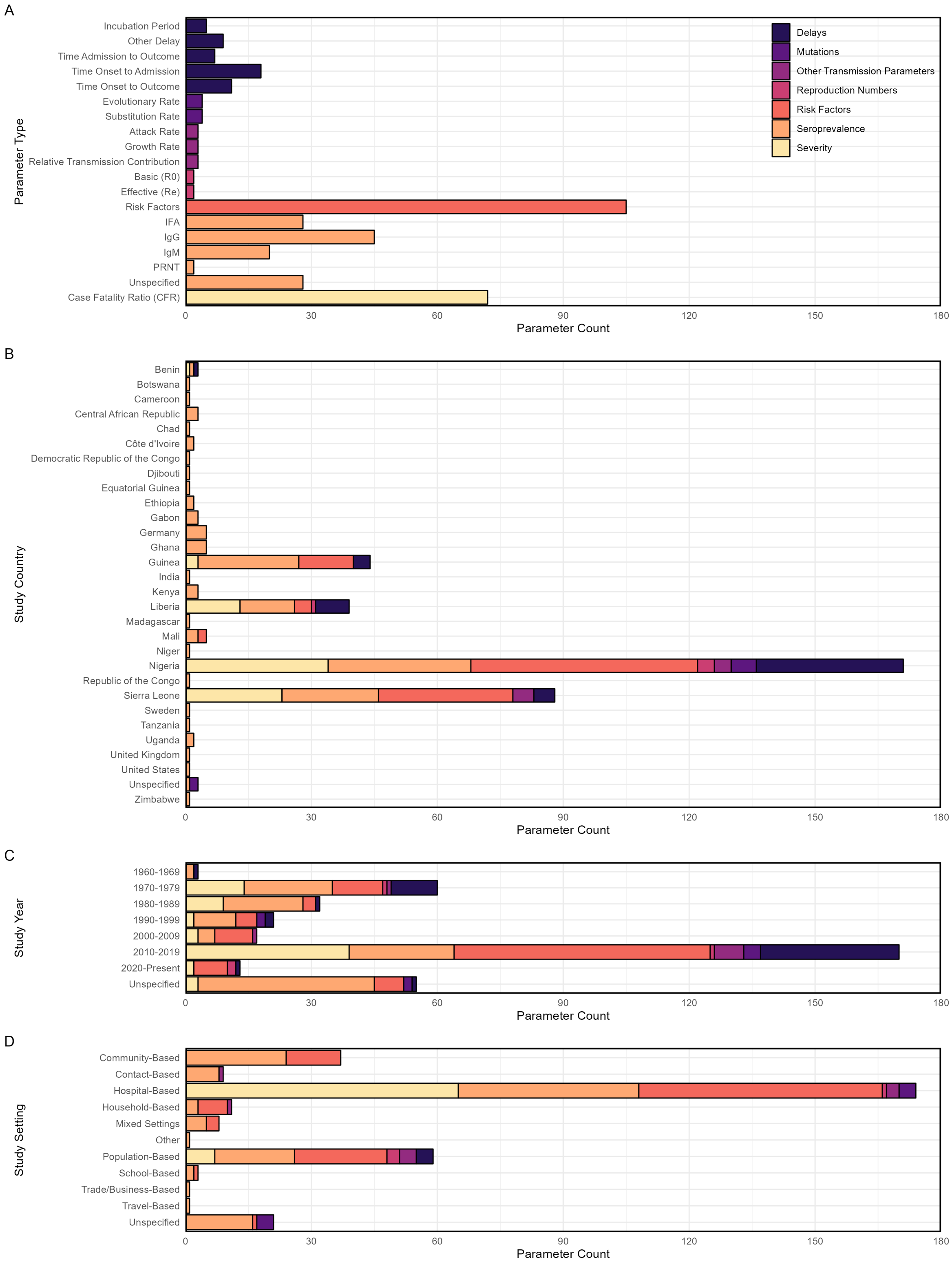

Figure C.3: Parameter type count by (A) parameter class, (B) survey country/countries, (C) survey decade (D) study setting. Colour corresponds to parameter class.

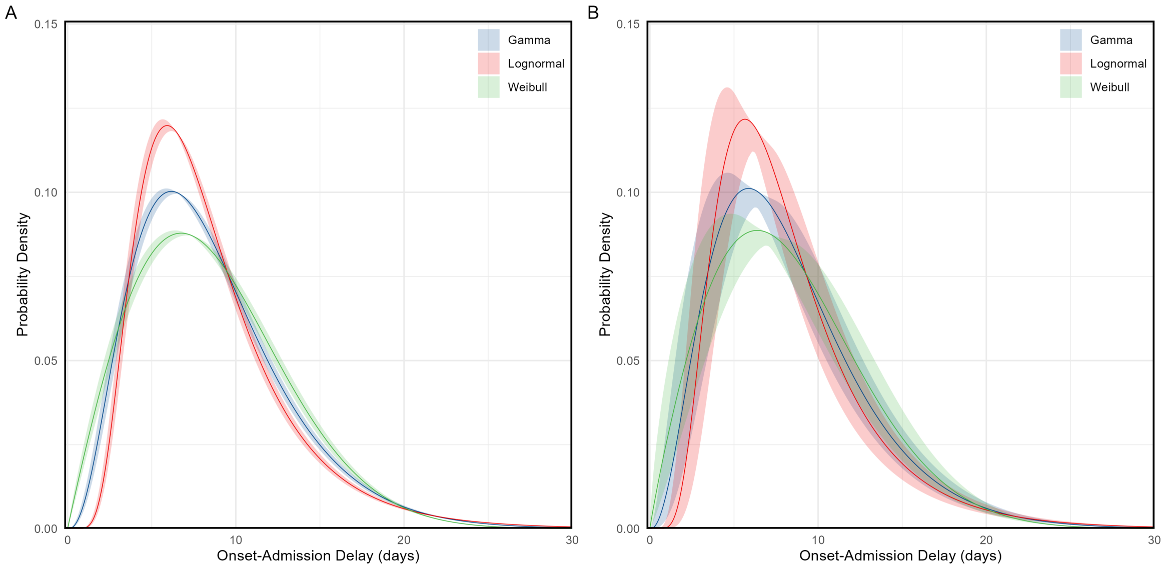

Figure C.4: Hypothetical symptom onset to hospital admission delay probability distributions for the (A) common- and (B) random-effects models. These are constructed using the pooled mean estimate and corresponding standard error from the meta-analysis (main text figure 4E) and the separately-calculated pooled standard deviation, and shown for Gamma, Lognormal and Weibull distributions for illustration. The shaded areas represent the 95% confidence in the mean value estimated in the meta-analysis.

| Study or Subgroup | Events | Total | GLMM, Fixed + Random, 95% CI |
|---|---|---|---|
| **Lineage II Region - Nigeria** | | | |
| Fisher-Hoch (1995) | 22 | 34 | 0.647 [0.465; 0.803] |
| Isere (2018) | 12 | 19 | 0.632 [0.384; 0.837] |
| Ajayi (2013) | 6 | 20 | 0.300 [0.119; 0.543] |
| Inegbenebor (2010) | 18 | 64 | 0.281 [0.176; 0.408] |
| Akpede (2019) | 362 | 1298 | 0.279 [0.255; 0.304] |
| **Total (common effect, 95% CI)** | **420** | **1435** | **0.293 [0.270; 0.317]** |
| **Total (random effect, 95% CI)** | | | **0.406 [0.265; 0.565]** |
| Heterogeneity: $Tau^2 = 0.4109$; $Chi^2 = 27.37$, df = 4 ($P < 0.01$); $I^2 = 85\%$ | | | |
| **Lineage III Region - Nigeria** | | | |
| Gomerep (2022) | 25 | 44 | 0.568 [0.410; 0.717] |
| Abdulkarim (2020) | 41 | 76 | 0.539 [0.421; 0.655] |
| White (1972) | 12 | 23 | 0.522 [0.306; 0.732] |
| **Total (common effect, 95% CI)** | **78** | **143** | **0.545 [0.463; 0.625]** |
| **Total (random effect, 95% CI)** | | | **0.545 [0.463; 0.625]** |
| Heterogeneity: $Tau^2 = 0$; $Chi^2 = 0.15$, df = 2 ($P = 0.93$); $I^2 = 0\%$ | | | |
| **Lineage IV Region - Guinea, Liberia, Sierra Leone** | | | |
| Keane (1977) | 9 | 12 | 0.750 [0.428; 0.945] |
| Shaffer (2014) | 109 | 158 | 0.690 [0.612; 0.761] |
| Samuels (2021) | 36 | 57 | 0.632 [0.493; 0.756] |
| Dahmane (2014) | 22 | 36 | 0.611 [0.435; 0.769] |
| Njuguna (2022) | 3 | 5 | 0.600 [0.147; 0.947] |
| Jetoh (2022) | 41 | 103 | 0.398 [0.303; 0.499] |
| Fraser (1974) | 24 | 63 | 0.381 [0.261; 0.512] |
| Monath (1973) | 4 | 11 | 0.364 [0.109; 0.692] |
| ter Meulen (1998) | 4 | 12 | 0.333 [0.099; 0.651] |
| Keane (1977) | 6 | 18 | 0.333 [0.133; 0.590] |
| Hamblion (2018) | 4 | 14 | 0.286 [0.084; 0.581] |
| Price (1988) | 14 | 68 | 0.206 [0.117; 0.321] |
| Bausch (2001) | 4 | 22 | 0.182 [0.052; 0.403] |
| Keane (1977) | 16 | 96 | 0.167 [0.098; 0.256] |
| McCormick (1987a) | 73 | 441 | 0.166 [0.132; 0.204] |
| Keane (1977) | 22 | 138 | 0.159 [0.103; 0.231] |
| Price (1988) | 10 | 79 | 0.127 [0.062; 0.220] |
| Webb (1986) | 6 | 51 | 0.118 [0.044; 0.239] |
| Frame (1989) | 23 | 246 | 0.093 [0.060; 0.137] |
| Bonner (2007) | 8 | 90 | 0.089 [0.039; 0.168] |
| **Total (common effect, 95% CI)** | **438** | **1720** | **0.255 [0.235; 0.276]** |
| **Total (random effect, 95% CI)** | | | **0.289 [0.203; 0.393]** |
| Heterogeneity: $Tau^2 = 0.9630$; $Chi^2 = 282.26$, df = 19 ($P < 0.01$); $I^2 = 93\%$ | | | |
| **Lineage VII Region - Benin,Togo** | | | |
| Yadouleton (2020) | 8 | 18 | 0.444 [0.215; 0.692] |
| **Unspecified - Nigeria** | | | |
| Dalhat (2022) | 550 | 3162 | 0.174 [0.161; 0.188] |
| **Total (common effect, 95% CI)** | **1494** | **6478** | **0.231 [0.221; 0.241]** |
| **Total (random effect, 95% CI)** | | | **0.331 [0.257; 0.415]** |
| Heterogeneity: $Tau^2 = 0.8555$; $Chi^2 = 482.78$, df = 29 ($P < 0.01$); $I^2 = 94\%$ | | | |
| Test for subgroup differences (common effect): $Chi^2 = 164.74$, df = 4 ($P < 0.01$) | | | |
| Test for subgroup differences (random effects): $Chi^2 = 120.23$, df = 4 ($P < 0.01$) | | | |

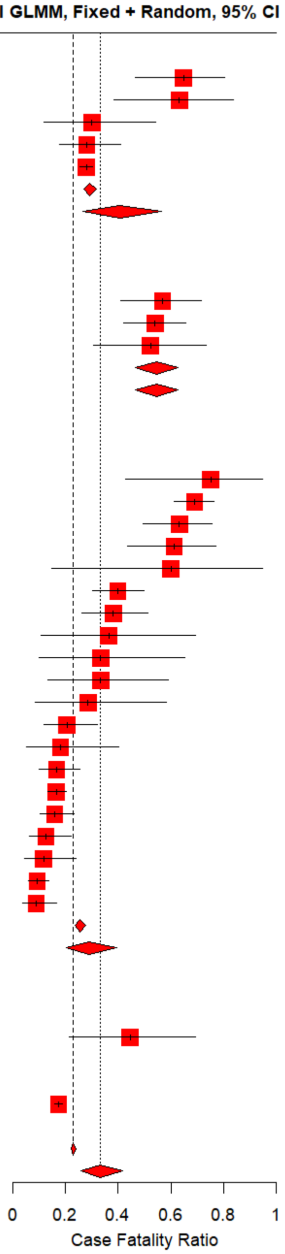

Figure C.5: Case fatality ratio (CFR) meta-analysis by lineage region (as shown in main text figure 3A), with individual study estimates. Plot includes additional lineage regions for which only one CFR estimate was available, for which meta-analysis was nor performed. Red squares indicate individual study estimates, with black lines representing a 95% binomial confidence interval; common and random effects are shown with red diamonds for subgroups and overall pooled estimate.

| Study or Subgroup | Events | Total | GLMM, Fixed + Random, 95% CI |
|---|---|---|---|
| **Benin** | | | |
| Yadouleton (2020) | 8 | 18 | 0.444 [0.215; 0.692] |
| **Guinea** | | | |
| ter Meulen (1998) | 4 | 12 | 0.333 [0.099; 0.651] |
| Bausch (2001) | 4 | 22 | 0.182 [0.052; 0.403] |
| **Total (common effect, 95% CI)** | **8** | **34** | **0.235 [0.122; 0.405]** |
| **Total (random effect, 95% CI)** | | | **0.235 [0.122; 0.405]** |
| Heterogeneity: $Tau^2 = 0$; $Chi^2 = 0.97$, df = 1 ($P = 0.33$); $I^2 = 0\%$ | | | |
| **Liberia** | | | |
| Jetoh (2022) | 41 | 103 | 0.398 [0.303; 0.499] |
| Monath (1973) | 4 | 11 | 0.364 [0.109; 0.692] |
| Hamblion (2018) | 4 | 14 | 0.286 [0.084; 0.581] |
| Frame (1989) | 23 | 246 | 0.093 [0.060; 0.137] |
| **Total (common effect, 95% CI)** | **72** | **374** | **0.193 [0.156; 0.236]** |
| **Total (random effect, 95% CI)** | | | **0.244 [0.121; 0.430]** |
| Heterogeneity: $Tau^2 = 0.5613$; $Chi^2 = 40.48$, df = 3 ($P < 0.01$); $I^2 = 93\%$ | | | |
| **Nigeria** | | | |
| Fisher-Hoch (1995) | 22 | 34 | 0.647 [0.465; 0.803] |
| Isere (2018) | 12 | 19 | 0.632 [0.384; 0.837] |
| Gomerep (2022) | 25 | 44 | 0.568 [0.410; 0.717] |
| Abdulkarim (2020) | 41 | 76 | 0.539 [0.421; 0.655] |
| White (1972) | 12 | 23 | 0.522 [0.306; 0.732] |
| Ajayi (2013) | 6 | 20 | 0.300 [0.119; 0.543] |
| Inegbenebor (2010) | 18 | 64 | 0.281 [0.176; 0.408] |
| Akpede (2019) | 362 | 1298 | 0.279 [0.255; 0.304] |
| Dalhat (2022) | 550 | 3162 | 0.174 [0.161; 0.188] |
| **Total (common effect, 95% CI)** | **1048** | **4740** | **0.221 [0.210; 0.233]** |
| **Total (random effect, 95% CI)** | | | **0.415 [0.298; 0.541]** |
| Heterogeneity: $Tau^2 = 0.5054$; $Chi^2 = 181.84$, df = 8 ($P < 0.01$); $I^2 = 96\%$ | | | |
| **Sierra Leone** | | | |
| Keane (1977) | 9 | 12 | 0.750 [0.428; 0.945] |
| Shaffer (2014) | 109 | 158 | 0.690 [0.612; 0.761] |
| Samuels (2021) | 36 | 57 | 0.632 [0.493; 0.756] |
| Dahmane (2014) | 22 | 36 | 0.611 [0.435; 0.769] |
| Njuguna (2022) | 3 | 5 | 0.600 [0.147; 0.947] |
| Fraser (1974) | 24 | 63 | 0.381 [0.261; 0.512] |
| Keane (1977) | 6 | 18 | 0.333 [0.133; 0.590] |
| Price (1988) | 14 | 68 | 0.206 [0.117; 0.321] |
| Keane (1977) | 16 | 96 | 0.167 [0.098; 0.256] |
| McCormick (1987a) | 73 | 441 | 0.166 [0.132; 0.204] |
| Keane (1977) | 22 | 138 | 0.159 [0.103; 0.231] |
| Price (1988) | 10 | 79 | 0.127 [0.062; 0.220] |
| Webb (1986) | 6 | 51 | 0.118 [0.044; 0.239] |
| Bonner (2007) | 8 | 90 | 0.089 [0.039; 0.168] |
| **Total (common effect, 95% CI)** | **358** | **1312** | **0.273 [0.249; 0.298]** |
| **Total (random effect, 95% CI)** | | | **0.309 [0.196; 0.450]** |
| Heterogeneity: $Tau^2 = 1.1849$; $Chi^2 = 236.24$, df = 13 ($P < 0.01$); $I^2 = 94\%$ | | | |
| **Total (common effect, 95% CI)** | **1494** | **6478** | **0.231 [0.221; 0.241]** |
| **Total (random effect, 95% CI)** | | | **0.331 [0.257; 0.415]** |
| Heterogeneity: $Tau^2 = 0.8555$; $Chi^2 = 482.78$, df = 29 ($P < 0.01$); $I^2 = 94\%$ | | | |
| Test for subgroup differences (common effect): $Chi^2 = 22.87$, df = 4 ($P < 0.01$) | | | |
| Test for subgroup differences (random effects): $Chi^2 = 5.35$, df = 4 ($P = 0.25$) | | | |

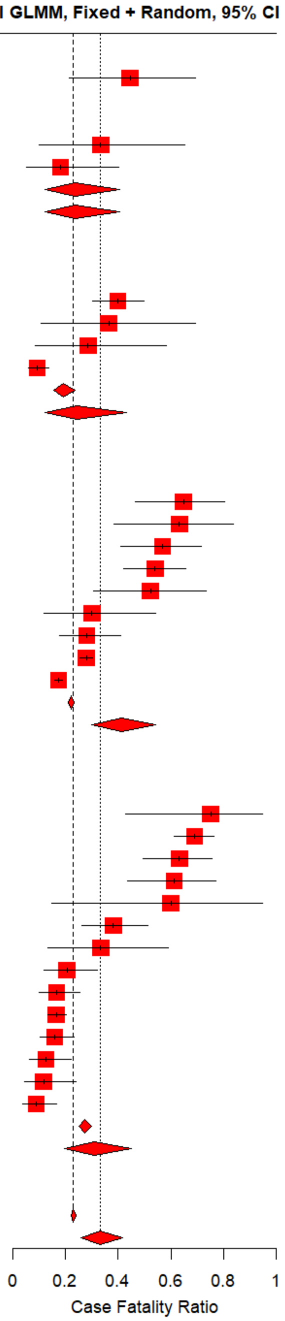

Figure C.6: Case fatality ratio (CFR) meta-analysis by country (as shown in main text figure 3C), with individual study estimates. Plot includes additional countries for which only one CFR estimate was available, for which meta-analysis was nor performed. Red squares indicate individual study estimates, with black lines representing a 95% binomial confidence interval; common and random effects are shown with red diamonds for subgroups and overall pooled estimate.

| Study or Subgroup | Events | Total | GLMM, Fixed + Random, 95% CI |
|---|---|---|---|
| **1970-1979** | | | |
| Keane (1977) | 9 | 12 | 0.750 [0.428; 0.945] |
| White (1972) | 12 | 23 | 0.522 [0.306; 0.732] |
| Fraser (1974) | 24 | 63 | 0.381 [0.261; 0.512] |
| Monath (1973) | 4 | 11 | 0.364 [0.109; 0.692] |
| Keane (1977) | 6 | 18 | 0.333 [0.133; 0.590] |
| Keane (1977) | 16 | 96 | 0.167 [0.098; 0.256] |
| McCormick (1987a) | 73 | 441 | 0.166 [0.132; 0.204] |
| Keane (1977) | 22 | 138 | 0.159 [0.103; 0.231] |
| Webb (1986) | 6 | 51 | 0.118 [0.044; 0.239] |
| **Total (common effect, 95% CI)** | **172** | **853** | **0.202 [0.176; 0.230]** |
| **Total (random effect, 95% CI)** | | | **0.281 [0.178; 0.413]** |
| Heterogeneity: $Tau^2 = 0.6391$; $Chi^2 = 49.13$, df = 8 ($P < 0.01$); $I^2 = 84\%$ | | | |
| **1980-1989** | | | |
| Fisher-Hoch (1995) | 22 | 34 | 0.647 [0.465; 0.803] |
| Price (1988) | 14 | 68 | 0.206 [0.117; 0.321] |
| Price (1988) | 10 | 79 | 0.127 [0.062; 0.220] |
| Frame (1989) | 23 | 246 | 0.093 [0.060; 0.137] |
| **Total (common effect, 95% CI)** | **69** | **427** | **0.162 [0.130; 0.200]** |
| **Total (random effect, 95% CI)** | | | **0.219 [0.086; 0.456]** |
| Heterogeneity: $Tau^2 = 1.1554$; $Chi^2 = 48.67$, df = 3 ($P < 0.01$); $I^2 = 94\%$ | | | |
| **1990-1999** | | | |
| Bausch (2001) | 4 | 22 | 0.182 [0.052; 0.403] |
| **2000-2009** | | | |
| Inegbenebor (2010) | 18 | 64 | 0.281 [0.176; 0.408] |
| Bonner (2007) | 8 | 90 | 0.089 [0.039; 0.168] |
| **Total (common effect, 95% CI)** | **26** | **154** | **0.169 [0.118; 0.236]** |
| **Total (random effect, 95% CI)** | | | **0.165 [0.069; 0.343]** |
| Heterogeneity: $Tau^2 = 0.3913$; $Chi^2 = 9$, df = 1 ($P < 0.01$); $I^2 = 89\%$ | | | |
| **2010-2019** | | | |
| Shaffer (2014) | 109 | 158 | 0.690 [0.612; 0.761] |
| Isere (2018) | 12 | 19 | 0.632 [0.384; 0.837] |
| Samuels (2021) | 36 | 57 | 0.632 [0.493; 0.756] |
| Dahmane (2014) | 22 | 36 | 0.611 [0.435; 0.769] |
| Njuguna (2022) | 3 | 5 | 0.600 [0.147; 0.947] |
| Gomerep (2022) | 25 | 44 | 0.568 [0.410; 0.717] |
| Abdulkarim (2020) | 41 | 76 | 0.539 [0.421; 0.655] |
| Yadouleton (2020) | 8 | 18 | 0.444 [0.215; 0.692] |
| Ajayi (2013) | 6 | 20 | 0.300 [0.119; 0.543] |
| Hamblion (2018) | 4 | 14 | 0.286 [0.084; 0.581] |
| Akpede (2019) | 362 | 1298 | 0.279 [0.255; 0.304] |
| **Total (common effect, 95% CI)** | **628** | **1745** | **0.360 [0.338; 0.383]** |
| **Total (random effect, 95% CI)** | | | **0.509 [0.410; 0.607]** |
| Heterogeneity: $Tau^2 = 0.3146$; $Chi^2 = 151.83$, df = 10 ($P < 0.01$); $I^2 = 93\%$ | | | |
| **2020-Present** | | | |
| Jetoh (2022) | 41 | 103 | 0.398 [0.303; 0.499] |
| Dalhat (2022) | 550 | 3162 | 0.174 [0.161; 0.188] |
| **Total (common effect, 95% CI)** | **591** | **3265** | **0.181 [0.168; 0.195]** |
| **Total (random effect, 95% CI)** | | | **0.265 [0.140; 0.443]** |
| Heterogeneity: $Tau^2 = 0.3030$; $Chi^2 = 30.66$, df = 1 ($P < 0.01$); $I^2 = 97\%$ | | | |
| **Unspecified** | | | |
| ter Meulen (1998) | 4 | 12 | 0.333 [0.099; 0.651] |
| **Total (common effect, 95% CI)** | **1494** | **6478** | **0.231 [0.221; 0.241]** |
| **Total (random effect, 95% CI)** | | | **0.331 [0.257; 0.415]** |
| Heterogeneity: $Tau^2 = 0.8555$; $Chi^2 = 482.78$, df = 29 ($P < 0.01$); $I^2 = 94\%$ | | | |
| Test for subgroup differences (common effect): $Chi^2 = 221.43$, df = 6 ($P < 0.01$) | | | |
| Test for subgroup differences (random effects): $Chi^2 = 20.47$, df = 6 ($P < 0.01$) | | | |

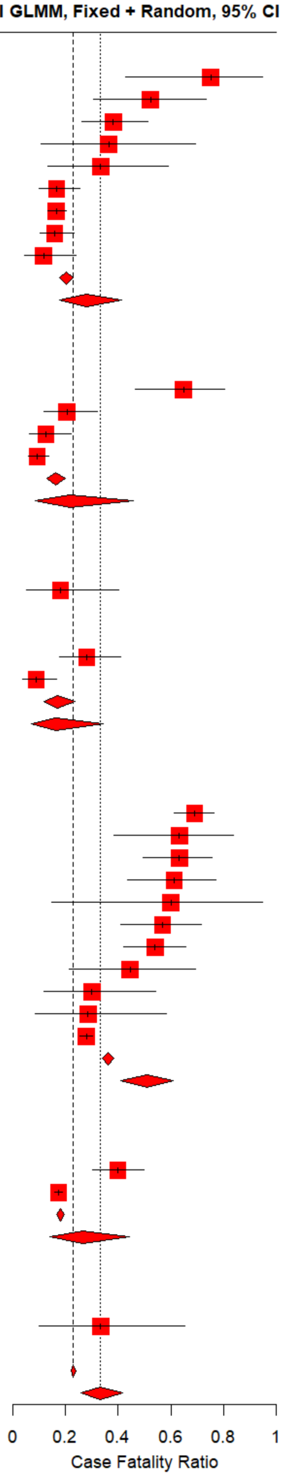

Figure C.7: Case fatality ratio (CFR) meta-analysis by decade, with individual study estimates. Red squares indicate individual study estimates, with black lines representing a 95% binomial confidence interval; common and random effects are shown with red diamonds for subgroups and overall pooled estimate.

| Study or Subgroup | Events | Total | GLMM, Fixed + Random, 95% CI |
|---|---|---|---|
| **Reported Cases < 30** | | | |
| Keane (1977) | 9 | 12 | 0.750 [0.428; 0.945] |
| Isere (2018) | 12 | 19 | 0.632 [0.384; 0.837] |
| Njuguna (2022) | 3 | 5 | 0.600 [0.147; 0.947] |
| White (1972) | 12 | 23 | 0.522 [0.306; 0.732] |
| Yadouleton (2020) | 8 | 18 | 0.444 [0.215; 0.692] |
| Monath (1973) | 4 | 11 | 0.364 [0.109; 0.692] |
| ter Meulen (1998) | 4 | 12 | 0.333 [0.099; 0.651] |
| Keane (1977) | 6 | 18 | 0.333 [0.133; 0.590] |
| Ajayi (2013) | 6 | 20 | 0.300 [0.119; 0.543] |
| Hamblion (2018) | 4 | 14 | 0.286 [0.084; 0.581] |
| Bausch (2001) | 4 | 22 | 0.182 [0.052; 0.403] |
| **Total (common effect, 95% CI)** | **72** | **174** | **0.414 [0.343; 0.488]** |
| **Total (random effect, 95% CI)** | | | **0.415 [0.317; 0.520]** |
| Heterogeneity: Tau$^2$ = 0.2233; Chi$^2$ = 17.26, df = 10 (P = 0.07); $I^2$ = 42% | | | |
| **Reported Cases = 30-99** | | | |
| Fisher-Hoch (1995) | 22 | 34 | 0.647 [0.465; 0.803] |
| Samuels (2021) | 36 | 57 | 0.632 [0.493; 0.756] |
| Dahmane (2014) | 22 | 36 | 0.611 [0.435; 0.769] |
| Gomerep (2022) | 25 | 44 | 0.568 [0.410; 0.717] |
| Abdulkarim (2020) | 41 | 76 | 0.539 [0.421; 0.655] |
| Fraser (1974) | 24 | 63 | 0.381 [0.261; 0.512] |
| Inegbenebor (2010) | 18 | 64 | 0.281 [0.176; 0.408] |
| Price (1988) | 14 | 68 | 0.206 [0.117; 0.321] |
| Keane (1977) | 16 | 96 | 0.167 [0.098; 0.256] |
| Price (1988) | 10 | 79 | 0.127 [0.062; 0.220] |
| Webb (1986) | 6 | 51 | 0.118 [0.044; 0.239] |
| Bonner (2007) | 8 | 90 | 0.089 [0.039; 0.168] |
| **Total (common effect, 95% CI)** | **242** | **758** | **0.319 [0.287; 0.353]** |
| **Total (random effect, 95% CI)** | | | **0.327 [0.210; 0.471]** |
| Heterogeneity: Tau$^2$ = 1.0424; Chi$^2$ = 126.42, df = 11 (P < 0.01); $I^2$ = 91% | | | |
| **Reported Cases = 100-299** | | | |
| Shaffer (2014) | 109 | 158 | 0.690 [0.612; 0.761] |
| Jetoh (2022) | 41 | 103 | 0.398 [0.303; 0.499] |
| Keane (1977) | 22 | 138 | 0.159 [0.103; 0.231] |
| Frame (1989) | 23 | 246 | 0.093 [0.060; 0.137] |
| **Total (common effect, 95% CI)** | **195** | **645** | **0.302 [0.268; 0.339]** |
| **Total (random effect, 95% CI)** | | | **0.291 [0.113; 0.570]** |
| Heterogeneity: Tau$^2$ = 1.3854; Chi$^2$ = 146.14, df = 3 (P < 0.01); $I^2$ = 98% | | | |
| **Reported Cases = 300-999** | | | |
| McCormick (1987a) | 73 | 441 | 0.166 [0.132; 0.204] |
| **Reported Cases >= 1000** | | | |
| Akpede (2019) | 362 | 1298 | 0.279 [0.255; 0.304] |
| Dalhat (2022) | 550 | 3162 | 0.174 [0.161; 0.188] |
| **Total (common effect, 95% CI)** | **912** | **4460** | **0.204 [0.193; 0.217]** |
| **Total (random effect, 95% CI)** | | | **0.221 [0.157; 0.302]** |
| Heterogeneity: Tau$^2$ = 0.0893; Chi$^2$ = 61.28, df = 1 (P < 0.01); $I^2$ = 98% | | | |
| **Total (common effect, 95% CI)** | **1494** | **6478** | **0.231 [0.221; 0.241]** |
| **Total (random effect, 95% CI)** | | | **0.331 [0.257; 0.415]** |
| Heterogeneity: Tau$^2$ = 0.8555; Chi$^2$ = 482.78, df = 29 (P < 0.01); $I^2$ = 94% | | | |
| Test for subgroup differences (common effect): Chi$^2$ = 109.49, df = 4 (P < 0.01) | | | |
| Test for subgroup differences (random effects): Chi$^2$ = 29.04, df = 4 (P < 0.01) | | | |

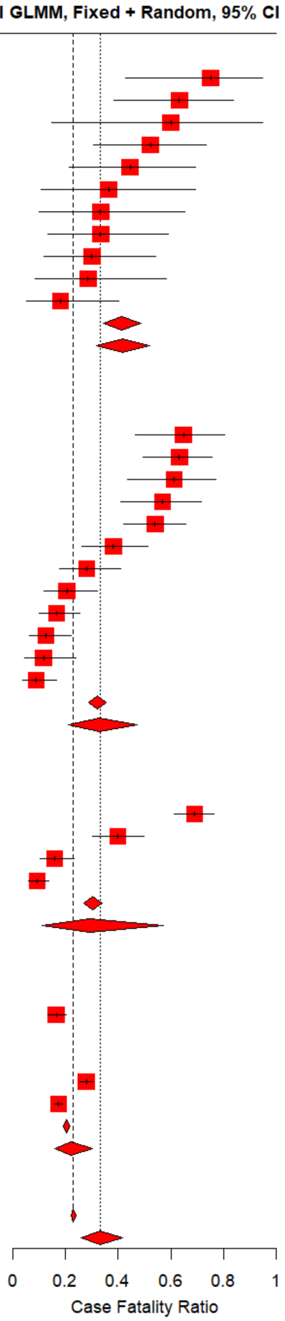

Figure C.8: Case fatality ratio (CFR) meta-analysis by number of reported cases (as shown in main text figure 3D), with individual study estimates. Plot includes additional ranges for which only one CFR estimate was available, for which meta-analysis was nor performed. Red squares indicate individual study estimates, with black lines representing a 95% binomial confidence interval; common and random effects are shown with red diamonds for subgroups and overall pooled estimate.

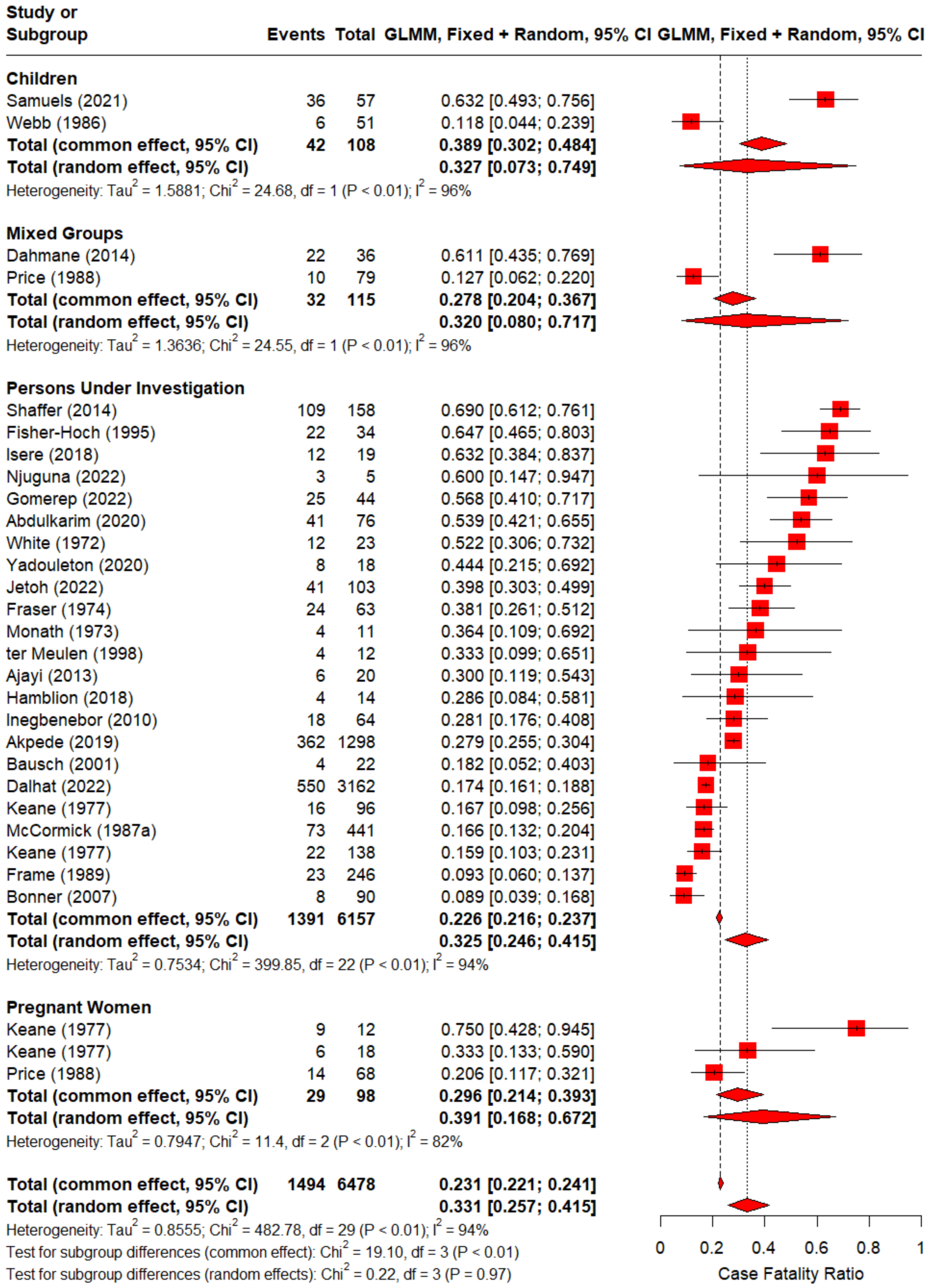

Figure C.9: Case fatality ratio (CFR) meta-analysis by population group (as shown in main text figure 3E), with individual study estimates. Red squares indicate individual study estimates, with black lines representing a 95% binomial confidence interval; common and random effects are shown with red diamonds for subgroups and overall pooled estimate.

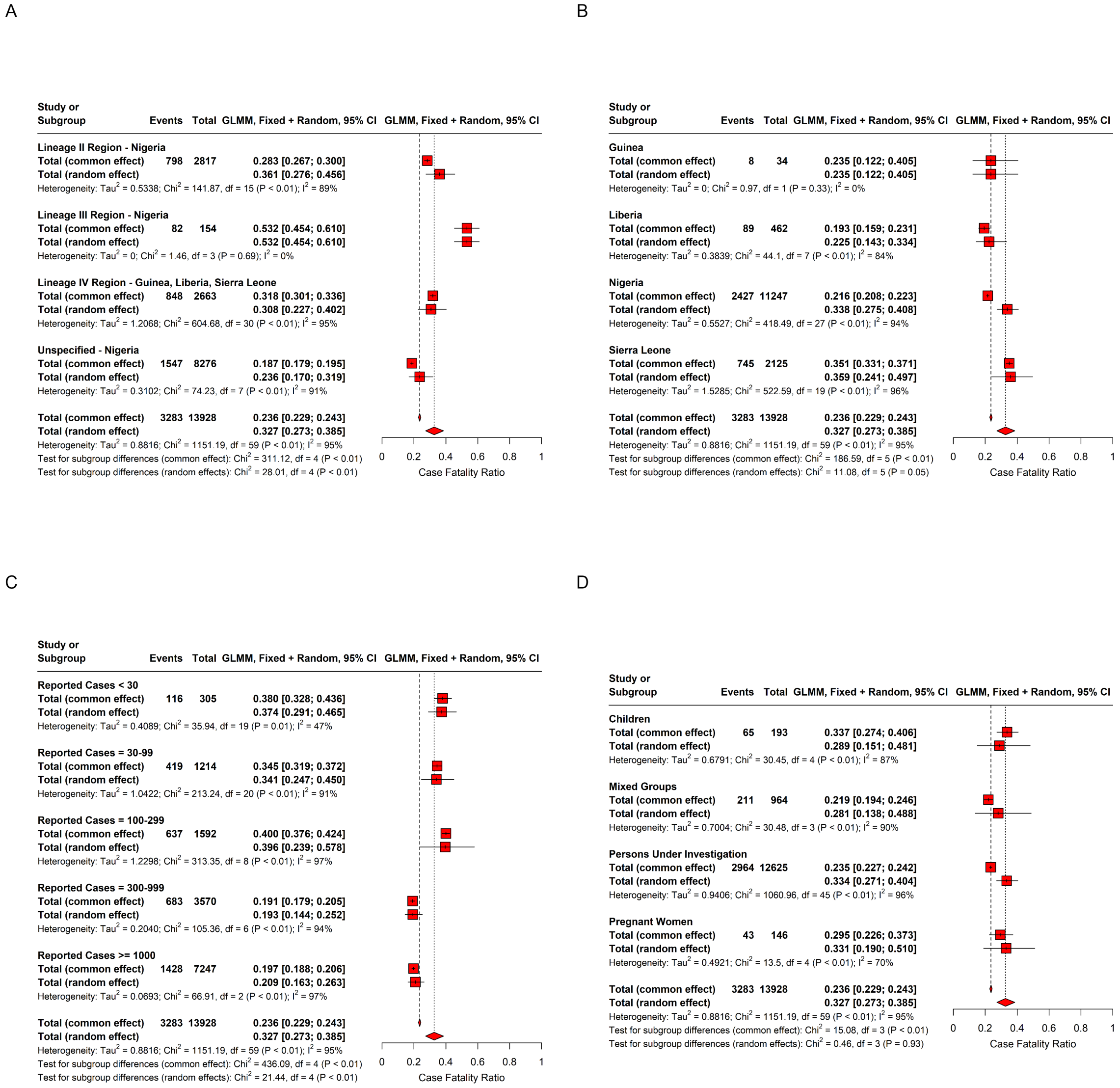

Figure C.10: Case fatality ratio (CFR) meta-analyses with only 'known duplicate' data removed, by (A) lineage region, (B) country, (C) number of reported cases, and (D) population group under study. CFR estimates are considered known duplicates if they refer to the same underlying data (e.g. naive and adjusted CFRs, a subset of reported cases). Here, 'assumed duplicate' data are included, for example CFRs with overlapping locations or time periods, in contrast with the stricter de-duplication carried out in the main text (described in appendix A.3). Red squares indicate common and random effects for each subgroup, and red diamonds overall common and random effects.

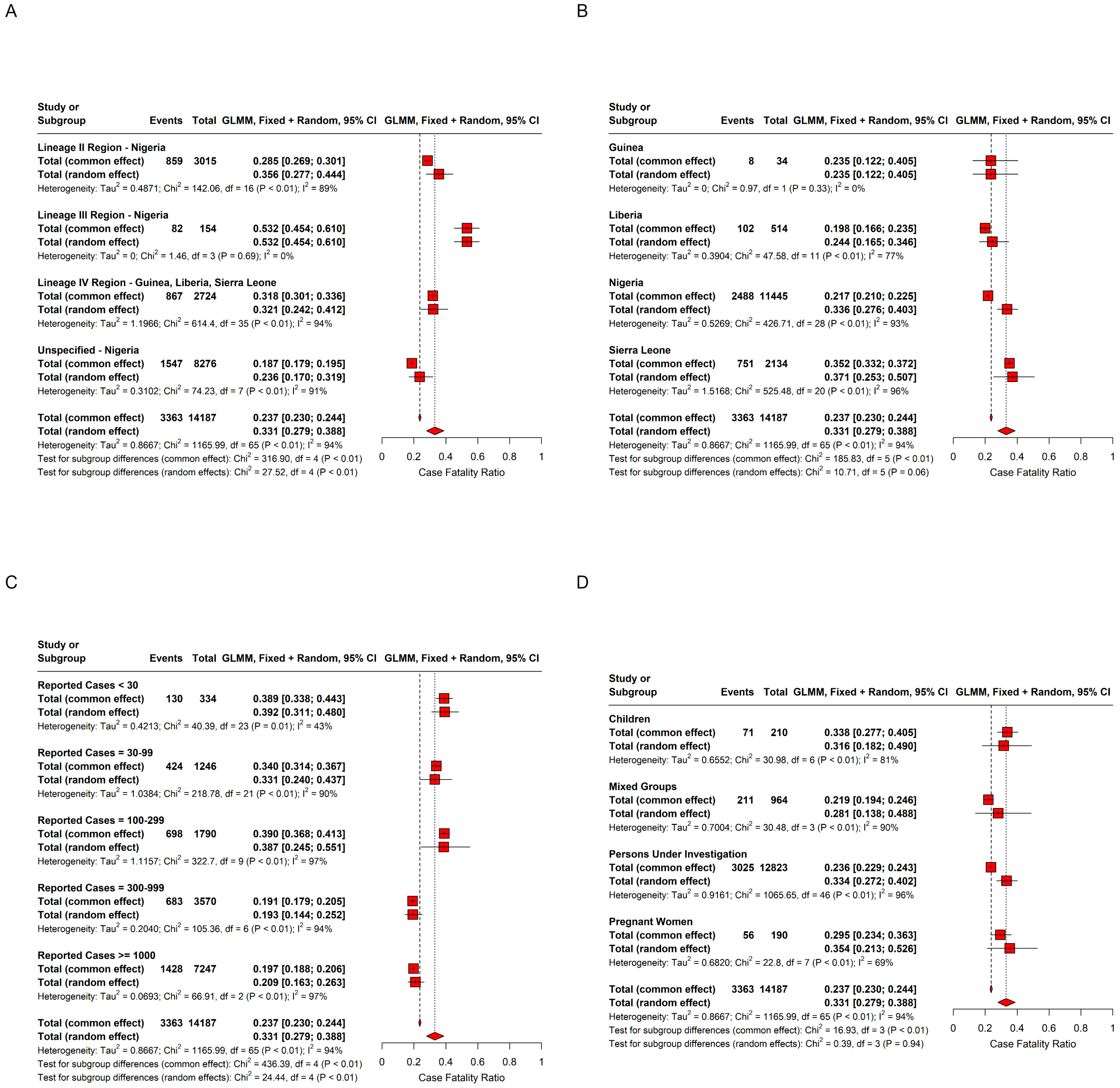

Figure C.11: Case fatality ratio (CFR) meta-analyses of all extracted data, by (A) lineage region, (B) country, (C) number of reported cases, and (D) population group under study. Here, no de-duplication is carried out and both 'known duplicates' (with the same underlying data, e.g. naive and adjusted CFRs, a subset of reported cases) and 'assumed duplicates' (with overlapping locations or time periods) are included, in contrast with the stricter de-duplication carried out in the main text (described in appendix A.3). Red squares indicate common and random effects for each subgroup, and red diamonds overall common and random effects.

### D epireview

We developed an R package called *epireview* that provides a central location to host and access the extracted data for the nine priority pathogens, allows for submissions of outbreak, model, and parameter data from new peer-reviewed papers via pull requests, and includes functions to produce the tables and figures included in this paper and update them with any additional data. This package will be updated as the overall project by the Pathogen Epidemiology Review Group (PERG) continues to extract modelling parameters for the rest of the nine priority pathogens as defined by WHO.

There are several vignettes available:

- A vignette that lists the options for each outbreak, model, or parameter field and describes how to access them using a function in the package.
- A vignette to explain the process of updating the database with new article, outbreak, model, or parameter data.

### E PRISMA 2020 Checklists

| Section & Topic | Item # | Checklist item | Reported (Yes/No) |
|---|---|---|---|
| **Title** | | | |
| Title | 1 | Identify the report as a systematic review. | Yes |
| **Background** | | | |
| Objectives | 2 | Provide an explicit statement of the main objective(s) or question(s) the review addresses. | Yes |
| **Methods** | | | |
| Eligibility criteria | 3 | Specify the inclusion and exclusion criteria for the review. | Yes |
| Information sources | 4 | Specify the information sources (e.g. databases, registers) used to identify studies and the date when each was last searched. | Yes |
| Risk of bias | 5 | Specify the methods used to assess risk of bias in the included studies. | Yes |
| Synthesis of results | 6 | Specify the methods used to present and synthesise results. | Yes |
| **Results** | | | |
| Included studies | 7 | Give the total number of included studies and participants and summarise relevant characteristics of studies. | Yes |
| Synthesis of results | 8 | Present results for main outcomes, preferably indicating the number of included studies and participants for each. If meta-analysis was done, report the summary estimate and confidence/credible interval. If comparing groups, indicate the direction of the effect (i.e. which group is favoured). | Yes |
| **Discussion** | | | |
| Limitations of evidence | 9 | Provide a brief summary of the limitations of the evidence included in the review (e.g. study risk of bias, inconsistency and imprecision). | Yes |
| Interpretation | 10 | Provide a general interpretation of the results and important implications. | Yes |
| **Other** | | | |
| Funding | 11 | Specify the primary source of funding for the review. | Yes |
| Registration | 12 | Provide the register name and registration number. | Yes |

Table E.13: PRISMA 2020 Abstracts Checklist. ([5])

| Section & Topic | Item # | Checklist item | Location where item is reported |
|---|---|---|---|
| **Title** | | | |
| Title | 1 | Identify the report as a systematic review. | page 1 |
| **Abstract** | | | |
| Abstract | 2 | See the PRISMA 2020 for Abstracts checklist. | Table E.13 |
| **Introduction** | | | |
| Rationale | 3 | Describe the rationale for the review in the context of existing knowledge. | page 2 |
| Objectives | 4 | Provide an explicit statement of the objective(s) or question(s) the review addresses. | page 2/3 |
| **Methods** | | | |
| Eligibility criteria | 5 | Specify the inclusion and exclusion criteria for the review and how studies were grouped for the syntheses. | page 3 |
| Information sources | 6 | Specify all databases, registers, websites, organisations, reference lists and other sources searched or consulted to identify studies. Specify the date when each source was last searched or consulted. | page 3 |
| Search strategy | 7 | Present the full search strategies for all databases, registers and websites, including any filters and limits used. | page 3 + Figure 1 |

Table E.14: PRISMA 2020 Checklist. ([5])

| Section & Topic | Item # | Checklist item | Location where item is reported |
|---|---|---|---|
| Selection process | 8 | Specify the methods used to decide whether a study met the inclusion criteria of the review, including how many reviewers screened each record and each report retrieved, whether they worked independently, and if applicable, details of automation tools used in the process. | page 3 |
| Data collection process | 9 | Specify the methods used to collect data from reports, including how many reviewers collected data from each report, whether they worked independently, any processes for obtaining or confirming data from study investigators, and if applicable, details of automation tools used in the process. | page 3/4 |
| Data items | 10a | List and define all outcomes for which data were sought. Specify whether all results that were compatible with each outcome domain in each study were sought (e.g. for all measures, time points, analyses), and if not, the methods used to decide which results to collect. | page 3/4 |
| | 10b | List and define all other variables for which data were sought (e.g. participant and intervention characteristics, funding sources). Describe any assumptions made about any missing or unclear information. | page 3/4 |
| Study risk of bias assessment | 11 | Specify the methods used to assess risk of bias in the included studies, including details of the tool(s) used, how many reviewers assessed each study and whether they worked independently, and if applicable, details of automation tools used in the process. | page 4 |
| Effect measures | 12 | Specify for each outcome the effect measure(s) (e.g. risk ratio, mean difference) used in the synthesis or presentation of results. | page 3/4 |
| Synthesis methods | 13a | Describe the processes used to decide which studies were eligible for each synthesis (e.g. tabulating the study intervention characteristics and comparing against the planned groups for each synthesis (item #5)). | page 4 |
| | 13b | Describe any methods required to prepare the data for presentation or synthesis, such as handling of missing summary statistics, or data conversions. | page 4 |
| | 13c | Describe any methods used to tabulate or visually display results of individual studies and syntheses. | page 4 |
| | 13d | Describe any methods used to synthesize results and provide a rationale for the choice(s). If meta-analysis was performed, describe the model(s), method(s) to identify the presence and extent of statistical heterogeneity, and software package(s) used. | page 4 |
| | 13e | Describe any methods used to explore possible causes of heterogeneity among study results (e.g. subgroup analysis, meta-regression). | - |
| | 13f | Describe any sensitivity analyses conducted to assess robustness of the synthesized results. | - |
| Reporting bias assessment | 14 | Describe any methods used to assess risk of bias due to missing results in a synthesis (arising from reporting biases). | page 4 |
| Certainty assessment | 15 | Describe any methods used to assess certainty (or confidence) in the body of evidence for an outcome. | page 4 |
| **Results** | | | |
| Study selection | 16a | Describe the results of the search and selection process, from the number of records identified in the search to the number of studies included in the review, ideally using a flow diagram. | page 4 |
| | 16b | Cite studies that might appear to meet the inclusion criteria, but which were excluded, and explain why they were excluded. | Table G.16 |
| Study characteristics | 17 | Cite each included study and present its characteristics. | pages 4-13 |
| Risk of bias in studies | 18 | Present assessments of risk of bias for each included study. | page 7 |
| Results of individual studies | 19 | For all outcomes, present, for each study: (a) summary statistics for each group (where appropriate) and (b) an effect estimate and its precision (e.g. confidence/credible interval), ideally using structured tables or plots. | pages 4-13 |
| Results of syntheses | 20a | For each synthesis, briefly summarise the characteristics and risk of bias among contributing studies. | page 6/7 |
| | 20b | Present results of all statistical syntheses conducted. If meta-analysis was done, present for each the summary estimate and its precision (e.g. confidence/credible interval) and measures of statistical heterogeneity. If comparing groups, describe the direction of the effect. | page 4-13 |
| | 20c | Present results of all investigations of possible causes of heterogeneity among study results. | page 4-13 |
| | 20d | Present results of all sensitivity analyses conducted to assess the robustness of the synthesized results. | page 4-13 |
| Reporting biases | 21 | Present assessments of risk of bias due to missing results (arising from reporting biases) for each synthesis assessed. | page 7 |

Table E.14: PRISMA 2020 Checklist. ([5])

| Section & Topic | Item # | Checklist item | Location where item is reported |
|---|---|---|---|
| Certainty of evidence | 22 | Present assessments of certainty (or confidence) in the body of evidence for each outcome assessed. | pages 4-13 |
| **Discussion** | | | |
| Discussion | 23a | Provide a general interpretation of the results in the context of other evidence. | page 14/15 |
| | 23b | Discuss any limitations of the evidence included in the review. | page 14-15 |
| | 23c | Discuss any limitations of the review processes used. | page 14-15 |
| | 23d | Discuss implications of the results for practice, policy, and future research. | page 14-15 |
| **Other Information** | | | |
| Registration and protocol | 24a | Provide registration information for the review, including register name and registration number, or state that the review was not registered. | page 14 |
| | 24b | Indicate where the review protocol can be accessed, or state that a protocol was not prepared. | page 16 |
| | 24c | Describe and explain any amendments to information provided at registration or in the protocol. | n/a |
| Support | 25 | Describe sources of financial or non-financial support for the review, and the role of the funders or sponsors in the review. | page 15/16 |
| Competing interests | 26 | Declare any competing interests of review authors. | page 16 |
| Availability of data, code and other materials | 27 | Report which of the following are publicly available and where they can be found: template data collection forms; data extracted from included studies; data used for all analyses; analytic code; any other materials used in the review. | page 16 |

Table E.14: PRISMA 2020 Checklist. ([5])

### F Other Systematic Reviews

Systematic reviews were excluded from our analysis, but the systematic reviews that were identified in the database search and used for validation purposes are presented in Table F.15.

| Study | Title | Journal | DOI |
|---|---|---|---|
| Monath 1975 | Lassa Fever - Review of Epidemiology and Epizootiology | Bulletin of the World Health Organization | |
| Wang 2021 | The reproductive number of Lassa fever: a systematic review. | Journal of travel medicine | 10.1093/jtm/taab029 |
| Kenmoe 2020 | Systematic review and meta-analysis of the epidemiology of Lassa virus in humans, rodents and other mammals in sub-Saharan Africa. | PLoS neglected tropical diseases | 10.1371/journal.pntd.0008589 |
| Merson 2021 | Clinical characterization of Lassa fever: A systematic review of clinical reports and research to inform clinical trial design. | PLoS neglected tropical diseases | 10.1371/journal.pntd.0009788 |
| Eberhardt 2019 | Ribavirin for the treatment of Lassa fever: A systematic review and meta-analysis. | International journal of infectious diseases : IJID : official publication of the International Society for Infectious Diseases | 10.1016/j.ijid.2019.07.015 |
| Agbonlahor 2021 | 52 Years of Lassa Fever Outbreaks in Nigeria, 1969-2020: An Epidemiologic Analysis of the Temporal and Spatial Trends. | The American journal of tropical medicine and hygiene | 10.4269/ajtmh.20-1160 |
| Kayem 2020 | Lassa fever in pregnancy: a systematic review and meta-analysis. | Transactions of the Royal Society of Tropical Medicine and Hygiene | 10.1093/trstmh/traa011 |
| Wolf 2020 | Fifty years of imported Lassa fever: a systematic review of primary and secondary cases. | Journal of travel medicine | 10.1093/jtm/taaa035 |
| Simons 2022 | Lassa fever cases suffer from severe underreporting based on reported fatalities. | International health | 10.1093/inthealth/ihac076 |
| Alli 2021 | Management of Lassa Fever: A Current Update | CUREUS JOURNAL OF MEDICAL SCIENCE | 10.7759/cureus.14797 |
| Belhadi 2022 | The number of cases, mortality and treatments of viral hemorrhagic fevers: A systematic review | PLOS NEGLECTED TROPICAL DISEASES | 10.1371/journal.pntd.0010889 |
| Garry 2022 | Lassa fever — the road ahead | Nature Reviews Microbiology | 10.1038/s41579-022-00789-8 |
| Balogun 2021 | Lassa Fever: An Evolving Emergency in West Africa | The American journal of tropical medicine and hygiene | 10.4269/ajtmh.20-0487 |

Table F.15: Other systematic reviews

### G Excluded Studies

We list all excluded studies with reasons for exclusion in Table G.16

| Study | Title | Journal | Notes |
|---|---|---|---|
| Weber 2016 | Emerging infectious diseases: Focus on infection control issues for novel coronaviruses (Severe Acute Respiratory Syndrome-CoV and Middle East Respiratory Syndrome-CoV), hemorrhagic fever viruses (Lassa and Ebola), and highly pathogenic avian influenza viruses, A(H5N1) and A(H7N9) | American Journal of Infection Control | Exclusion reason: 6 not original estimates/ primary data; |
| Weber 2001 | Risks and prevention of nosocomial transmission of rare zoonotic diseases | Clinical Infectious Diseases | Exclusion reason: 6 not original estimates/ primary data; |
| Webber 2009 | Diseases transmitted via body fluids | | Exclusion reason: 7 not peer reviewed; Natsuko Imai (2019-10-26 03:30:40)(Select): book, exlude; |
| Tomori 1999 | Options for the control of rodent-borne viral diseases in West Africa | | Exclusion reason: 7 not peer reviewed; Natsuko Imai (2019-10-26 03:30:22)(Select): conference proceedings exclude; |
| Tomori 1988 | Lassa Fever and Other Viral Hemorrhagic Disorders | Impact of Science on Society | Exclusion reason: 6 not original estimates/ primary data; Natsuko Imai (2019-10-24 00:58:09)(Select): paper copy; |
| Tobin 2014 | Risk factors for Lassa fever in endemic communities of Edo State, Nigeria | International Journal of Infectious Diseases | Exclusion reason: 7 not peer reviewed; David Jorgensen (2019-06-22 00:21:31)(Select): poster; |
| Meulen 2001 | Lassa fever in Sierra Leone: UN peacekeepers are at risk | Tropical Medicine & International Health | Exclusion reason: 4 case report/study; Natsuko Imai (2019-03-28 03:31:48)(Screen) |
| Meulen 2000 | Lassa fever: immuno-epidemiological approach to the study of an endemic viral haemorrhagic fever | Medecine tropicale : revue du Corps de sante colonial | Exclusion reason: 6 not original estimates/ primary data; Natsuko Imai (2019-10-24 00:56:45)(Select): paper copy; |
| Tambo 2018 | Re-emerging Lassa fever outbreaks in Nigeria: Re-enforcing "One Health" community surveillance and emergency response practice | Infectious Diseases of Poverty | Exclusion reason: 6 not original estimates/ primary data; |
| Swaan 2002 | Management of viral haemorrhagic fever in the Netherlands | Euro surveillance : bulletin Europeen sur les maladies transmissibles = European communicable disease bulletin | Exclusion reason: 5 no report of parameters ; |
| Sureau 1989 | Recent findings on the African viral haemorrhagic fevers | | Exclusion reason: 2 not English; Natsuko Imai (2019-10-28 20:17:11)(Select): exclude is in French; Natsuko Imai (2019-10-26 03:30:03)(Select): waiting on FT, from marburg group; |
| Stephenson 1984 | Effect of Environmental-Factors on Aerosol-Induced Lassa Virus-Infection | Journal of Medical Virology | Exclusion reason: 5 no report of parameters ; |

Table G.16: Excluded studies at full text review with exclusion reason

| Study | Title | Journal | Notes |
|---|---|---|---|
| Speed 1996 | Viral haemorrhagic fevers: Current status, future threats | Medical Journal of Australia | Exclusion reason: 6 not original estimates/ primary data; |
| Sogoba 2012 | Lassa Fever in West Africa: Evidence for an Expanded Region of Endemicity | Zoonoses and Public Health | Exclusion reason: 1 duplicate; |
| Smith 1979 | Epidemiological Aspect of the 1976 Pankshin Lassa Fever Outbreak | Nigerian Medical Journal | Exclusion reason: 6 not original estimates/ primary data; David Jorgensen (2019-03-29 04:18:37)(Screen): https://www.journalofinfection.com/article/S0163-4453(79)90977-0/abstract; |
| Simpson 1980 | Nasty Viruses - Lassa, Marburg, and Ebola | British Journal of Hospital Medicine | Exclusion reason: 6 not original estimates/ primary data; |
| Simpson 1978 | Viral Hemorrhagic Fevers of Man | Bulletin of the World Health Organization | Exclusion reason: 6 not original estimates/ primary data; Natsuko Imai (2019-03-29 04:25:00)(Screen): This article reviews the current state of knowledge on the viral haemorrhagic fevers that infect man, namely smallpox, chikungunya fever, dengue fever, Rift Valley fever, yellow fever, Crimean haemorrhagic fever, Kyasanur Forest disease, Omsk haemorrhagic fever, Argentinian haemorrhagic fever (Junin virus), Bolivian haemorrhagic fever (Machupo virus), Lassa fever, haemorrhagic fever with renal syndrome, and Marburg and Ebola virus diseases.; |
| Shlaeffer 1988 | Lassa Fever First Case Diagnosed in Israel | Harefuah | Exclusion reason: 2 not English; David Jorgensen (2019-06-22 00:06:09)(Select): Hebrew ; |
| Shelokov 1971 | New Viruses Associated with the Hemorrhagic Fever Syndrome | Annals of the New York Academy of Sciences | Exclusion reason: 3 wrong pathogen / epi not main focus; |
| Seriki 2018 | Descriptive analysis of Lassa fever outbreak data in Edo State, Nigeria, 2016 | International Journal of Infectious Diseases | Exclusion reason: 7 not peer reviewed; David Jorgensen (2019-06-22 00:01:19)(Select): poster abstract ; |
| Seah 1978 | Lassa, Marburg and Ebola - Newly Described African Fevers | Canadian Medical Association Journal | Exclusion reason: 6 not original estimates/ primary data; |
| Schoepp 2014 | Undiagnosed Acute Viral Febrile Illnesses, Sierra Leone | Emerging Infectious Diseases | Exclusion reason: 5 no report of parameters ; David Jorgensen (2019-10-24 01:38:17)(Select): not a representative sample, only taken from those with LASV symptoms; |
| Schmitz 2002 | Monitoring of clinical and laboratory data in two cases of imported Lassa fever | Microbes and Infection | Exclusion reason: 4 case report/study; |
| Schlaeffer 1988 | Evidence against High Contagiousness of Lassa Fever | Transactions of the Royal Society of Tropical Medicine and Hygiene | Exclusion reason: 4 case report/study; Natsuko Imai (2019-03-28 04:06:33)(Screen): conference abstract; |
| Safronetz 2017 | Annual Incidence of Lassa Virus Infection in Southern Mali | American Journal of Tropical Medicine and Hygiene | Exclusion reason: 5 no report of parameters ; |
| Safronetz 2010 | Detection of Lassa Virus, Mali | Emerging Infectious Diseases | Exclusion reason: 5 no report of parameters ; |
| Robbins 1977 | Algorithms in Diagnosis and Management of Exotic Diseases .19. Major Tropical Viral-Infections - Smallpox, Yellow-Fever, and Lassa Fever | Journal of Infectious Diseases | Exclusion reason: 6 not original estimates/ primary data; Natsuko Imai (2019-03-28 03:10:15)(Screen): |

Table G.16: Excluded studies at full text review with exclusion reason

| Study | Title | Journal | Notes |
|---|---|---|---|
| Richmond 2004 | Lassa fever: epidemiology, clinical features, and social consequences (vol 327, pg 1271, 2003) | British Medical Journal | Exclusion reason: 1 duplicate; Natsuko Imai (2019-03-28 04:00:25)(Screen): potentially duplicate?; |
| Richmond 2004 | Lassa fever: epidemiology, clinical features, and social consequences (vol 327, pg 1271, 2003) | British Medical Journal | Exclusion reason: 6 not original estimates/ primary data; Natsuko Imai (2019-10-03 00:23:45)(Select): including review as it summarises seroprevalence from other papers for easy ref; |
| Raeburn 1976 | Lassa fever | Nursing times | Exclusion reason: 7 not peer reviewed; |
| | Lassa fever 1982 | British medical journal (Clinical research ed.) | Exclusion reason: 4 case report/study; Natsuko Imai (2019-03-29 01:41:28)(Screen): https://www.bmj.com/content/287/6384/48; |
| Pontremoli 2018 | Arenavirus genomics: novel insights into viral diversity, origin, and evolution | Current Opinion in Virology | Exclusion reason: 5 no report of parameters ; |
| Pigott 2017 | Local, national, and regional viral haemorrhagic fever pandemic potential in Africa: a multistage analysis | Lancet | Exclusion reason: 6 not original estimates/ primary data; |
| Pigott 2005 | Hemorrhagic fever viruses | Critical Care Clinics | Exclusion reason: 6 not original estimates/ primary data; Natsuko Imai (2019-10-23 23:32:49)(Select): review ; Natsuko Imai (2019-10-23 23:32:28)(Select): paper copy; |
| Peterson 2014 | Mapping Transmission Risk of Lassa Fever in West Africa: The Importance of Quality Control, Sampling Bias, and Error Weighting | Plos One | Exclusion reason: 5 no report of parameters ; |
| Peters 1995 | Arena virus diseases | | Exclusion reason: 7 not peer reviewed; Natsuko Imai (2019-10-26 03:08:11)(Select): exclude book; |
| Patassi 2017 | Emergence of Lassa Fever Disease in Northern Togo: Report of Two Cases in Oti District in 2016 | Case Reports in Infectious Diseases | Exclusion reason: 4 case report/study; |
| Panning 2010 | Laboratory Diagnosis of Lassa Fever, Liberia | Emerging Infectious Diseases | Exclusion reason: 5 no report of parameters ; |
| Zwizwai 2016 | Infection disease surveillance update | Lancet Infectious Diseases | Exclusion reason: 7 not peer reviewed; David Jorgensen (2019-03-29 04:30:31)(Screen): Monthly bulletin ; |
| Yun 2012 | Pathogenesis of Lassa Fever | Viruses-Basel | Exclusion reason: 6 not original estimates/ primary data; |
| Yalleyogunro 1984 | Endemic Lassa Fever in Liberia .6. Village Serological Surveys for Evidence of Lassa Virus Activity in Lofa County, Liberia | Transactions of the Royal Society of Tropical Medicine and Hygiene | Exclusion reason: 1 duplicate; |
| Wulff 1975 | Recent Isolations of Lassa Virus from Nigerian Rodents | Bulletin of the World Health Organization | Exclusion reason: 5 no report of parameters ; |
| Wright 1978 | Rare virus infections from the tropics | Nursing times | Exclusion reason: 6 not original estimates/ primary data; Natsuko Imai (2019-10-24 01:13:05)(Select): paper copy; |
| Winn 1975 | Lassa Virus Hepatitis - Observations on a Fatal Case from 1972 Sierra Leone Epidemic | Archives of Pathology | Exclusion reason: 4 case report/study; Natsuko Imai (2019-10-24 01:06:13)(Select): paper copy; |

Table G.16: Excluded studies at full text review with exclusion reason

| Study | Title | Journal | Notes |
|---|---|---|---|
| Whitmer 2018 | New Lineage of Lassa Virus, Togo, 2016 | Emerging Infectious Diseases | Exclusion reason: 5 no report of parameters ; Natsuko Imai (2019-03-29 01:15:37)(Screen): from abstract, unsure whether it will report mutation rates. Including to be on the safe side.; |
| Paix 1988 | A Sero-Epidemiological Study of Hemorrhagic-Fever Viruses in a Urban-Population of Cameroon | Bulletin De La Societe De Pathologie Exotique | Exclusion reason: 2 not English; David Jorgensen (2019-06-21 23:35:35)(Select): French; |
| Omilabu 2005 | Lassa fever, Nigeria, 2003 and 2004 | Emerging Infectious Diseases | Exclusion reason: 5 no report of parameters ; |
| Olugasa 2015 | Development of a time-trend model for analyzing and predicting case-pattern of Lassa fever epidemics in Liberia, 2013-2017 | Annals of African Medicine | Exclusion reason: 5 no report of parameters ; |
| Olugasa 2015 | Mapping of Lassa fever cases in post-conflict Liberia, 2008-2012: A descriptive and categorical analysis of age, gender and seasonal pattern | Annals of African Medicine | Exclusion reason: 5 no report of parameters ; |
| Okokhere 2018 | Clinical and laboratory predictors of Lassa fever outcome in a dedicated treatment facility in Nigeria: a retrospective, observational cohort study | The Lancet. Infectious diseases | Exclusion reason: 1 duplicate; |
| O'Hearn 2016 | Serosurveillance of viral pathogens circulating in West Africa | Virology Journal | Exclusion reason: 5 no report of parameters ; |
| Ogunniyi 2014 | Lassa fever outbreak investigation in a Nigerian bakery - August, 2012 | International Journal of Infectious Diseases | Exclusion reason: 7 not peer reviewed; David Jorgensen (2019-06-21 23:27:51)(Select): Poster ; |
| Ogbu 2007 | Lassa fever in West African sub-region: an overview | Journal of Vector Borne Diseases | Exclusion reason: 6 not original estimates/ primary data; |
| Niedrig 2001 | Management and control of viral haemorrhagic fevers in Europe | Infection | Exclusion reason: 7 not peer reviewed; Natsuko Imai (2019-03-29 04:22:44)(Screen): http://www.enivd.de/NETZ.PDF; |
| Nakounne 2001 | Microbiological surveillance: viral haemorrhagic fevers in the Central African Republic; updated serological data for human beings | Bulletin De La Societe De Pathologie Exotique | Exclusion reason: 2 not English; David Jorgensen (2019-06-21 23:18:01)(Select): French ; |
| Mylne 2015 | Mapping the zoonotic niche of Lassa fever in Africa | Transactions of the Royal Society of Tropical Medicine and Hygiene | Exclusion reason: 5 no report of parameters ; Natsuko Imai (2019-10-02 21:10:53)(Select): not entirely sure what to do with spatial modelling papers? including for now,; |
| Markin 2002 | Viral haemorrhagic fevers: Evolution of the epidemic potential | Zhurnal Mikrobiologii Epidemiologii I Immunobiologii | Exclusion reason: 2 not English; David Jorgensen (2019-06-21 21:52:27)(Select): Russian ; |

Table G.16: Excluded studies at full text review with exclusion reason

| Study | Title | Journal | Notes |
|---|---|---|---|
| Manning 2015 | Lassa virus isolates from Mali and the Ivory Coast represent an emerging fifth lineage | Frontiers in Microbiology | Exclusion reason: 5 no report of parameters ; |
| Makinde 2016 | As Ebola winds down, Lassa Fever reemerges yet again in West Africa | Journal of Infection in Developing Countries | Exclusion reason: 7 not peer reviewed; Natsuko Imai (2019-03-28 01:49:00)(Screen): commentary: https://jidc.org/index.php/journal/article/view/26927465/1465; |
| Mahdy 1989 | Lassa fever: the first confirmed case imported into Canada | Canada diseases weekly report = Rapport hebdomadaire des maladies au Canada | Exclusion reason: 4 case report/study; Natsuko Imai (2019-10-24 01:21:38)(Select): paper copy; David Jorgensen (2019-06-21 21:50:42)(Select): weekly report; |
| Lukashevich 1993 | Serologic Evidence of Lassa Virus Circulation in Guinea | Voprosy virusologii | Exclusion reason: 2 not English; David Jorgensen (2019-03-29 02:54:42)(Screen): Russian; |
| Li 2016 | The etiology of Ebola virus disease-like illnesses in Ebola virus-negative patients from Sierra Leone | Oncotarget | Exclusion reason: 5 no report of parameters ; |
| Leduc 1989 | Epidemiology of Hemorrhagic-Fever Viruses | Reviews of Infectious Diseases | Exclusion reason: 3 wrong pathogen / epi not main focus; |
| Lecompte 2006 | Mastomys natalensis and Lassa fever, West Africa | Emerging Infectious Diseases | Exclusion reason: 5 no report of parameters ; Natsuko Imai (2019-10-02 20:52:30)(Select): reports human seroprevalence; |
| Lalis 2012 | The Impact of Human Conflict on the Genetics of Mastomys natalensis and Lassa Virus in West Africa | Plos One | Exclusion reason: 5 no report of parameters ; Natsuko Imai (2019-10-02 20:51:57)(Select): Reports evolutionary rate (of LASV I think, not the rodent host); |
| Lacy 1996 | Viral hemorrhagic fevers | Advances in pediatric infectious diseases | Exclusion reason: 6 not original estimates/ primary data; |
| Kyei 2015 | Imported Lassa fever: a report of 2 cases in Ghana | Bmc Infectious Diseases | Exclusion reason: 4 case report/study; |
| Saluzzo 1982 | Sero-Epidemiology of Arbovirus in the Southern Part of the Republic-of-Central-Africa | Annales De Microbiologie | Exclusion reason: 2 not English; Natsuko Imai (2019-10-26 03:18:08)(Select): abstract; David Jorgensen (2019-03-30 02:35:08)(Screen): arbovirus; |
| Saluzzo 1999 | Outbreak of human diseases associated with hantaviruses and arenaviruses hosted by rodents | M S-Medecine Sciences | Exclusion reason: 2 not English; Christian Morgenstern (2023-09-06 00:26:28)(Select): French only, hence exclude; Christian Morgenstern (2023-09-02 18:25:19)(Select): requested from library again; Natsuko Imai (2019-10-26 03:12:58)(Select): waiting on full text; |
| Saluzzo 1988 | Lassa Fever Virus in Senegal | Journal of Infectious Diseases | Exclusion reason: 5 no report of parameters ; Natsuko Imai (2019-03-28 03:15:31)(Screen): correspondence I think but include for now: |
| Kofman 2019 | Lassa Fever in Travelers from West Africa, 1969-2016 | Emerging Infectious Diseases | Exclusion reason: 6 not original estimates/ primary data; |
| Knobloch 1999 | Importing dangerous pathogens | Internist | Exclusion reason: 2 not English; |
| Kitching 2009 | A fatal case of Lassa fever in London, January 2009 | Euro surveillance : bulletin Europeen sur les maladies transmissibles = European communicable disease bulletin | Exclusion reason: 4 case report/study; |
| Kiley 1986 | Serological and Biological Evidence That Lassa-Complex Arenaviruses Are Widely Distributed in Africa | Medical Microbiology and Immunology | Exclusion reason: 5 no report of parameters ; |

Table G.16: Excluded studies at full text review with exclusion reason

| Study | Title | Journal | Notes |
|---|---|---|---|
| Khan 2008 | New opportunities for field research on the pathogenesis and treatment of Lassa fever | Antiviral Research | Exclusion reason: 6 not original estimates/ primary data; |
| Kemp 1975 | Viruses Other Than Arenaviruses from West-African Wild Mammals - Factors Affecting Transmission to Man and Domestic-Animals | Bulletin of the World Health Organization | Exclusion reason: 3 wrong pathogen / epi not main focus; David Jorgensen (2019-03-30 01:20:00)(Screen): Other than AVs?; Natsuko Imai (2019-03-28 01:36:38)(Screen): At least thirty-seven different viruses have been isolated from wild mammals in West Africa since 1962. Some of these, including Lassa virus, are already known to cause serious human morbidity and mortality. Crimean haemorrhagic fever-Congo virus, Dugbe virus, Mokola virus, and a smallpox-like agent from a gerbil in Dahomey are briefly discussed. An account of social and ecologic factors affecting man, domestic animals, and their interaction with wild mammals is given.; |
| Kaslow 2014 | Viral infections of humans: epidemiology and control | | Exclusion reason: 7 not peer reviewed; David Jorgensen (2019-06-21 21:32:14)(Select): Book chapter ; |
| Kallio-Kokko 2005 | Viral zoonoses in Europe | Fems Microbiology Reviews | Exclusion reason: 3 wrong pathogen / epi not main focus; |
| Johnson 1990 | Imported Lassa Fever - Reexamining the Algorithms | New England Journal of Medicine | Exclusion reason: 6 not original estimates/ primary data; Natsuko Imai (2019-03-27 22:54:07)(Screen): https://www.nejm.org/doi/full/10.1056/NEJM199010183231611; |
| Johnson 1993 | Emerging viruses in context: An overview of viral hemorrhagic fevers | | Exclusion reason: 7 not peer reviewed; Natsuko Imai (2019-10-26 02:52:53)(Select): exclude, book; |
| Johnson 1990 | Lymphocytic Choriomeningitis Virus Lassa Virus Lassa Fever and Other Arenaviruses | | Exclusion reason: 7 not peer reviewed; Natsuko Imai (2019-10-26 02:52:24)(Select): book exclude; |
| Johnson 1980 | Contagious Viral Hemorrhagic Fevers - Epidemiology of the Diseases and Cultures Involved and Potentially Involved with Them | American Journal of Epidemiology | Exclusion reason: 7 not peer reviewed; David Jorgensen (2019-06-21 21:27:45)(Select): conference abstract ; |
| Johnson 1979 | Andromedas Kittens - Hemorrhagic Fevers Caused by Lassa, Marburg and Ebola Viruses | Australian and New Zealand Journal of Medicine | Exclusion reason: 3 wrong pathogen / epi not main focus; Natsuko Imai (2019-10-24 01:10:53)(Select): paper copy; |
| Johnson 1975 | Arenaviruses - Some Priorities for Future Research | Bulletin of the World Health Organization | Exclusion reason: 3 wrong pathogen / epi not main focus; |
| Jay 2005 | The arenaviruses | Javma-Journal of the American Veterinary Medical Association | Exclusion reason: 6 not original estimates/ primary data; Natsuko Imai (2019-03-27 22:43:18)(Screen): https://avmajournals.avma.org/doi/pdf/10.2460/javma.2005.227.904; |
| Jahrling 1986 | Serology and Virulence Diversity among Old-World Arenaviruses, and the Relevance to Vaccine Development | Medical Microbiology and Immunology | Exclusion reason: 5 no report of parameters ; |
| Jahrling 1985 | Arenaviruses | | Exclusion reason: 7 not peer reviewed; Natsuko Imai (2019-10-26 02:44:09)(Select): exclude as book; |

Table G.16: Excluded studies at full text review with exclusion reason

| Study | Title | Journal | Notes |
|---|---|---|---|
| Jahrling 1985 | Early Diagnosis of Human Lassa Fever by Elisa Detection of Antigen and Antibody | Lancet | Exclusion reason: 4 case report/study; David Jorgensen (2019-10-24 20:28:43)(Select): Progression of Ab titre? ; Natsuko Imai (2019-10-23 23:31:43)(Select): paper copy; Natsuko Imai (2019-03-28 03:45:00)(Screen): Sequential serum samples, beginning on the day of hospital admission, from three patients with Lassa fever were tested for the presence of Lassa-virus antigens and antibodies by means of an enzyme-linked immunosorbent assay. Lassa-virus antigens were detected in the first serum sample from each patient, thus providing an early definitive diagnosis. In contrast, seroconversion was not detectable by ELISA or indirect fluorescent antibody techniques until 3 days or more after admission. The antigen-detection ELISA has important advantages over conventional infectivity titrations; it takes only hours to carry out and can be accomplished safely, with beta-propiolactone-inactivated samples. Development of antibodies coincided with a decline in antigenaemia. All acute-phase Lassa-fever sera contained either antigen or IgM antibody, and most contained both, thus allowing early diagnosis with single serum samples. More extensive testing of these ELISA techniques is recommended in field hospitals where Lassa fever is endemic and rapid diagnostic tools are needed.; |
| Jahrling 1992 | Arenaviruses and Filoviruses | | Exclusion reason: 7 not peer reviewed; |
| Jahrling 1991 | Filoviruses and Arenaviruses | | Exclusion reason: 7 not peer reviewed; Natsuko Imai (2019-10-26 02:43:26)(Select): exclude as it is a book; |
| Izzah 2016 | Geospatial Analysis of Urban Land Use Pattern Analysis for Hemorrhagic Fever Risk - a Review | | Exclusion reason: 5 no report of parameters ; |
| Hallam 2018 | Baseline mapping of Lassa fever virology, epidemiology and vaccine research and development | NPJ vaccines | Exclusion reason: 6 not original estimates/ primary data; |
| Haas 2003 | Imported Lassa fever in Germany: Surveillance and management of contact persons | Clinical Infectious Diseases | Exclusion reason: 4 case report/study; |
| Gunther 2004 | Lassa virus | Critical Reviews in Clinical Laboratory Sciences | Exclusion reason: 6 not original estimates/ primary data; |
| Grundy 1980 | Isolated Case of Lassa Fever in Zaria, Northern Nigeria | Lancet | Exclusion reason: 7 not peer reviewed; Natsuko Imai (2019-10-23 23:30:29)(Select): paper copy; David Jorgensen (2019-06-21 20:51:03)(Select): letters; |
| Gratz 1997 | The burden of rodent-borne diseases in Africa south of the Sahara | Belgian Journal of Zoology | Exclusion reason: 6 not original estimates/ primary data; |
| Gonzalez 1983 | African Viral Hemorrhagic Fevers Studies in the Central-African-Republic | Cahiers O.R.S.T.O.M. (Office de la Recherche Scientifique et Technique Outre-Mer) Serie Entomologie Medicale et Parasitologie | Exclusion reason: 2 not English; |

Table G.16: Excluded studies at full text review with exclusion reason

| Study | Title | Journal | Notes |
|---|---|---|---|
| Gonzalez 1989 | Antibody Prevalence against Hemorrhagic-Fever Viruses in Randomized Representative Central African Populations | Research in Virology | Exclusion reason: 1 duplicate; |
| Gonzalez 1986 | Evolutionary Biology of a Lassa Virus Complex | Medical Microbiology and Immunology | Exclusion reason: 5 no report of parameters ; |
| Gonzales 1985 | Genetic Variation among Lassa and Lassa Related Arenaviruses from Different African Origins | Virus Research | Exclusion reason: 3 wrong pathogen / epi not main focus; |
| Goeijenbier 2013 | Rodent-borne hemorrhagic fevers: under-recognized, widely spread and preventable - epidemiology, diagnostics and treatment | Critical Reviews in Microbiology | Exclusion reason: 6 not original estimates/ primary data; |
| Gibb 2017 | Understanding the cryptic nature of Lassa fever in West Africa | Pathogens and Global Health | Exclusion reason: 6 not original estimates/ primary data; |
| Getso 2014 | Lassa fever outbreak involving healthcare workers in Taraba State, Nigeria, March 2012 | International Journal of Infectious Diseases | Exclusion reason: 7 not peer reviewed; David Jorgensen (2019-06-21 20:36:04)(Select): poster ; |
| Gear 1977 | Hemorrhagic Fevers of Africa - Account of 2 Recent Outbreaks | Journal of the South African Veterinary Association-Tydskrif Van Die Suid-Afrikaanse Veterinere Vereniging | Exclusion reason: 4 case report/study; |
| Galbraith 1980 | Changing Patterns of Communicable Disease in England and Wales .1. Newly Recognized Diseases | Bmj-British Medical Journal | Exclusion reason: 6 not original estimates/ primary data; Natsuko Imai (2019-03-27 22:16:33)(Screen): https://www.ncbi.nlm.nih.gov/pmc/articles/PMC1713331/?page=1; |
| Galbraith 1978 | Lassa Fever (Lf) and Marburg Disease (Mvd) - Public-Health Aspects of Viral Hemorrhagic Fevers in Britain | Royal Society of Health Journal | Exclusion reason: 6 not original estimates/ primary data; |
| Ftika 2013 | Viral haemorrhagic fevers in healthcare settings | Journal of Hospital Infection | Exclusion reason: 6 not original estimates/ primary data; |
| Friedrich 2018 | Nigerian Lassa Fever Outbreak Caused by Rodents | Jama-Journal of the American Medical Association | Exclusion reason: 5 no report of parameters ; Natsuko Imai (2019-03-27 22:11:09)(Screen): commentary; |
| Frame 1992 | The Story of Lassa Fever .2. Learning More About the Disease | New York State Journal of Medicine | Exclusion reason: 6 not original estimates/ primary data; Christian Morgenstern (2023-09-06 03:41:06)(Select): page 382 out of 806 in online archive; Natsuko Imai (2019-10-26 02:30:16)(Select): waiting on full text; |

Table G.16: Excluded studies at full text review with exclusion reason

| Study | Title | Journal | Notes |
|---|---|---|---|
| Frame 1975 | Surveillance of Lassa Fever in Missionaries Stationed in West-Africa | Bulletin of the World Health Organization | Exclusion reason: 5 no report of parameters ; Natsuko Imai (2019-03-27 23:23:35)(Screen): To determine the distribution of Lassa virus in West Africa, a serological survey was undertaken. A number of mission hospital supplied sera from patients admitted with a history of fever and specimens were also collected in New York from missionaries who had experienced an unusual febrile illness while working in Africa. More cases of Lassa fever were detected among missionaries than among Africans, possibly because many African patients had left hospital before the complement fixation tests had become positive. Although most adults had fairly high fever and some were prostrated, fever was less severe in the children examined. In general the findings confirm that not all Lassa fever patients have the severe syndrome described in the original reports; |
| Morens 1980 | Longitudinal Surveillance of Lassa Fever in a West-African Village with Endemic Disease - a One-Year Report | American Journal of Epidemiology | Exclusion reason: 7 not peer reviewed; Natsuko Imai (2019-10-26 03:06:40)(Select): abstract exclude; |
| Moraz 2011 | Pathogenesis of arenavirus hemorrhagic fevers | Expert Review of Anti-Infective Therapy | Exclusion reason: 6 not original estimates/ primary data; |
| Monath 1974 | Lassa Virus Isolation from Mastomys Natalensis Rodents during an Epidemic in Sierra-Leone | Science | Exclusion reason: 5 no report of parameters ; Natsuko Imai (2019-03-28 01:59:13)(Screen): Lassa fever is a severe febrile illness of man, first recognized in West Africa in 1969. During an epidemic in Sierra Leone, Lassa virus was isolated for the first time from wild rodents of Mastomys natalensis. A high prevalence of infected Mastomys was found in houses occupied by patients with Lassa fever. The data presented provide the first demonstration of an extra-human cycle of Lassa virus transmission and suggest that rodent control may be an effective method of limiting the disease.; |
| Monath 1973 | Lassa Fever Current Epidemiological and Clinico Pathological Concepts | | Exclusion reason: 7 not peer reviewed; Natsuko Imai (2019-10-26 03:06:19)(Select): exclude book; |
| Monath 1987 | Lassa Fever - New Issues Raised by Field Studies in West-Africa | Journal of Infectious Diseases | Exclusion reason: 6 not original estimates/ primary data; |
| Monath 1977 | Lassa Fever Current Problems and Future Prospects | Vestnik Akademii Meditsinskikh Nauk SSSR | Exclusion reason: 2 not English; Natsuko Imai (2019-10-26 03:00:20)(Select): russian; |
| Monath 1977 | Lassa Fever - Past, Present and Possible Future Status | Medicina-Buenos Aires | Exclusion reason: 6 not original estimates/ primary data; |
| Monath 1975 | Lassa Fever - Review of Epidemiology and Epizootiology | Bulletin of the World Health Organization | Exclusion reason: 6 not original estimates/ primary data; |
| Monath 1974 | Lassa Fever and Marburg Virus Disease | Who Chronicle | Exclusion reason: 7 not peer reviewed; |
| Monath 1973 | Lassa fever: a new appraisal | Nigerian medical journal : journal of the Nigeria Medical Association | Exclusion reason: 7 not peer reviewed; Natsuko Imai (2019-11-01 03:08:12)(Select): can exclude – as not peer reviewed ; Natsuko Imai (2019-10-26 04:00:55)(Select): waiting on FT; |
| Monath 1973 | Lassa fever | Tropical Doctor | Exclusion reason: 6 not original estimates/ primary data; Natsuko Imai (2019-10-24 01:17:26)(Select): paper copy; |

Table G.16: Excluded studies at full text review with exclusion reason

| Study | Title | Journal | Notes |
|---|---|---|---|
| Mofolorunsho 2016 | Outbreak of lassa fever in Nigeria: measures for prevention and control | Pan African Medical Journal | Exclusion reason: 7 not peer reviewed; |
| Mills 2010 | Small Mammal-Associated Zoonoses | Vector-Borne and Zoonotic Diseases | Exclusion reason: 5 no report of parameters ; Natsuko Imai (2019-03-28 01:58:46)(Screen): Recognition of viruses as a major contributor to vector-borne diseases and other zoonoses dates back many decades. The term arbovirus (arthropod-borne virus) is widely recognized even among the lay public. A few rodent-borne viruses recognized long ago (e.g., the arenaviruses associated with Lassa fever and South American hemorrhagic fevers, and the hantaviruses associated with hemorrhagic fever with renal syndrome in Asia and Europe) were discussed under the rubric of arboviruses and even listed in the Arbovirus Catalog. A turning point came 17 years ago with the discovery of the often-fatal hantavirus pulmonary syndrome in the Americas. This discovery stimulated interest, funding, and research on rodent hosts of viruses and led to the discovery of dozens of new hantaviruses and arenaviruses; ultimately, the study of roboviruses (rodent-borne viruses) became a discipline in itself. In the last few years, we have learned that a separate order of mammals, the insectivores, are also hosts to likely dozens of hantaviruses whose role in disease is uncertain. The roboviruses have become the rainboviruses (rodent- and insectivore-borne viruses). The discovery of this tremendous diversity of viruses and hosts and the desire to understand their relationships to human disease and to environmental change have spawned new theories, controversies, and terminology: coevolution, cospeciation, spillover, host-jumping, bottom-up trophic cascades, dilution effects, and delayed density-dependence. The development of these concepts and much of the rapid growth in understanding of hostÄìvirusÄìhuman disease relationships are due to a multidisciplinary approach that combines ecology, epidemiology, virology, and molecular biology.; |
| Meunier 1987 | Current Serological Data on Viral Hemorrhagic Fevers in the Central-African-Republic | Bulletin De La Societe De Pathologie Exotique | Exclusion reason: 2 not English; David Jorgensen (2019-06-21 22:58:41)(Select): French; |
| Meunier 1987 | Serological Study of Hemorrhagic Fevers in the Province of Haut-Ogooue, Gabon | Annales De L Institut Pasteur-Virology | Exclusion reason: 2 not English; David Jorgensen (2019-06-21 22:57:41)(Select): French; |
| Mertens 1973 | Clinical Presentation of Lassa Fever Cases during Hospital Epidemic at Zorzor, Liberia, March-April 1972 | American Journal of Tropical Medicine and Hygiene | Exclusion reason: 4 case report/study; Natsuko Imai (2019-10-24 00:44:52)(Select): case study fewer 10 cases; Natsuko Imai (2019-10-24 00:43:38)(Select): paper copy; |
| McCormick 1986 | Lassa Virus Hepatitis - a Study of Fatal Lassa Fever in Humans | American Journal of Tropical Medicine and Hygiene | Exclusion reason: 5 no report of parameters ; Natsuko Imai (2019-10-24 01:16:15)(Select): paper copy; |
| McCormick 2002 | Lassa fever | | Exclusion reason: 7 not peer reviewed; David Jorgensen (2019-06-21 22:00:53)(Select): book chapter ; |

Table G.16: Excluded studies at full text review with exclusion reason

| Study | Title | Journal | Notes |
|---|---|---|---|
| McCormick 1999 | Lassa fever | | Exclusion reason: 7 not peer reviewed; Natsuko Imai (2019-10-26 02:53:46)(Select): book exclude; |
| McCormick 1988 | Lassa fever: epidemiology, therapy and vaccine development | Kansenshogaku zasshi. The Journal of the Japanese Association for Infectious Diseases | Exclusion reason: 6 not original estimates/ primary data; Natsuko Imai (2019-10-24 01:24:01)(Select): paper copy ; |
| McCormick 1987 | Epidemiology and control of Lassa fever | Current Topics in Microbiology and Immunology | Exclusion reason: 7 not peer reviewed; Natsuko Imai (2019-10-02 22:50:20)(Select): book - excl; |
| McCormick 1986 | Clinical, Epidemiologic, and Therapeutic Aspects of Lassa Fever | Medical Microbiology and Immunology | Exclusion reason: 7 not peer reviewed; David Jorgensen (2019-06-21 22:00:08)(Select): conference paper; |
| Mathiot 1990 | Antibodies to Hemorrhagic-Fever Viruses and to Selected Arboviruses in Monkeys from the Central-African-Republic | Transactions of the Royal Society of Tropical Medicine and Hygiene | Exclusion reason: 5 no report of parameters ; Natsuko Imai (2019-10-24 00:53:48)(Select): paper copy; Natsuko Imai (2019-03-28 21:13:59)(Screen): https://academic.oup.com/trstmh/article-abstract/84/5/732/1915728?redirectedFrom=fulltext; |
| Fichet-Calvet 2009 | Risk Maps of Lassa Fever in West Africa | PLoS Neglected Tropical Diseases | Exclusion reason: 3 wrong pathogen / epi not main focus; Kelly McCain (2023-09-14 21:05:25)(Included): environmental paper - no new oubreaks or parameters, non-transmission models; |
| Fichet-Calvet 2007 | Fluctuation of abundance and Lassa virus prevalence in Mastomys natalensis in Guinea, West Africa | Vector-Borne and Zoonotic Diseases | Exclusion reason: 5 no report of parameters ; Natsuko Imai (2019-03-27 21:54:27)(Screen): animal focus but talks about risk factors.; |
| Fichet-Calvet 2005 | Spatial distribution of commensal rodents in regions with high and low Lassa fever prevalence in Guinea | Belgian Journal of Zoology | Exclusion reason: 6 not original estimates/ primary data; Natsuko Imai (2019-10-26 02:28:57)(Select): seroprevalence in humans reported from previous study; |
| Fernandes 1984 | Lassa fever in London: environmental health aspects | Community medicine | Exclusion reason: 5 no report of parameters ; |
| Fabiyi 1979 | Lassa Fever Antibodies in Hospital Personnel in the Plateau State of Nigeria | Nigerian Medical Journal | Exclusion reason: 7 not peer reviewed; Natsuko Imai (2019-10-24 01:00:13)(Excluded): paper copy; David Jorgensen (2019-06-21 20:01:40)(Select): abstract only ; |
| Fabiyi 1975 | Use of Complement-Fixation (Cf) Test in Lassa Fever Surveillance - Evidence for Persistent Cf Antibodies | Bulletin of the World Health Organization | Exclusion reason: 1 duplicate; Natsuko Imai (2019-09-17 01:52:58)(Select): duplicate; |
| Fabiyi 1979 | Epidemiological significance of Lassa antibodies in Mastomys natalensis from several parts of Nigeria | Nigerian Journal of Medical Sciences | Exclusion reason: 5 no report of parameters ; Natsuko Imai (2019-10-24 01:26:10)(Select): seroprevalence in rodents; Natsuko Imai (2019-10-24 01:25:39)(Select): paper copy; |
| Fabiyi 1976 | Lassa fever (arenaviruses) as a public health problem | Bulletin of the Pan American Health Organization | Exclusion reason: 6 not original estimates/ primary data; David Jorgensen (2019-03-29 02:38:54)(Screen): incubation and shedding but w/o primary reference ; David Jorgensen (2019-03-29 02:37:41)(Screen): |
| Ethelwald 1978 | Lassa Fever (Lf) and Marburg Disease (Mvd) - Occurrences, Origins and Diagnoses | Royal Society of Health Journal | Exclusion reason: 7 not peer reviewed; Kelly Charniga (2019-10-31 20:40:24)(Select): perspective, exclude; David Jorgensen (2019-10-24 20:57:07)(Select): can't access full text; |
| Emond 1986 | Viral Hemorrhagic Fevers | Journal of Infection | Exclusion reason: 6 not original estimates/ primary data; |

Table G.16: Excluded studies at full text review with exclusion reason

| Study | Title | Journal | Notes |
|---|---|---|---|
| Emond 1980 | Lassa Fever | Royal Society of Health Journal | Exclusion reason: 6 not original estimates/ primary data; Natsuko Imai (2019-10-24 00:52:11)(Select): paper copy; Natsuko Imai (2019-03-27 23:21:17)(Screen): https://journals.sagepub.com/doi/abs/10.1177/146642408010000203; |
| El-Yuguda 2009 | Peridomestic rodents and Lassa fever virus infection of humans in urban and rural communities in Borno state, Nigeria | Nigerian Veterinary Journal | Exclusion reason: 5 no report of parameters ; |
| El-Bahnasawy 2015 | Lassa fever or lassa hemorrhagic fever risk to humans from rodent-borne zoonoses | Journal of the Egyptian Society of Parasitology | Exclusion reason: 8 No full text; Natsuko Imai (2019-10-24 19:46:10)(Select): full text not available from library exclude; |
| Ehlkes 2017 | Management of a Lassa fever outbreak, Rhineland-Palatinate, Germany, 2016 | Eurosurveillance | Exclusion reason: 3 wrong pathogen / epi not main focus; |
| Ehichioya 2008 | Clinical and Serological Evidence of Lassa Fever in Edo State, Nigeria | International Journal of Infectious Diseases | Exclusion reason: 7 not peer reviewed; David Jorgensen (2019-06-21 19:36:29)(Select): poster ; |
| Ehichioya 2010 | Lassa Fever, Nigeria, 2005-2008 | Emerging Infectious Diseases | Exclusion reason: 4 case report/study; David Jorgensen (2019-03-29 02:07:20)(Screen): Letters; |
| Dowdle 1980 | Exotic Viral Diseases | Yale Journal of Biology and Medicine | Exclusion reason: 6 not original estimates/ primary data; Natsuko Imai (2019-03-27 21:44:50)(Screen): Marburg virus disease, Lassa fever, monkeypox, and Ebola virus diseases of humans have all been recognized since 1967. These are examples of some of the exotic virus diseases which through importation may present a potential public health problem in the United States. Some of these viruses are also highly hazardous to laboratory and medical personnel. This paper is a review of the general characteristics, the epidemiology, and laboratory diagnosis of the exotic viruses which have been described during the last 25 years.; |
| Davis 2005 | Fluctuating rodent populations and risk to humans from rodent-borne zoonoses | Vector-Borne and Zoonotic Diseases | Exclusion reason: 3 wrong pathogen / epi not main focus; Natsuko Imai (2019-10-02 19:51:37)(Select): including for the foi model.; |
| Cummins 1990 | Lassa Fever | British Journal of Hospital Medicine | Exclusion reason: 5 no report of parameters ; Natsuko Imai (2019-10-23 23:29:49)(Select): reviewish; Natsuko Imai (2019-10-23 23:29:34)(Select): paper copy; Natsuko Imai (2019-03-28 01:15:43)(Screen): Lassa fever is an acute viral illness which causes considerable morbidity and mortality in West Africa. The risk of importing the disease into the UK is small but real, and it continues to be a worldwide concern among public health officials. This article summarizes its epidemiology and clinical presentation, and discusses current theories of its pathogenesis.; |
| Coyle 2016 | Lassa fever | Nursing | Exclusion reason: 6 not original estimates/ primary data; Natsuko Imai (2019-03-28 01:14:04)(Screen): https://insights.ovid.com/pubmed?pmid=27333233; |

Table G.16: Excluded studies at full text review with exclusion reason

| Study | Title | Journal | Notes |
|---|---|---|---|
| Coulibaly 2000 | Use of filter paper-collected blood specimens in a sero-survey for antibodies to Lassa virus in Guinea, West Africa: Validation of the technique and epidemiologic findings | American Journal of Tropical Medicine and Hygiene | Exclusion reason: 7 not peer reviewed; Natsuko Imai (2019-10-24 19:41:57)(Select): exclude - conf abstract; |
| Clayton 1979 | Lassa Fever, Marburg and Ebola Virus Diseases and Other Exotic Diseases - Is There a Risk to Canada | Canadian Medical Association Journal | Exclusion reason: 6 not original estimates/ primary data; David Jorgensen (2019-03-29 01:26:52)(Screen): has an early history with references; |
| Choi 2018 | A Case of Lassa Fever Diagnosed at a Community Hospital-Minnesota 2014 | Open Forum Infectious Diseases | Exclusion reason: 4 case report/study; |
| Casals 1974 | Lassa Fever | | Exclusion reason: 6 not original estimates/ primary data; Natsuko Imai (2019-10-24 01:25:01)(Select): paper copy; |
| Isaacson 2001 | Viral hemorrhagic fever hazards for travelers in Africa | Clinical Infectious Diseases | Exclusion reason: 6 not original estimates/ primary data; |
| Isa 2016 | Postexposure prophylaxis for Lassa fever: Experience from a recent outbreak in Nigeria | Nigerian medical journal : journal of the Nigeria Medical Association | Exclusion reason: 5 no report of parameters ; |
| Iroezindu 2015 | Lessons learnt from the management of a case of Lassa fever and follow-up of nosocomial primary contacts in Nigeria during Ebola virus disease outbreak in West Africa | Tropical Medicine & International Health | Exclusion reason: 4 case report/study; |
| Ike 2016 | Detection of clusters and geographical hotspot for Lassa fever in Edo Central Senatorial district of Nigeria: A step into a nation-wide mapping of Lassa fever | International Journal of Infectious Diseases | Exclusion reason: 7 not peer reviewed; David Jorgensen (2019-06-21 21:05:25)(Select): poster ; |
| Idemyor 2010 | Lassa Virus Infection in Nigeria: Clinical Perspective Overview | Journal of the National Medical Association | Exclusion reason: 6 not original estimates/ primary data; Natsuko Imai (2019-10-23 23:36:34)(Select): paper copy; |
| Ibekwe 2012 | Lassa fever: the challenges of curtailing a deadly disease | The Pan African medical journal | Exclusion reason: 6 not original estimates/ primary data; |
| Howard 1984 | Viral Hemorrhagic Fevers - Properties and Prospects for Treatment and Prevention | Antiviral Research | Exclusion reason: 6 not original estimates/ primary data; |
| Holmes 1990 | Lassa Fever in the United-States - Investigation of a Case and New Guidelines for Management | New England Journal of Medicine | Exclusion reason: 4 case report/study; Natsuko Imai (2019-03-27 22:34:28)(Screen): https://www.researchgate.net/publication/20939640$_{Lassa_{f}ever_{i}n_{t}he_{U}}$ |

Table G.16: Excluded studies at full text review with exclusion reason

| Study | Title | Journal | Notes |
|---|---|---|---|
| Hidalgo 2017 | Viral hemorrhagic fever in the tropics: Report from the task force on tropical diseases by the World Federation of Societies of Intensive and Critical Care Medicine | Journal of Critical Care | Exclusion reason: 6 not original estimates/ primary data; |
| Hartnett 2015 | Current and emerging strategies for the diagnosis, prevention and treatment of Lassa fever | Future Virology | Exclusion reason: 6 not original estimates/ primary data; |
| Hamblion 2016 | The challenges of mounting a successful response to a Lassa Fever outbreak in a post-EVD resource limited setting, Liberia 2016 | International Journal of Infectious Diseases | Exclusion reason: 7 not peer reviewed; David Jorgensen (2019-06-21 20:56:31)(Select): conf abs; |
| Burki 2018 | Lassa fever in Nigeria: the great unknown | Lancet | Exclusion reason: 6 not original estimates/ primary data; |
| Burke 2012 | A Review of Zoonotic Disease Surveillance Supported by the Armed Forces Health Surveillance Center | Zoonoses and Public Health | Exclusion reason: 5 no report of parameters ; |
| Bowen 2000 | Genetic diversity among lassa virus strains | Journal of Virology | Exclusion reason: 5 no report of parameters ; |
| Bonney 2013 | Hospital-Based Surveillance for Viral Hemorrhagic Fevers and Hepatitides in Ghana | PLoS Neglected Tropical Diseases | Exclusion reason: 5 no report of parameters ; |
| Boardman 2003 | Viral hemorrhagic fever | Primary Care Update for Ob/Gyns | Exclusion reason: 6 not original estimates/ primary data; |
| Bhadelia 2019 | Understanding Lassa fever | Science | Exclusion reason: 6 not original estimates/ primary data; |
| Bello 2016 | Lassa Fever in Pregnancy: Report of 2 Cases Seen at the University College Hospital, Ibadan | Case reports in obstetrics and gynecology | Exclusion reason: 4 case report/study; |
| Bannister 2010 | Viral haemorrhagic fevers imported into non-endemic countries: risk assessment and management | British Medical Bulletin | Exclusion reason: 6 not original estimates/ primary data; |
| Bangura 2009 | Epidemiology of Lassa Fever in the Mano River Union Countries of West Africa, 2004-2008 | American Journal of Tropical Medicine and Hygiene | Exclusion reason: 7 not peer reviewed; Natsuko Imai (2019-10-24 19:37:17)(Select): conf abstract; David Jorgensen (2019-06-21 19:04:03)(Select): probably a conference abstract, most of this issue is; |
| Banatvala 1986 | Lassa Fever | British Medical Journal | Exclusion reason: 1 duplicate; Natsuko Imai (2019-03-27 00:43:59)(Screen): duplicate; David Jorgensen (2019-03-15 02:34:24)(Screen): https://www.jstor.org/stable/29525137?seq=2metadata$_{i}nfo_{t}ab_{c}ontent$ |
| | Imported Lassa fever–New Jersey, 2004 | MMWR. Morbidity and mortality weekly report | Exclusion reason: 4 case report/study; |
| Attar 2015 | The history of Lassa virus | Nature Reviews Microbiology | Exclusion reason: 1 duplicate; David Jorgensen (2019-06-21 02:18:33)(Select): duplicate ; |

Table G.16: Excluded studies at full text review with exclusion reason

| Study | Title | Journal | Notes |
|---|---|---|---|
| Atkin 2009 | The first case of Lassa fever imported from Mali to the United Kingdom, February 2009 | Euro surveillance : bulletin Europeen sur les maladies transmissibles = European communicable disease bulletin | Exclusion reason: 4 case report/study; |
| Armignacco 2001 | The model of response to viral haemorrhagic fevers of the National Institute for Infectious Diseases ""Lazzaro Spallanzani"" | Journal of Biological Regulators and Homeostatic Agents | Exclusion reason: 6 not original estimates/ primary data; Natsuko Imai (2019-10-23 23:35:42)(Select): paper copy; David Jorgensen (2019-06-21 02:11:04)(Select): cant find - in spreadsheet ; |
| Amorosa 2010 | Imported Lassa Fever, Pennsylvania, USA, 2010 | Emerging Infectious Diseases | Exclusion reason: 4 case report/study; |
| Akpede 2018 | Lassa fever outbreaks in Nigeria | Expert Review of Anti-Infective Therapy | Exclusion reason: 6 not original estimates/ primary data; Natsuko Imai (2019-09-17 01:29:31)(Select): editorial; |
| Akpede 2010 | Prevalence and presentation of Lassa fever in Nigerian children | International Journal of Infectious Diseases | Exclusion reason: 7 not peer reviewed; David Jorgensen (2019-06-21 01:54:20)(Select): poster abstract ; |
| Akpede 2010 | Spatial and temporal trends of the Lassa fever epidemic in Nigeria 2001-2009, with particular reference to the Edo State experience | International Journal of Infectious Diseases | Exclusion reason: 7 not peer reviewed; David Jorgensen (2019-06-21 01:53:23)(Select): poster/conf abstract ; |
| Ajayi 2014 | Lassa fever - full recovery without ribavarin treatment: a case report | African Health Sciences | Exclusion reason: 4 case report/study; |
| Agbonlahor 2017 | Prevalence of Lassa virus among rodents trapped in three South-South States of Nigeria | Journal of Vector Borne Diseases | Exclusion reason: 5 no report of parameters ; |
| Adesina 2018 | Phylogenetic analysis and prevalence of Lassa virus in multimammate mice within the highly endemic Edo-Ondo hotspot for Lassa fever, Nigeria | Julius-Kuhn-Archiv | Exclusion reason: 7 not peer reviewed; David Jorgensen (2019-06-21 01:45:03)(Select): poster ; |
| Adedire 2014 | Outbreak of Lassa fever in a bakery: Investigation and epidemiological surveillance of contact persons - Ibadan, Oyo state Nigeria; August, 2012 | International Journal of Infectious Diseases | Exclusion reason: 7 not peer reviewed; David Jorgensen (2019-06-21 01:39:48)(Select): poster ; |
| Adedire 2015 | Investigation and Epidemiological Surveillance of Contact Persons During an Outbreak of Lassa Fever | International Journal of Epidemiology | Exclusion reason: 7 not peer reviewed; David Jorgensen (2019-06-21 01:38:46)(Select): this is a poster ; |
| Abdullahi 2015 | Sensitivity Analysis in a Lassa Fever Deterministic Mathematical Model | | Exclusion reason: 7 not peer reviewed; |
| | After the blood diamond conflict: lassa fever in Sierra Leone | Clinical infectious diseases : an official publication of the Infectious Diseases Society of America | Exclusion reason: 7 not peer reviewed; Natsuko Imai (2019-03-29 01:49:08)(Screen): commentary; |
| Dzotsi 2012 | The first cases of Lassa fever in Ghana | Ghana medical journal | Exclusion reason: 4 case report/study; |

Table G.16: Excluded studies at full text review with exclusion reason

| Study | Title | Journal | Notes |
|---|---|---|---|
| Dzingirai 2017 | Structural drivers of vulnerability to zoonotic disease in Africa | Philosophical Transactions of the Royal Society B-Biological Sciences | Exclusion reason: 6 not original estimates/ primary data; |
| Dzingirai 2017 | Zoonotic diseases: who gets sick, and why? Explorations from Africa | Critical Public Health | Exclusion reason: 5 no report of parameters ; |
| Dyer 2019 | Lassa outbreak: WHO warns of unusually rapid spread in Nigeria | Bmj-British Medical Journal | Exclusion reason: 7 not peer reviewed; David Jorgensen (2019-06-21 19:35:28)(Select): news ; |
| Durojaiye 1984 | Viral Zoonoses in Nigeria .2. Non-Rabies Zoonoses | International Journal of Zoonoses | Exclusion reason: 5 no report of parameters ; Natsuko Imai (2019-10-24 01:20:54)(Select): paper copy; |
| | Viral haemorrhagic fevers | Public health | Exclusion reason: 7 not peer reviewed; David Jorgensen (2019-06-22 00:54:27)(Select): letter from the editor; |
| | Editorial: Lassa fever | Lancet (London, England) | Exclusion reason: 7 not peer reviewed; |
| | Haemorrhagic fevers | South African medical journal = Suid-Afrikaanse tydskrif vir geneeskunde | Exclusion reason: 7 not peer reviewed; Natsuko Imai (2019-10-26 04:06:10)(Select): editorial; |
| | Unrecognized diseases in Africa. II. Lassa fever | International Journal of Epidemiology | Exclusion reason: 6 not original estimates/ primary data; Natsuko Imai (2019-03-29 01:27:43)(Screen): editorial include for references? |
| | Lassa Fever | British Medical Journal | Exclusion reason: 6 not original estimates/ primary data; |
| | Lassa fever | Lancet (London, England) | Exclusion reason: 5 no report of parameters ; Natsuko Imai (2019-10-24 01:03:15)(Select): paper copy; |
| | Epidemiology of hemorrhagic fever viruses. | Reviews of infectious diseases | Exclusion reason: 1 duplicate; |
| | Haemorrhagic fever virus activity in equatorial Africa: distribution and prevalence of filovirus reactive antibody in the Central African Republic. | Transactions of the Royal Society of Tropical Medicine and Hygiene | Exclusion reason: 1 duplicate; |
| Frame 1970 | Lassa fever, a new virus disease of man from West Africa. I. Clinical description and pathological findings. | The American journal of tropical medicine and hygiene | Exclusion reason: 1 duplicate; |
| White 1972 | Lassa fever. A study of 23 hospital cases. | Transactions of the Royal Society of Tropical Medicine and Hygiene | Exclusion reason: 1 duplicate; |
| Simpson 1978 | Viral haemorrhagic fevers of man. | Bulletin of the World Health Organization | Exclusion reason: 1 duplicate; |
| Troup 1970 | An outbreak of Lassa fever on the Jos plateau, Nigeria, in January-February 1970. A preliminary report. | The American journal of tropical medicine and hygiene | Exclusion reason: 1 duplicate; |
| | Arbovirus infections and viral haemorrhagic fevers in Uganda: a serological survey in Karamoja district, 1984. | Transactions of the Royal Society of Tropical Medicine and Hygiene | Exclusion reason: 1 duplicate; |
| Carey 1972 | Lassa fever. Epidemiological aspects of the 1970 epidemic, Jos, Nigeria. | Transactions of the Royal Society of Tropical Medicine and Hygiene | Exclusion reason: 1 duplicate; |

Table G.16: Excluded studies at full text review with exclusion reason

| Study | Title | Journal | Notes |
|---|---|---|---|
| | Antibodies to haemorrhagic fever viruses in Madagascar populations. | Transactions of the Royal Society of Tropical Medicine and Hygiene | Exclusion reason: 1 duplicate; |
| | Antibody prevalence against haemorrhagic fever viruses in randomized representative Central African populations. | Research in virology | Exclusion reason: 1 duplicate; |
| Johnson 1983 | Antibodies against haemorrhagic fever viruses in Kenya populations. | Transactions of the Royal Society of Tropical Medicine and Hygiene | Exclusion reason: 1 duplicate; |
| Galbraith 1978 | Public health aspects of viral haemorrhagic fevers in Britain. | Royal Society of Health journal | Exclusion reason: 1 duplicate; Natsuko Imai (2019-10-24 00:55:14)(Select): paper copy; |
| Winn 1975 | Lassa virus hepatitis. Observations on a fatal case from the 1972 Sierra Leone epidemic. | Archives of pathology | Exclusion reason: 1 duplicate; |
| Johnson 1975 | The arenaviruses: some priorities for future research. | Bulletin of the World Health Organization | Exclusion reason: 1 duplicate; |
| | Clinical features of Lassa fever in Liberia. | Reviews of infectious diseases | Exclusion reason: 1 duplicate; |
| Emond 1980 | Exotic infectious diseases: Lassa fever. | Royal Society of Health journal | Exclusion reason: 1 duplicate; |
| Simpson 1980 | The nasty viruses–Lassa, Marburg, and Ebola. | British journal of hospital medicine | Exclusion reason: 1 duplicate; |
| Vella 1978 | Lassa fever (LF) and Marburg disease (MVD): occurrences, origins and diagnoses. | Royal Society of Health journal | Exclusion reason: 1 duplicate; |
| Monson 1984 | Endemic Lassa fever in Liberia. I. Clinical and epidemiological aspects at Curran Lutheran Hospital, Zorzor, Liberia. | Transactions of the Royal Society of Tropical Medicine and Hygiene | Exclusion reason: 1 duplicate; |
| Frame 1984 | Endemic Lassa fever in Liberia. II. Serological and virological findings in hospital patients. | Transactions of the Royal Society of Tropical Medicine and Hygiene | Exclusion reason: 1 duplicate; |
| Keane 1977 | Lassa fever in Panguma Hospital, Sierra Leone, 1973-6. | British medical journal | Exclusion reason: 1 duplicate; |
| Fisher-Hoch 1988 | Hematologic dysfunction in Lassa fever. | Journal of medical virology | Exclusion reason: 5 no report of parameters ; |
| Frame 1992 | The story of Lassa fever. Part III: The disease in the community. | New York state journal of medicine | Exclusion reason: 1 duplicate; |
| Fraser 1974 | Lassa fever in the Eastern Province of Sierra Leone, 1970-1972. I. Epidemiologic studies. | The American journal of tropical medicine and hygiene | Exclusion reason: 1 duplicate; |

Table G.16: Excluded studies at full text review with exclusion reason

| Study | Title | Journal | Notes |
|---|---|---|---|
| Woodruff 1973 | Lassa fever in Britain: an imported case. | British medical journal | Exclusion reason: 4 case report/study; Natsuko Imai (2019-03-29 01:59:22)(Screen): The diagnosis of Lassa fever has been made in a patient at the Hospital for Tropical Diseases, London, who arrived in Britain by air from West Africa and was at the time of admission suffering from pyrexia of unknown origin. The infection is among the most dangerous of those currently known; mortality rates of 36-52% have been reported for hospitalized cases in the few outbreaks so far known. This report is therefore made with the object of bringing to the notice of practitioners the main features of the disease, particularly in so far as they influence the recognition and management. The complement fixation tests on serum from this patient and on other sera that were examined at the time enabled it to be shown that an outbreak of fever present in Sierra Leone was caused by the Lassa virus. Another patient who recently arrivived in Britain from West Africa has been admitted to the Hospital for Tropical Diseases, London, and it is probable that she has been infected ; |
| Monath 1974 | Lassa fever in the Eastern Province of Sierra Leone, 1970-1972. II. Clinical observations and virological studies on selected hospital cases. | The American journal of tropical medicine and hygiene | Exclusion reason: 1 duplicate; Natsuko Imai (2019-03-28 02:00:02)(Screen): Twelve patients hospitalized with a confirmed diagnosis of Lassa fever were studied during an epidemic of this disease in Sierra Leone. Clinical observations confirmed and extended those made in previous outbreaks in Liberia and Nigeria. Two patients were treated with plasma containing antibodies to Lassa virus; both had a favorable response. Lassa virus was isolated from serum, throat swabs, or urine from all patients sampled during the first 15 days of illness. Virus was recovered from the pharynx of patients with circulating complement-fixing (CF) antibodies. CF antibodies were detectable by the 3rd week of illness. A strain of Lassa virus from one of the patients was compared with strains recovered in Liberia and Nigeria by CF test. Serum from convalescent patients infected with Nigerian virus did not fix complement in the presence of strains from Liberia or Sierra Leone; this result may indicate antigenic differences between Lassa virus strains which must be confirmed by neutralization test.; |
| Ivanoff 1982 | Haemorrhagic fever in Gabon. I. Incidence of Lassa, Ebola and Marburg viruses in Haut-Ogoou©. | Transactions of the Royal Society of Tropical Medicine and Hygiene | Exclusion reason: 1 duplicate; |
| Bloch 1978 | A serological survey of Lassa fever in Liberia. | Bulletin of the World Health Organization | Exclusion reason: 1 duplicate; Natsuko Imai (2019-09-17 01:40:15)(Select): duplicate; |
| Frame 1984 | Endemic Lassa fever in Liberia. V. Distribution of Lassa virus activity in Liberia: hospital staff surveys. | Transactions of the Royal Society of Tropical Medicine and Hygiene | Exclusion reason: 1 duplicate; |
| Monath 1973 | A hospital epidemic of Lassa fever in Zorzor, Liberia, March-April 1972. | The American journal of tropical medicine and hygiene | Exclusion reason: 1 duplicate; |

Table G.16: Excluded studies at full text review with exclusion reason

| Study | Title | Journal | Notes |
|---|---|---|---|
| Mertens 1973 | Clinical presentation of Lassa fever cases during the hospital epidemic at Zorzor, Liberia, March-April 1972. | The American journal of tropical medicine and hygiene | Exclusion reason: 1 duplicate; |
| Sogoba 2012 | Lassa fever in West Africa: evidence for an expanded region of endemicity. | Zoonoses and public health | Exclusion reason: 6 not original estimates/ primary data; |
| Bond 2013 | A historical look at the first reported cases of Lassa fever: IgG antibodies 40 years after acute infection. | The American journal of tropical medicine and hygiene | Exclusion reason: 4 case report/study; Christian Morgenstern (2023-08-30 03:37:40)(Included): Case report of 2 survivors after 40 years but no serostudy or other relevant parameters; Christian Morgenstern (2023-08-30 03:37:13)(Included): records long lasting IgG (40+ years); |
| Woodruff 1978 | Viral infections in travellers from tropical Africa. | British medical journal | Exclusion reason: 1 duplicate; |
| Attar 2015 | Viral evolution: The history of Lassa virus. | Nature reviews. Microbiology | Exclusion reason: 6 not original estimates/ primary data; David Jorgensen (2019-06-21 02:18:13)(Select): nature research highlight of a cell paper; check original is present ; |
| Smither 2023 | Ecology of Lassa Virus. | Current topics in microbiology and immunology | Exclusion reason: 7 not peer reviewed; |
| Musa 2022 | COVID-19 and Lassa fever in Nigeria: A deadly alliance? | International journal of infectious diseases : IJID : official publication of the International Society for Infectious Diseases | Exclusion reason: 5 no report of parameters ; |
| Wang 2021 | The reproductive number of Lassa fever: a systematic review. | Journal of travel medicine | Exclusion reason: 6 not original estimates/ primary data; |
| Klitting 2022 | Predicting the evolution of the Lassa virus endemic area and population at risk over the next decades. | Nature communications | Exclusion reason: 5 no report of parameters ; Juliette Unwin (2023-09-05 17:52:21)(Select): future risk; |
| Mohammed 2023 | Inter-State Transmission of Lassa Fever during the 2015-2016 Lassa Outbreak in Nigeria: An Implication for Infection Prevention and Control Practices. | West African journal of medicine | Exclusion reason: 8 No full text; Christian Morgenstern (2023-09-09 01:47:20)(Select): Imperial College Library and British Library both unable to obtain Can't find full text online either; Christian Morgenstern (2023-09-02 18:43:02)(Select): requested from library; |
| Kenmoe 2020 | Systematic review and meta-analysis of the epidemiology of Lassa virus in humans, rodents and other mammals in sub-Saharan Africa. | PLoS neglected tropical diseases | Exclusion reason: 6 not original estimates/ primary data; |
| Wada 2022 | Knowledge of Lassa fever, its prevention and control practices and their predictors among healthcare workers during an outbreak in Northern Nigeria: A multi-centre cross-sectional assessment. | PLoS neglected tropical diseases | Exclusion reason: 5 no report of parameters ; |

Table G.16: Excluded studies at full text review with exclusion reason

| Study | Title | Journal | Notes |
|---|---|---|---|
| Happi 2022 | Increased Prevalence of Lassa Fever Virus-Positive Rodents and Diversity of Infected Species Found during Human Lassa Fever Epidemics in Nigeria. | Microbiology spectrum | Exclusion reason: 3 wrong pathogen / epi not main focus; Christian Morgenstern (2023-08-17 00:04:15)(Screen): Keep for write-up / information on prevalence in rodents. (reject at full text screening stage); |
| Babalola 2019 | Lassa virus RNA detection from suspected cases in Nigeria, 2011-2017. | The Pan African medical journal | Exclusion reason: 5 no report of parameters ; Kelly McCain (2023-09-25 23:41:19)(Included): Reports only PCR results not seroprevalence; Juliette Unwin (2023-09-02 00:24:47)(Select): wouldn't you show this from the context part of the form. It's sort of an attack rate. Happy to be overruled though; David Jorgensen (2023-08-22 19:37:54)(Screen): sero in suspected cases not in the population; |
| Baumann 2019 | Lassa and Crimean-Congo Hemorrhagic Fever Viruses, Mali. | Emerging infectious diseases | Exclusion reason: 4 case report/study; Juliette Unwin (2023-09-02 00:40:34)(Select): it's a dispatch so unclear if peer reviewed. unsure what would extract its sort of a cases study 2/xx samples positive; |
| Li 2023 | Genetic basis underlying Lassa fever endemics in the Mano River region, West Africa. | Virology | Exclusion reason: 5 no report of parameters ; |
| Douno 2021 | Hunting and consumption of rodents by children in the Lassa fever endemic area of Faranah, Guinea. | PLoS neglected tropical diseases | Exclusion reason: 3 wrong pathogen / epi not main focus; |
| Kakaf´ 2020 | Improving Cross-Border Preparedness and Response: Lessons Learned from 3 Lassa Fever Outbreaks Across Benin, Nigeria, and Togo, 2017-2019. | Health security | Exclusion reason: 3 wrong pathogen / epi not main focus; |
| Nuismer 2020 | Bayesian estimation of Lassa virus epidemiological parameters: Implications for spillover prevention using wildlife vaccination. | PLoS neglected tropical diseases | Exclusion reason: 3 wrong pathogen / epi not main focus; Juliette Unwin (2023-09-05 17:46:12)(Select): rodent model ; |
| Sesay 2022 | Late diagnosis of Lassa fever outbreak in endemic areas lead to high mortality, Kenema District, Sierra Leone, February - March 2019. | The Pan African medical journal | Exclusion reason: 4 case report/study; |
| Forni 2020 | Population structure of Lassa Mammarenavirus in West Africa. | Viruses | Exclusion reason: 5 no report of parameters ; Christian Morgenstern (2023-08-30 22:22:26)(Select): Genetic diversity clustered geographically; |
| Thielebein 2022 | Virus persistence after recovery from acute Lassa fever in Nigeria: a 2-year interim analysis of a prospective longitudinal cohort study. | The Lancet. Microbe | Exclusion reason: 5 no report of parameters ; |

Table G.16: Excluded studies at full text review with exclusion reason

| Study | Title | Journal | Notes |
|---|---|---|---|
| Merson 2021 | Clinical characterization of Lassa fever: A systematic review of clinical reports and research to inform clinical trial design. | PLoS neglected tropical diseases | Exclusion reason: 6 not original estimates/ primary data; David Jorgensen (2023-08-16 23:39:25)(Screen): systematic review of clinical reports ; |
| Penfold 2023 | A prospective, multi-site, cohort study to estimate incidence of infection and disease due to Lassa fever virus in West African countries (the Enable Lassa research programme)-Study protocol. | PloS one | Exclusion reason: 3 wrong pathogen / epi not main focus; |
| Eberhardt 2019 | Ribavirin for the treatment of Lassa fever: A systematic review and meta-analysis. | International journal of infectious diseases : IJID : official publication of the International Society for Infectious Diseases | Exclusion reason: 6 not original estimates/ primary data; David Jorgensen (2023-08-16 19:36:42)(Screen): systematic review/meta; |
| Kulkarni 2018 | Case Report: Imported Case of Lassa Fever - New Jersey, May 2015. | The American journal of tropical medicine and hygiene | Exclusion reason: 4 case report/study; |
| Mari´n 2019 | Evaluation of rodent control to fight Lassa fever based on field data and mathematical modelling. | Emerging microbes & infections | Exclusion reason: 3 wrong pathogen / epi not main focus; Christian Morgenstern (2023-08-27 01:41:38)(Select): Rodent based study; |
| Clark 2021 | Domestic risk factors for increased rodent abundance in a Lassa fever endemic region of rural Upper Guinea. | Scientific reports | Exclusion reason: 3 wrong pathogen / epi not main focus; Juliette Unwin (2023-09-05 17:33:56)(Select): risks for rodents not Lassa; |
| Kraft 2020 | Serosurvey on healthcare personnel caring for patients with Ebola virus disease and Lassa virus in the United States. | Infection control and hospital epidemiology | Exclusion reason: 4 case report/study; |
| Mari´n 2020 | Households as hotspots of Lassa fever? Assessing the spatial distribution of Lassa virus-infected rodents in rural villages of Guinea. | Emerging microbes & infections | Exclusion reason: 3 wrong pathogen / epi not main focus; Juliette Unwin (2023-09-05 22:19:43)(Select): rodent study; |
| Adesina 2023 | Circulation of Lassa virus across the endemic Edo-Ondo axis, Nigeria, with cross-species transmission between multimammate mice. | Emerging microbes & infections | Exclusion reason: 3 wrong pathogen / epi not main focus; David Jorgensen (2023-08-22 01:26:51)(Select): Phylo but only in rats, could be nice for a future phylo model though; David Jorgensen (2023-08-16 19:42:03)(Screen): think it only covers rats but check full text; |
| Min 2021 | An exploration of the protective effect of rodent species richness on the geographical expansion of Lassa fever in West Africa. | PLoS neglected tropical diseases | Exclusion reason: 3 wrong pathogen / epi not main focus; David Jorgensen (2023-08-25 23:49:55)(Select): risk factors ; |

Table G.16: Excluded studies at full text review with exclusion reason

| Study | Title | Journal | Notes |
|---|---|---|---|
| Overbosch 2020 | Public health response to two imported, epidemiologically related cases of Lassa fever in the Netherlands (ex Sierra Leone), November 2019. | Euro surveillance : bulletin Europeen sur les maladies transmissibles = European communicable disease bulletin | Exclusion reason: 4 case report/study; |
| Ipadeola 2020 | Epidemiology and case-control study of Lassa fever outbreak in Nigeria from 2018 to 2019. | The Journal of infection | Exclusion reason: 7 not peer reviewed; |
| Okoro 2020 | Descriptive epidemiology of Lassa fever in Nigeria, 2012-2017. | The Pan African medical journal | Exclusion reason: 5 no report of parameters ; |
| Agbonlahor 2021 | 52 Years of Lassa Fever Outbreaks in Nigeria, 1969-2020: An Epidemiologic Analysis of the Temporal and Spatial Trends. | The American journal of tropical medicine and hygiene | Exclusion reason: 6 not original estimates/ primary data; |
| Kajero 2019 | New methodologies for the estimation of population vulnerability to diseases: a case study of Lassa fever and Ebola in Nigeria and Sierra Leone. | Philosophical transactions of the Royal Society of London. Series B, Biological sciences | Exclusion reason: 5 no report of parameters ; David Jorgensen (2023-08-30 20:43:14)(Select): included but is a strange model without normal epi parameters reported; |
| Kayem 2020 | Lassa fever in pregnancy: a systematic review and meta-analysis. | Transactions of the Royal Society of Tropical Medicine and Hygiene | Exclusion reason: 6 not original estimates/ primary data; David Jorgensen (2023-08-16 23:43:58)(Screen): systematic review ; |
| Wolf 2020 | Fifty years of imported Lassa fever: a systematic review of primary and secondary cases. | Journal of travel medicine | Exclusion reason: 6 not original estimates/ primary data; David Jorgensen (2023-08-16 18:59:22)(Screen): systematic - check for inclusion criteria; |
| Kolawole 2021 | Phylogenetic and Mutational Analysis of Lassa Virus Strains Isolated in Nigeria: Proposal for an In Silico Study. | JMIR research protocols | Exclusion reason: 5 no report of parameters ; |
| Salu 2019 | Molecular confirmation and phylogeny of Lassa fever virus in Benin Republic 2014-2016. | African journal of laboratory medicine | Exclusion reason: 5 no report of parameters ; David Jorgensen (2023-08-16 19:29:02)(Screen): only 5 sequences reconstructed; |
| Okokhere 2016 | Aseptic Meningitis Caused by Lassa Virus: Case Series Report. | Case reports in neurological medicine | Exclusion reason: 4 case report/study; |
| Wood 2021 | Detection of Lassa virus in wild rodent feces: Implications for Lassa fever burden within households in the endemic region of Faranah, Guinea. | One health (Amsterdam, Netherlands) | Exclusion reason: 5 no report of parameters ; Juliette Unwin (2023-09-02 00:32:09)(Select): don't think any parameters we want to extract; |
| Smither 2023 | Novel Tools for Lassa Virus Surveillance in Peri-domestic Rodents. | medRxiv : the preprint server for health sciences | Exclusion reason: 3 wrong pathogen / epi not main focus; Christian Morgenstern (2023-08-23 03:52:12)(Screen): pre-print; |

Table G.16: Excluded studies at full text review with exclusion reason

| Study | Title | Journal | Notes |
|---|---|---|---|
| Olayemi 2020 | Determining Ancestry between Rodent- and Human-Derived Virus Sequences in Endemic Foci: Towards a More Integral Molecular Epidemiology of Lassa Fever within West Africa. | Biology | Exclusion reason: 5 no report of parameters ; |
| Tahmo 2023 | Modeling the Lassa fever outbreak synchronously occurring with cholera and COVID-19 outbreaks in Nigeria 2021: A threat to Global Health Security. | PLOS global public health | Exclusion reason: 3 wrong pathogen / epi not main focus; Christian Morgenstern (2023-09-01 22:26:58)(Select): time series model without disease transmission mechanism ; David Jorgensen (2023-08-16 19:16:01)(Screen): co-modelled ; |
| Simons 2022 | Lassa fever cases suffer from severe underreporting based on reported fatalities. | International health | Exclusion reason: 6 not original estimates/ primary data; Juliette Unwin (2023-09-02 00:47:30)(Select): Review uses data from other places; |
| Bangura 2021 | Lassa Virus Circulation in Small Mammal Populations in Bo District, Sierra Leone. | Biology | Exclusion reason: 3 wrong pathogen / epi not main focus; |
| Obionu 2021 | Evaluation of infection prevention and control practices in Lassa fever treatment centers in north-central Nigeria during an ongoing Lassa fever outbreak. | Journal of infection prevention | Exclusion reason: 5 no report of parameters ; David Jorgensen (2023-08-25 23:58:25)(Select): No outcome data; |
| Akpede 2019 | Corrigendum: Caseload and Case Fatality of Lassa Fever in Nigeria, 2001-2018: A Specialist Center's Experience and Its Implications. | Frontiers in public health | Exclusion reason: 1 duplicate; David Jorgensen (2023-08-26 00:50:00)(Select): Author name correction to a paper we already have; |
| Grahn 2016 | Imported Case of Lassa Fever in Sweden With Encephalopathy and Sensorineural Hearing Deficit. | Open forum infectious diseases | Exclusion reason: 4 case report/study; Christian Morgenstern (2023-08-24 03:54:00)(Select): Single case, 118 close contacts with no onwards transmission; |
| Gobir 2019 | Hygiene Practices in a Nigerian rural Community during Lassa Fever Epidemic. | Saudi journal of medicine & medical sciences | Exclusion reason: 5 no report of parameters ; David Jorgensen (2023-08-26 00:01:20)(Select): also not a full article; David Jorgensen (2023-08-16 19:03:39)(Screen): risk factor but no link to outcome ; |
| Whitlock 2023 | Identifying the genetic basis of viral spillover using Lassa virus as a test case. | Royal Society open science | Exclusion reason: 5 no report of parameters ; David Jorgensen (2023-08-26 00:56:21)(Select): identifying genetic variants rather than epi parameters to do with spillover; |
| Begeman 2023 | The pathogenesis of zoonotic viral infections: Lessons learned by studying reservoir hosts. | Frontiers in microbiology | Exclusion reason: 3 wrong pathogen / epi not main focus; |
| Simons 2023 | Rodent trapping studies as an overlooked information source for understanding endemic and novel zoonotic spillover. | PLoS neglected tropical diseases | Exclusion reason: 5 no report of parameters ; David Jorgensen (2023-08-22 19:52:18)(Screen): meta-analysis; |

Table G.16: Excluded studies at full text review with exclusion reason

| Study | Title | Journal | Notes |
|---|---|---|---|
| Gulumbe 2023 | Recurring Outbreaks of Lassa Fever in Nigeria: Understanding the Root Causes and Strategies for the Future | SUDAN JOURNAL OF MEDICAL SCIENCES | Exclusion reason: 5 no report of parameters ; |
| Alli 2021 | Management of Lassa Fever: A Current Update | CUREUS JOURNAL OF MEDICAL SCIENCE | Exclusion reason: 6 not original estimates/ primary data; David Jorgensen (2023-08-16 19:44:17)(Screen): systematic review ; |
| Adewole 2022 | Lassa fever in pregnancy: Report of two maternal deaths in a tertiary center in the middle-belt region of Nigeria | AFRICAN JOURNAL OF REPRODUCTIVE HEALTH | Exclusion reason: 4 case report/study; |
| Balogun 2021 | Lassa Fever: An Evolving Emergency in West Africa | AMERICAN JOURNAL OF TROPICAL MEDICINE AND HYGIENE | Exclusion reason: 6 not original estimates/ primary data; |
| MONATH 1975 | LASSA FEVER - REVIEW OF EPIDEMIOLOGY AND EPIZOOTIOLOGY | BULLETIN OF THE WORLD HEALTH ORGANIZATION | Exclusion reason: 6 not original estimates/ primary data; David Jorgensen (2023-08-26 01:09:57)(Select): Seems to be some unpublished data but is zero seroprevalence data; David Jorgensen (2023-08-16 18:34:59)(Screen): review - check refs; |
| Uyigue 2018 | IMMUNE RESPONSE PROFILING OF SUSPECTED LASSA FEVER CASES AT IRRUA SPECIALIST TEACHING HOSPITAL DURING 2018 LASSA FEVER OUTBREAK | AMERICAN JOURNAL OF TROPICAL MEDICINE AND HYGIENE | Exclusion reason: 7 not peer reviewed; Christian Morgenstern (2023-09-04 19:37:20)(Select): conference abstract; Christian Morgenstern (2023-09-02 18:16:27)(Select): requested from library; David Jorgensen (2023-08-25 23:50:49)(Select): can't find ; |
| Oyegoke 2020 | Lassa fever outbreak in Bauchi state, Nigeria January to April 2019 | INTERNATIONAL JOURNAL OF INFECTIOUS DISEASES | Exclusion reason: 7 not peer reviewed; Juliette Unwin (2023-09-02 00:49:55)(Select): outbreak data; |
| WULFF 1975 | INDIRECT IMMUNOFLUORESCENCE FOR DIAGNOSIS OF LASSA FEVER INFECTION | BULLETIN OF THE WORLD HEALTH ORGANIZATION | Exclusion reason: 3 wrong pathogen / epi not main focus; |
| KILEY 1986 | SEROLOGICAL AND BIOLOGICAL EVIDENCE THAT LASSA-COMPLEX ARENAVIRUSES ARE WIDELY DISTRIBUTED IN AFRICA | MEDICAL MICROBIOLOGY AND IMMUNOLOGY | Exclusion reason: 5 no report of parameters ; Christian Morgenstern (2023-08-22 21:00:26)(Select): sero study in rodent population only; |
| Mba 2020 | A description of Lassa Fever mortality during the 2019 outbreak in Nigeria | INTERNATIONAL JOURNAL OF INFECTIOUS DISEASES | Exclusion reason: 8 No full text; Christian Morgenstern (2023-08-30 21:10:58)(Select): Abstract only in a supplement; |
| Ilesanmi 2019 | MORTALITY AMONG CONFIRMED LASSA FEVER CASES DURING THE 2017-2019 OUTBREAK IN ONDO STATE, NIGERIA | TRANSACTIONS OF THE ROYAL SOCIETY OF TROPICAL MEDICINE AND HYGIENE | Exclusion reason: 7 not peer reviewed; David Jorgensen (2023-08-30 18:30:56)(Select): wrong journal for the article - probably a conference presentation?; |

Table G.16: Excluded studies at full text review with exclusion reason

| Study | Title | Journal | Notes |
|---|---|---|---|
| Siddle 2018 | GENOMIC EPIDEMIOLOGY OF LASSA VIRUS IN NIGERIA DURING A SEASON OF UNUSUALLY HIGH INCIDENCE | AMERICAN JOURNAL OF TROPICAL MEDICINE AND HYGIENE | Exclusion reason: 7 not peer reviewed; Christian Morgenstern (2023-09-04 19:38:26)(Select): conference abstract; Christian Morgenstern (2023-09-02 18:17:52)(Select): requested from library to confirm; David Jorgensen (2023-08-26 00:30:50)(Select): Probably a conference title - seemed to come out in nejm with a different title; |
| Uzoma 2020 | Increasing trend of Lassa fever outbreak in Nigeria: The more you look, the more you see | INTERNATIONAL JOURNAL OF INFECTIOUS DISEASES | Exclusion reason: 6 not original estimates/ primary data; Christian Morgenstern (2023-08-31 22:45:51)(Select): abstract in supplement; David Jorgensen (2023-08-16 18:29:57)(Screen): Review ; Natsuko Imai (2023-08-16 01:26:45)(Screen): Background: Lassa fever (LF) is an acute viral hemorrhagic fever endemic in Nigeria. The bane of endemicity of LF has reached an unprecedented proportion in recent years; with each yearly outbreak bigger in cases, deaths and spread, compared to the previous years. A critical review of the increase and associated factors are presented in this paper. Methods and materials: This study is based on desk review, analysis of secondary data, review of surveillance indicators and Key Informant Interviews (KII) with members of the National Lassa Fever Technical Working Group at the Nigeria Centre for Disease Control. Case definition for suspected and confirmed cases of Lassa fever were interpreted as contained in the National IDSR Guideline. Prevalence, Case Fatality Rate (CFR) and Timeliness of report were computed and compared across the three years. Results: The study observed that reliable Lassa fever data was unavailable at the national level before 2017. However, in 2017, 1022 suspected cases were identified in 19 states and 322 tested positive for Lassa fever. Between 2017 and 2018, states have received training and tools on Lassa fever surveillance, state surveillance and response structure had been standardized and preparedness assessment were carried out. Case detection started early in 2018 and by December, 3498 suspects were identified representing about three-fold increase over previous year, and 633 cases tested positive. With further strengthening of the surveillance system in 2019, a total of 4099 suspected cases were identified with 739 confirmed for weeks 1Äì41, 2019. Similarly, deaths among cases increased from 127 (39.4% CFR) to 171 (27% CFR) to 154 (20.8% CFR) over the same time period. Conclusion: The period between 2016 and 2019 signify the improvement of the surveillance system for infectious diseases in Nigeria. The result can be translated to mean increase in number of cases of Lassa fever detection. The percent increase from 2018 to 2019 is 17% and this would continue to reduce until we reach an equilibrium. Only until this happens can we speculate on the burden of LF or we can also conduct a seroprevalence study to determine the true burden of the infectious disease.; |
| Belhadi 2022 | The number of cases, mortality and treatments of viral hemorrhagic fevers: A systematic review | PLOS NEGLECTED TROPICAL DISEASES | Exclusion reason: 6 not original estimates/ primary data; David Jorgensen (2023-08-16 19:47:45)(Screen): systematic review but covers mutiple hemorrhagic fevers; |

Table G.16: Excluded studies at full text review with exclusion reason

| Study | Title | Journal | Notes |
|---|---|---|---|
| Agboeze 2020 | Epidemiological investigation of Lassa fever outbreak in a resource limited setting: Lessons learnt. (January to March 2018) | INTERNATIONAL JOURNAL OF INFECTIOUS DISEASES | Exclusion reason: 7 not peer reviewed; David Jorgensen (2023-08-22 01:33:07)(Select): conference abstract; Natsuko Imai (2023-08-16 01:41:45)(Screen): Background: An outbreak of Lassa Fever (LF) reported and confirmed in Ebonyi state, Southwest Nigeria in January 2018 was investigated. This article provides the epidemiology of the LF and lessons learnt from the investigation of the outbreak. Methods and materials: The Emergency operational centre (EOC) model was used for the outbreak coordination. Cases and deaths were identified through the routine surveillance system using standard definitions for suspected and confirmed cases and deaths respectively. Blood specimens collected from suspected cases were sent for confirmation at the virology centre Abakaliki. Active case search was intensified, and identified contacts of confirmed cases were followed up for the maximum incubation period of the disease. Other public health responses included infection prevention and control, communication and advocacy as well as case management. Data collected were analysed using Epi info, by time, place and persons and important lessons drawn were discussed. Results: We identified 89 suspected LF cases of which 61 were confirmed with mean age of 35ÄÖ¬±ÄÖ16.2. More than half (59.7%) of the confirmed cases were females and the age group mostly affected was 30Äì39 years. Majority of the cases presented late. The Case Fatality Rate (CFR) was 68.5% among the laboratory confirmed cases. Among the health care workers 5 confirmed cases died. Three hundred and twenty-five contacts of the confirmed cases were identified, out of which 304(99.7%) completed the follow-up without developing any symptoms and 1(0.3%) developed symptoms consistent with LF and was confirmed by the laboratory. Conclusion: Key lessons learnt from the investigation were: high fatality rate in those presenting late to the hospital and especially in those presenting with complications underscore the need for intensive public enlightenment that encourages early presentation to hospital, majority of the confirmed cases were primary cases hence efforts should be intensified in breaking the chain of transmission in the animalÄìman interphase, death of healthcare workers involved in management of Lassa fever raises the importance of providing life insurance. The presence of virology centre and the impact of both local and international partners contributed significantly in curbing the outbreak.; |
| Ulor 2020 | Environmental determinants and markers of Lassa fever transmission in a low socioeconomic community in Akure, southwestern Nigeria | INTERNATIONAL JOURNAL OF INFECTIOUS DISEASES | Exclusion reason: 5 no report of parameters ; Christian Morgenstern (2023-08-17 00:18:46)(Select): Supplement, not the actual article; David Jorgensen (2023-08-16 18:52:46)(Screen): risk factor ; |

Table G.16: Excluded studies at full text review with exclusion reason

| Study | Title | Journal | Notes |
|---|---|---|---|
| Ejembi 2020 | Knowledge and attitude of community members and health care workers on Lassa fever during an outbreak in Kogi State, Nigeria 2016 | INTERNATIONAL JOURNAL OF INFECTIOUS DISEASES | Exclusion reason: 7 not peer reviewed; David Jorgensen (2023-08-30 18:34:04)(Select): supplement - conference presentation?; |
| Chika-Igwenyi 2020 | Another form of Lassa fever? Early neurological symptoms and high mortality reveal differences in two outbreaks in Ebonyi State, Nigeria 2017-2019 | INTERNATIONAL JOURNAL OF INFECTIOUS DISEASES | Exclusion reason: 7 not peer reviewed; Juliette Unwin (2023-09-05 22:46:48)(Select): conference; Natsuko Imai (2023-08-16 01:57:01)(Screen): Background: Lassa fever (LF) is an acute viral haemorrhagic illness with various clinical manifestations. Neurological symptoms are not commonly present at the early stage of the disease; however, early manifestation of central nervous system features depicts poor prognostication. In Ebonyi state, an unusual pattern was observed between two outbreaks with patients presenting early neurological symptoms and a high mortality rate in the second outbreak. The study described the epidemiological evolution, socio-demographic profiles, clinical characteristics and patientsÄô outcomes. Methods and materials: A retrospective analytic analysis of routinely collected clinical data was conducted of all confirmed and probable LF patients admitted to the Virology Centre of the AEFUTHA in Ebonyi State, December 2017 to January 2019. Results: In a total of 83 cases, 70 were RT-PCR confirmed and 13 probable cases. In outbreak 1, 69 were seen with 53.6% being urban residents, 19% farmers, 15% students, and 10% health workers. Fourteen cases were seen in outbreak 2 with 92.9% rural residents, 58.3% being farmers and 49.9% students. There were differences in clinical and laboratory signs and symptoms between the two outbreaks with neurological symptoms present 43% of the time in outbreak 1 and 93% in outbreak 2 (pÄÖ=ÄÖ0.001), with a shorter time of onset for these symptoms in outbreak 2. The mortality rate was 85.7% in outbreak 2 versus 29.9% in outbreak 1 (pÄÖ¡ÄÖ0.001). Patients with neurological symptoms, who were more common in outbreak 2 had a RR of dying of 8.5 compared to those without. Conclusion: This study revealed a different form of LF that is of great concern due to its high mortality rate. Further studies are needed to better define its characteristics; |
| Starnes 2021 | Diagnosis and Management of Lassa Fever Among Children in Eastern Province, Sierra Leone: A 7-year Retrospective Analysis | PEDIATRICS | Exclusion reason: 7 not peer reviewed; Christian Morgenstern (2023-09-02 18:34:37)(Select): contains risk factors for children but is meeting / conference abstract; |
| Adeyemi 2019 | LASSA FEVER SEROPREVALENCE IN PLATEAU STATE, NIGERIA | AMERICAN JOURNAL OF TROPICAL MEDICINE AND HYGIENE | Exclusion reason: 7 not peer reviewed; Christian Morgenstern (2023-09-04 19:41:59)(Select): conference abstract; Christian Morgenstern (2023-09-02 18:30:55)(Select): requested from library; Juliette Unwin (2023-09-02 02:26:11)(Select): I think this one is probably from a conference as one page; David Jorgensen (2023-08-22 01:30:43)(Select): can't find ; |

Table G.16: Excluded studies at full text review with exclusion reason

| Study | Title | Journal | Notes |
|---|---|---|---|
| FRAME 1992 | THE STORY OF LASSA FEVER .3. THE DISEASE IN THE COMMUNITY | NEW YORK STATE JOURNAL OF MEDICINE | Exclusion reason: 6 not original estimates/ primary data; Patrick Doohan (2023-09-06 02:43:14)(Select): Available in paper copy in the office; David Jorgensen (2023-08-16 18:33:36)(Screen): Can't find, could be some epi?; |
| Fichet-Calvet 2019 | Estimation of Lassa virus emergence in Upper Guinea through a time-calibrated phylogeny | VIRUS EVOLUTION | Exclusion reason: 7 not peer reviewed; Christian Morgenstern (2023-08-30 22:43:03)(Select): Abstract in supplement only; |
| HOLMES 1990 | LASSA FEVER IN THE UNITED-STATES - INVESTIGATION OF A CASE AND NEW GUIDELINES FOR MANAGEMENT | NEW ENGLAND JOURNAL OF MEDICINE | Exclusion reason: 4 case report/study; Christian Morgenstern (2023-08-24 04:18:11)(Select): single case report, 102 contacts with no onward transmission; |
| MORENS 1980 | LONGITUDINAL SURVEILLANCE OF LASSA FEVER IN A WEST-AFRICAN VILLAGE WITH ENDEMIC DISEASE - A ONE-YEAR REPORT | AMERICAN JOURNAL OF EPIDEMIOLOGY | Exclusion reason: 7 not peer reviewed; Christian Morgenstern (2023-09-04 22:41:14)(Select): conference abstract; |

Table G.16: Excluded studies at full text review with exclusion reason

### H Pathogen Epidemiology Review Group (PERG) Membership

| First Name | Surname | Affiliation |
|---|---|---|
| Aaron | Morris | University of Oxford |
| Alpha | Forna | University of Georgia |
| Amy | Dighe | Johns Hopkins |
| Anne | Cori | Imperial College London |
| Arran | Hamlet | Imperial College London |
| Ben | Lambert | Manchester University |
| Charlie | Whittaker | Imperial College London |
| Christian | Morgenstern | Imperial College London |
| Cyril | Geismar | Imperial College London |
| Dariya | Nikitin | Imperial College London |
| David | Jorgensen | Imperial College London |
| Ed | Knock | Imperial College London |
| Ettie | Unwin | Imperial College London |
| Gina | Cuomo-Dannenburg | Imperial College London |
| Hayley | Thompson | PATH |
| Isobel | Routledge | UCSF |
| Janetta | Skarp | Imperial College London |
| Joseph | Hicks | Imperial College London |
| Keith | Fraser | Imperial College London |
| Kelly | Charniga | Imperial College London |
| Kelly | McCain | Imperial College London |
| Lily | Geidelberg | Imperial College London |
| Lorenzo | Cattarino | UKHSA |
| Mara | Kont | Imperial College London |
| Marc | Baguelin | Imperial College London |
| Natsuko | Imai | Imperial College London |
| Nima | Moghaddas | Imperial College London |
| Patrick | Doohan | Imperial College London |
| Rebecca | Nash | Imperial College London |
| Ruth | McCabe | University of Oxford |
| Sabine | van Elsland | Imperial College London |
| Sangeeta | Bhatia | Imperial College London |
| Sreejith | Radhakrishnan | University of Glasgow |
| Zulma | Cucunuba Perez | Pontificia Universidad Javeriana |
| Jack | Wardle | Imperial College London |
| Thomas | Rawson | Imperial College |
| Richard | Sheppard | Imperial College |
| Tristan | Naidoo | Imperial College |

Table H.17: Pathogen Epidemiology Review Group (PERG) membership
